# Importations of SARS-CoV-2 lineages decline after nonpharmaceutical interventions in phylogeographic analyses

**DOI:** 10.1101/2023.11.10.23298337

**Authors:** S. Goliaei, M.H. Foroughmand-Araabi, A. Roddy, A. Weber, S. Översti, D. Kühnert, A.C. McHardy

## Abstract

The onset of the SARS-CoV-2 pandemic marked a period of substantial challenges as the virus and its variants rapidly spread, placing enormous strain on both society and healthcare systems. Prior to the widespread availability of vaccines, non-pharmaceutical interventions such as reducing contacts, antigenic testing, or travel restrictions were the primary means of reducing viral transmission and case numbers, and quantifying the success of these measures is therefore key for future pandemic preparedness. Using SARS-CoV-2 genomes collected in systematic surveillance, we studied lineage importations for the third, pandemic wave in Germany, employing a large-scale Bayesian phylogenetic and phylogeographic analysis coupled to a longitudinal assessment of lineage importation dynamics over multiple sampling strategies. We evaluated the effect of twelve major nationwide nonpharmaceutical interventions (NPIs) on lineage importations and dissemination within the country. All NPIs were followed by reduced lineage importations, with the most substantial decreases seen for the provision of free rapid tests, the strengthening of regulations on mask-wearing in public transport and stores, as well as on internal movements and gatherings. Most SARS-CoV-2 lineages first appeared in the three states with the largest populations and most cases, and from there spread within the country. Importations began to rise before and peaked shortly after the Christmas holidays. Analysis of SARS-CoV-2 data revealed the substantial effects of free rapid tests and obligatory medical/surgical mask-wearing, suggesting these as key for pandemic preparedness, given their relatively few, negative socioeconomic effects. The approach quantifies the relationships between environmental factors at the host population level to viral lineage dissemination from genomic surveillance data, facilitating similar analyses of rapidly evolving pathogens in the future.

## Introduction

The Severe Acute Respiratory Syndrome Coronavirus 2 (SARS-CoV-2) is a rapidly spreading, highly infectious virus of zoonotic origin causing Coronavirus Disease 2019 (COVID-19) [1]. SARS-CoV-2 was first identified in December 2019 in Wuhan, China, and continued to spread globally, resulting in approximately 770 million confirmed infections and seven million deaths to date [2,3]. Prior to the widespread availability of vaccines, nonpharmaceutical interventions (NPIs) such as reducing contacts, antigenic testing, and travel restrictions were the primary means of reducing SARS-CoV-2 transmission and case numbers in the pandemic.

To trace viral spread across countries, as well as the emergence of novel variants with concerning phenotypic alterations, viral genomic surveillance was established in many countries. This resulted in unprecedented amounts of genome sequences being rapidly generated worldwide. By combining genomic information with sampling times and locations, the spatial dispersal and lineage evolution can be reconstructed with viral phylogeographic techniques. Phylogeographic analyses provide insight into SARS-CoV-2 importations into Europe at the beginning of the pandemic [4]. indicating that it was transported from Hubei, China, to multiple European countries several times between mid-January and early February 2020, before the large outbreak in northern Italy. The first wave of COVID-19 cases was studied in the U.K. and Portugal, both of which had high early sequencing rates and were therefore able to characterize the importation and diversity of lineages in depth [5,6]. For the U.K., international travel led to the importation of over 1000 co-circulating transmission lineages. Both for the U.K. and Portugal, most introductions occurred prior to lockdown measures, with the earliest ones becoming the largest and most persistent lineages post-lockdown [5]. Although Portugal quickly implemented lockdown measures, SARS-CoV-2 was likely circulating in late February, weeks before the first detected case [6]. Contrasting this, a study of Belarus reported few early importations that were largely brought in from neighboring countries, which is consistent with travel data [7].

Following the lifting of the first lockdown measures, the B.1.117 lineage spread in a second infection wave across Europe over the summer of 2020 [8,9], leading to a persistently high volume of cases across Europe, despite B.1.117 having no notable transmission advantage. Further, a study of the third and fourth infection waves in Hong Kong provides insight into the successes and challenges of an elimination strategy [10], as opposed to the mitigation approach adopted by the countries described before. As a result of strict border control measures, there was a low level of importation with the third and fourth waves, which occurred over the period July 2020 – April 2021. Local transmission was much higher in wave four than wave three, which was attributed to reduced compliance with control measures due to pandemic fatigue [10]. Ultimately, SARS-CoV-2 was eliminated during the study period due to contact tracing and mandatory quarantine measures. Further, low levels of importation were maintained due to highly stringent border controls, including a 21-day quarantine on arrival. Similarly, a Bayesian phylogeography analysis showed that Switzerland’s strict border closures alongside the 2020 partial lockdown were effective in controlling the entrance of new lineages into the country [11]. However, later VOCs (Alpha, Delta, and Omicron) exhibited increased transmissibility, making an elimination strategy much more challenging to maintain.

The third infection wave in Europe during spring 2021 largely consisted of the B.1.1.7 (Alpha) lineage, which is thought to have first emerged in Kent or Greater London [12]. Alpha exhibited higher transmissibility compared to previous ones and was not contained by the U.K. lockdown measures [13], which controlled other lineages in late 2020 [14]. Instead, stricter measures had to be introduced in early 2021 [15]. Studying both the third and fourth waves in England, characterized by Alpha and Delta variants, respectively, revealed that their growth was initially masked by falling case counts in more dominant lineages [16].

Systematically assessing the effects of different nonpharmaceutical interventions (NPIs) on viral lineage importations and spread is key for future pandemic preparedness, although currently, these are not entirely clear [17]. Here we systematically analyzed how the NPIs that were successively implemented within a country affected SARS-CoV-2 lineage importations. For this, we used data from representative genomic surveillance collected over the course of the third pandemic wave in Germany, together with large-scale Bayesian phylogenetic analyses. We then identified stable properties across multiple sampling strategies that consistently led to fewer importations into the country and less dissemination across states.

## Methods

### Bayesian phylogenetic and phylogeographic analyses

All available SARS-CoV-2 genome sequences, 1,819,996 sequences overall, and the corresponding metadata, were downloaded from the GISAID database on June 2nd, 2021 [18,19]. Following established approaches, e.g. in the studies of Bbosa et al. [20], Bollen et al. [21], Hodcroft et al. [9], Lemey et al. [8], and Nemira et al. [7], we subsampled viral genome sequences using sampling dates, confirmed case numbers, and the country of origin, to account for varying data availability and ensure computational tractability in Bayesian phylogenetic and phylogeographic analyses. To assess the consistency of results across sampling schemes, we created two more datasets using different genome subsampling schemes for phylogeographic analyses [5,21]. To infer a time-calibrated phylogenetic tree for each dataset, we performed a Bayesian analysis with Thorney BEAST version 0.1.1 (https://beast.community/thorney_beast). Importations of SARS-CoV-2 lineages into Germany were identified as described by du Plessis et al. [5], corresponding to lineages that include at least one German viral genome and descend from an ancestral lineage inferred to have circulated outside of Germany (**Fig. 1A**). Following the inference of importation lineages, we examined the lineage diversity within states using two metrics: the Shannon Index (SI) and lineage evenness [22,23]. To infer the spread of SARS-CoV-2 importation lineages within Germany between German states, we applied a 16-state Bayesian DTA method to the phylogenies of the inferred importation lineages using BEAST. Further details on all steps are described in the Supplementary Methods.

**Fig. 1.**
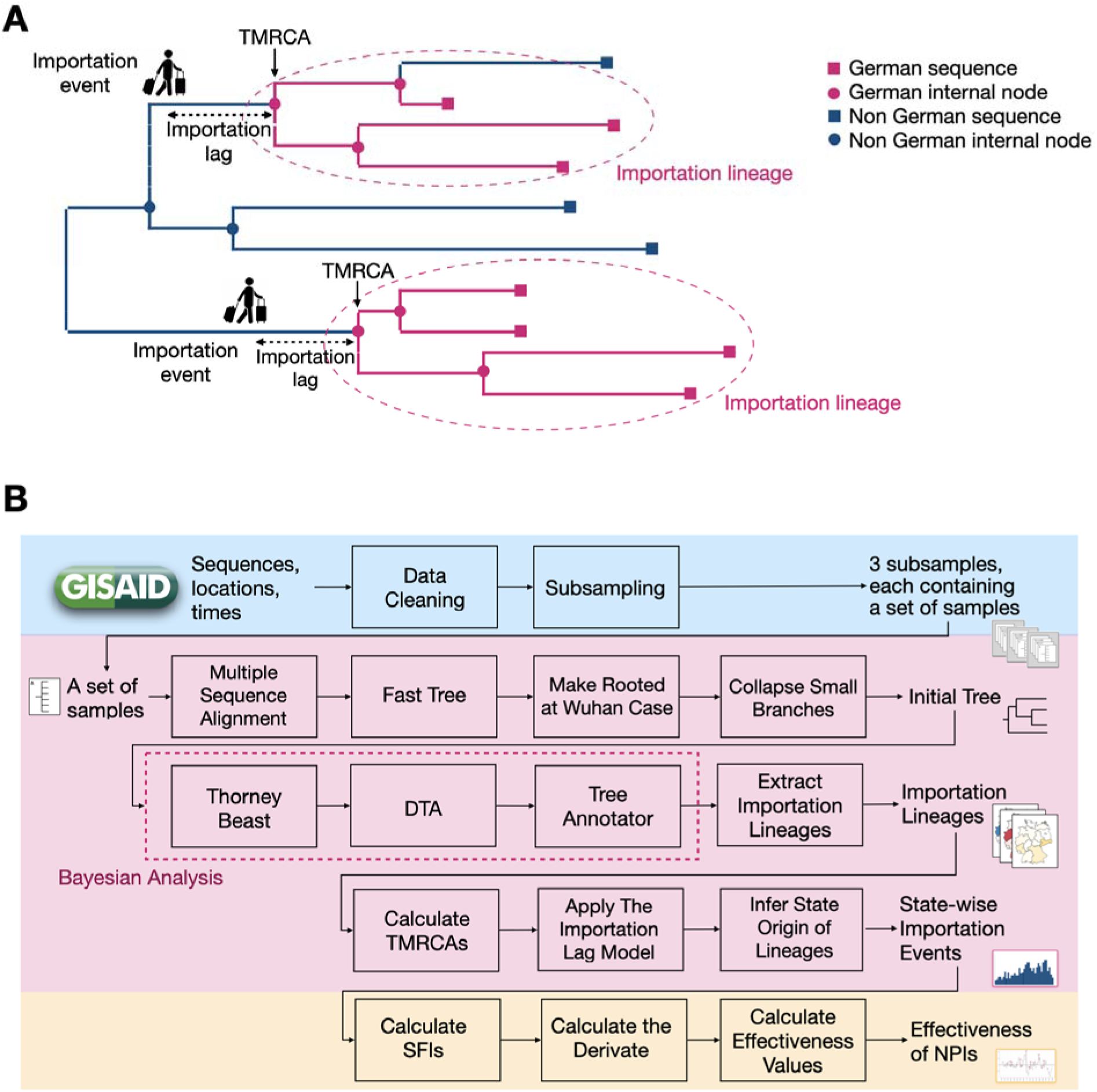
Schematic overview of the method. **(A)** Definition of importation lineages on an inferred phylogeny (orange indicates ancestral lineages and sampled isolates circulating in Germany, blue those found outside of Germany). An importation lineage is defined as a subtree with all nodes (lineages) from Germany that descend directly from an ancestral lineage (node) inferred to have circulated outside of Germany. The time of the most recent common ancestor (TMRCA) is the inferred time of the first junction in an importation lineage. The importation lag is the difference in time between the entrance of an infected person, i.e., the importation event, to Germany and its TMRCA. **(B)** Analytic workflow overview, with (i) genome data selection via sampling (blue), (ii) phylogenetic and phylogeographic analyses of importation lineages using a Bayesian framework (pink), and (iii) assessment of importation lineage dynamics together with NPIs implemented in Germany during the third wave (yellow).

### Relating lineage importation dynamics to nonpharmaceutical interventions

To assess the effect of each nonpharmaceutical intervention on lineage importations, we defined a smoothed importation frequency (SIF) for each day as a weighted sum of the number of importation events that happened over a period of 9 days, starting from the current one. The weight coefficients w(i) are defined as w(i) = i for 1<=i<=5 and w(i)=10-i for 6<=i<=9, creating a triangular graph with a peak at w(5). The measure is calculated as the sum of the number of importation events on the i-th day multiplied by the weight w(i), which produces a smooth measure with a peak that focuses around day 5. Then, we calculated the change rate of SIF as the difference between the SIF for two consecutive days. We determined the effectiveness of an NPI considering the previous seven days and the next seven days as its most influential period, as the maximum reduction in the SIF in the upcoming seven days compared to the highest SIF in the corresponding period before it. This considers importation events over 22 consecutive days overall. Seasonal effects on the number of cases as well as other potential long-term effects do not affect the metric (**Fig. 1B**).

We gathered information about 110 NPIs implemented in Germany from published sources (**Table S1**) [24–27], summarized the national and local NPIs into 12 major NPIs (**Fig. 4**), and classified them as internal NPIs, border control NPIs, and NPIs including both internal and border control measures. Internal measures include workplace closing, restrictions on gathering, restrictions on internal movements, canceling public events, schools closing, obligation on surgical/FFP2 mask wearing, and availability of free antigen tests. We studied the effectiveness of NPIs on lineage importations and spread, first for the country and then for the states with the highest importation activities and population sizes.

## Results

### SARS-CoV-2 lineage importations into Germany

Nationwide viral surveillance through genome sequencing, covering up to 5-15% of registered cases [18], commenced in Germany in January 2021, before the third pandemic wave gained momentum in February 2021 [28]. Of the SARS-CoV-2 isolates sequenced between February and May 2021 in Germany, 71.95% belonged to the B.1.1.7 lineage, with an increasing frequency over time. Additionally, 11.30% belonged to the B.1.177 lineage, which had previously spread across Europe [9]. Notably, B.1.1.7 was predominant among sequenced isolates from the German states of North Rhine-Westphalia, Bavaria, and Baden-Württemberg.

The genomic surveillance program allowed us to conduct a systematic analysis of viral lineage imported into the country during this period using large-scale phylogenetic analysis techniques. We adapted a Bayesian phylogeographic approach to infer a phylogeny from publicly available SARS-CoV-2 genomes up to June 2, 2021 [5]. This global dataset consisted of 1.8 million viral genome sequences, with varying coverage of case numbers in different countries and time periods due to differences in sequencing rates. To address geographic and temporal sampling biases, assess the stability of results, and identify consistent properties across different data subsets, we created three distinct datasets using previously employed sampling strategies [7,8,11]: First, we sampled sequences proportional to the number of confirmed cases in each country for each week; then, we sampled 100 sequences from Germany and 100 sequences from all other countries for each week; and finally, up to 25 sequences from each other country, if available, and 100 sequences from Germany for each week (Methods, **Table S2**). From each of these datasets, we identified SARS-CoV-2 lineages imported into Germany, along with their estimated importation times (Methods).

Many properties remained consistent across the three samplings, while some properties varied. With an increasing number of non-German sequences included in the data sets, the age of the TMRCAs decreased, while the number of importation lineages increased (50:50 vs. 25:100; Table S2, **Fig. S1B,C**). This indicates that the inclusion of more non-German sequences results in the splitting of importation lineages, leading to later TMRCAs and smaller importation lineages and that the numbers of truly circulating lineages are likely higher than the obtained estimates.

We examined the largest lineages that were imported and their subsequent spread, as these had the most substantial impact on Germany during the third pandemic wave. Consistently, only a few large lineages were detected (**Fig. 2A, S1**). In the main sampling, proportional to the number of cases, the 15 largest importation lineages cover 20.3% of the German samples, while the remaining samples were assigned to 661 additional lineages. Also, the absolute number of identified importation lineages varied across samplings.This aligns with sizes of the importation lineages following a power-law distribution, as seen for the UK [5], where growth is primarily seen for the large lineages, and most lineages are small [29].

**Fig. 2.**
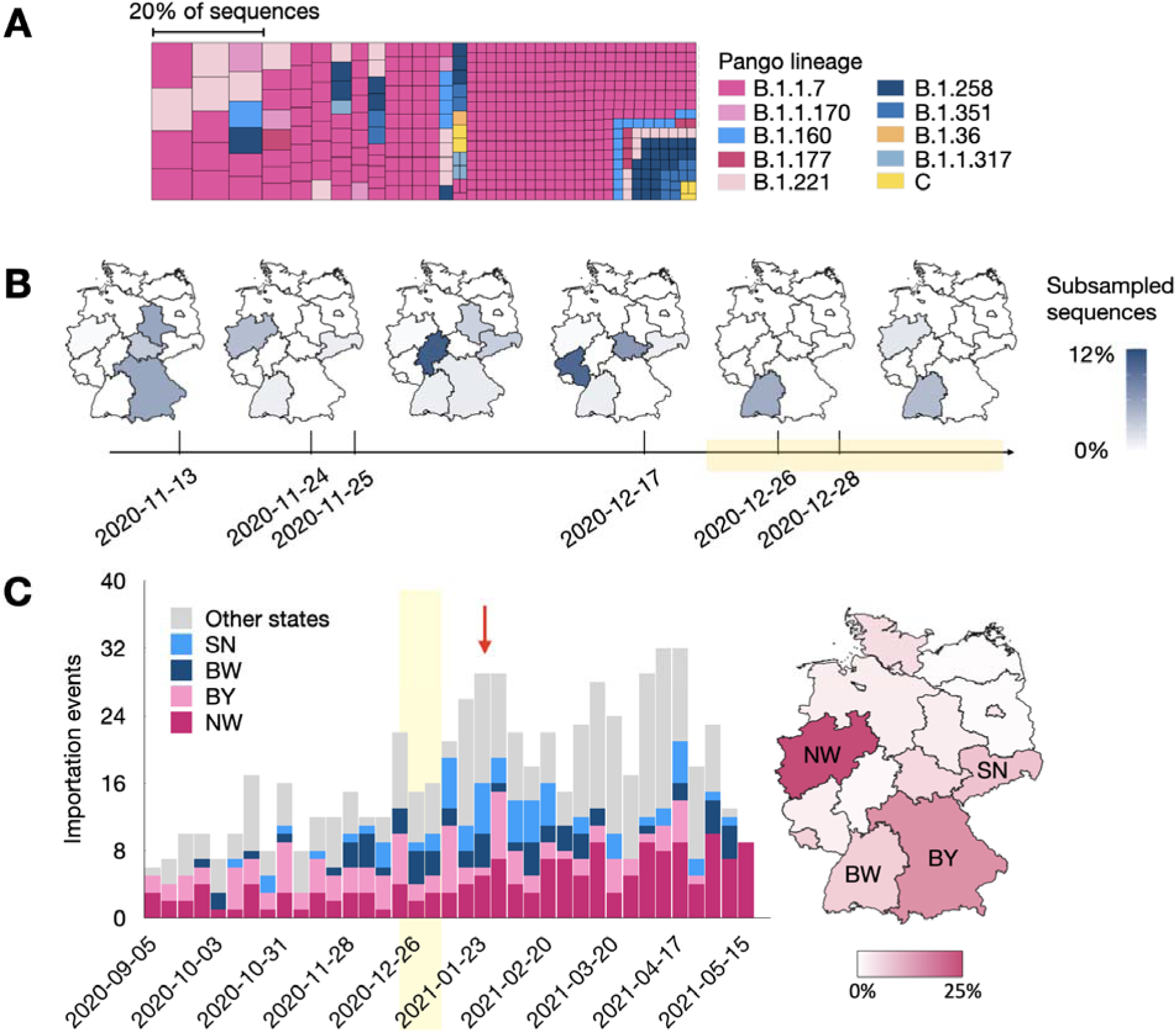
Epidemiological properties of viral importation lineages. **(A)** Importation lineages are colored by the corresponding Pango-lineage. The size of squares represents the size of the importation lineages. **(B)** Estimated importation time and geographic distribution over states for the six largest B.1.1.7 importation lineages. The colors represent the number of viral genomes belonging to the respective importation lineage for each individual state. The light orange block indicates the time period of the Christmas holidays. **(C)** Number of lineages imported into the country per week, colored by the state that they were first observed in, shown individually for the states with the most importation events (dark pink: North Rhine-Westphalia, pink: Bavaria, dark blue: Baden-Württemberg, blue: Saxony) and for the remaining states in gray. The light orange block denotes the Christmas holidays, from 24 Dec 24th of 2020 to Jan. 2nd-9th 2021, depending on state, and the red arrow indicates the peak in importation events after Christmas holidays. See Fig S2 for the results for other samplings The map represents the percentage of detected importation lineages, first observed in individual states (white: fewer, pink: more). NW = North Rhine-Westphalia, BW = Baden-Württemberg, BY = Bavaria, SN = Saxony.

Most importation lineages belong to B.1.1.7 (**Table S3**), the main driver of the third pandemic wave in Germany. Notably, the B.1.1.7 importation lineages responsible for most cases were imported within a two-month period, spanning from mid-November 2020 to the end of December 2020 (**Fig. 2B**). The overall number of inferred importation lineages began to rise shortly before and reached its peak after the Christmas holidays (**Fig. 2C**), indicating that holiday-related travels played a key role in importations and the spread of lineages within the country, consistently across samplings.

### Appearance and diversity of importation lineages within the country

Most importation lineages first appeared in North Rhine-Westphalia (25.1%), Bavaria (15.8%) and Baden-Württemberg (7.1%; **Fig. 2C**) in all samplings, across all Pangolin lineages and B.1.1.7-associated importation lineages only (**Fig. S3**). This indicates the relevance of these three most populous German states for lineage importation and within-country dissemination during the third wave. Stably, the fewest lineages first appeared in Mecklenburg-Western Pomerania and Brandenburg, both 0.5% (**Fig. S4**). The initial appearances correlate with the states’ population sizes (Pearson CC 0.82) and confirmed case numbers (Pearson CC 0.86). There was no correlation with state-wise sequencing rates of the representative SARS-CoV-2 sequencing programme initiated for Germany in January 2021 (Pearson CC −0.01).

Notably, despite having the airport with the most annual number of travelers in Germany, comparatively few (<2%) importation lineages were first observed in Hesse. Among the ten most frequented airports in Germany in 2020, three are located in the state of North Rhine-Westphalia (ranked third, eighth, and tenth), one in Bavaria (ranked second), one in Baden-Württemberg (ranked seventh), while none is found in Mecklenburg-Vorpommern and Brandenburg [30]. While we anticipated that air travel would have a substantial impact on lineage importations, we observed that the relationship to the state-wise population was more pronounced. We hypothesize that the state population is more reflective of the ultimate destination of these travels, while airports serve as intermediate stops.

To assess the diversity of importation lineages within states, we calculated the Shannon Ind x (SI), as by Spellerberg et al. [22], and the lineage evenness for each state. The SI is influenc d by the total number of lineages and their respective size distributions, where having ma y lineages of similar sizes results in larger SI values. Across samplings, the state-wise SI strongly correlates with the logarithm of the number of analyzed sequences. In the case of evenness, t e SI of a state is divided by the number of lineages present in that state [23], providing a direct reflection of the lineage size distribution. SIs were highest for the population-rich states of North Rhine-Westphalia, Bavaria, and Baden-Württemberg, and Saxony, as expected due to the large number of cases and importation lineages observed. The lowest SI was determined for Schleswig-Holstein (**Fig. 3A**), which exhibited a substantially lower SI value than several other states with fewer confirmed cases and circulated importation lineages, as the lineage size distribution is strongly skewed towards one dominant lineage that entered the state before the Christmas holidays (**Fig. S5**), indicating an exceptionally low connectivity to other regions. This result remains stable across samplings (**Fig. S6**). With the exception of Hamburg, Bremen, and Schleswig-Holstein, lower lineage numbers correlated with more equal lineage sizes, higher evenness, across states (**Fig. 3B, S6**). While these relationships are not unambiguously resolved [23], such effects can be explained by niche preemption, which posits that a highly diverse environment, i.e., one with many lineages, is more challenging for incoming lineages to invade.

**Fig. 3.**
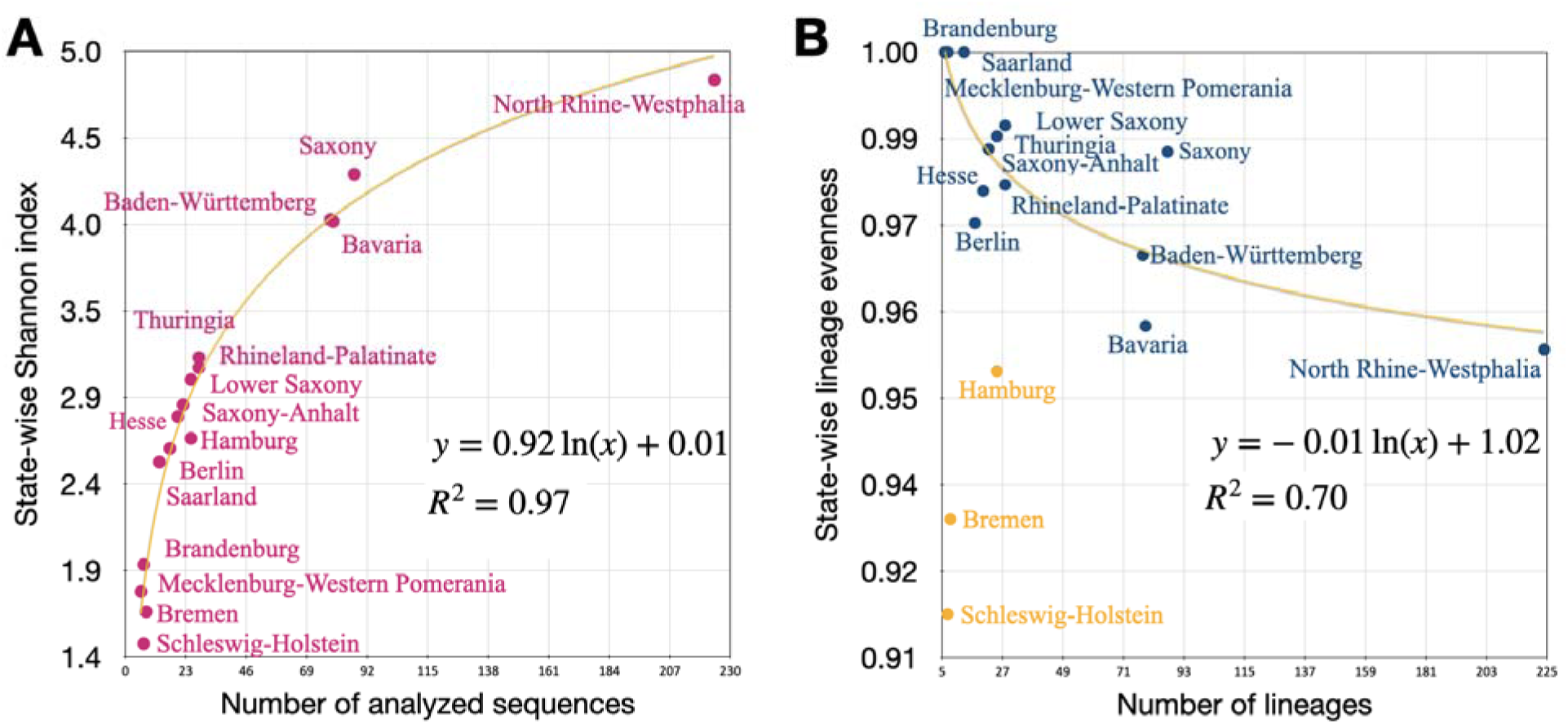
Diversity and Evenness of Lineages. **(A)**: State-wise Shannon index of importation lineage diversity, which increases both with the number of lineages and evenness of lineage abundance distributions, versus the number of analyzed sequences generated after the start of systematic genomic surveillance in Germany on Jan 1st of 2021. The state-wise Shannon index is highly correlated with the logarithm of the number of analyzed sequences. **(B)** State-wise evenness of lineage abundance distributions versus the number of analyzed sequences generated after 01 Jan 2021. The orange curves (**A** and **B**) indicate a fitted logarithmic function. R2 denotes the extent of variance explained by the fitted functions, with orange points indicating outliers excluded from the analysis.

### Lineage importations are reduced after nonpharmaceutical interventions

Throughout the study period, multiple NPIs were implemented and adapted both at the federal and state levels to control viral transmission and case numbers in the population, including both nationwide NPIs and state-specific ones. To evaluate how NPIs affected the rate of lineage importations into the country and states, we gathered information about 4,000 national and local NPIs and summarized them into 12 major NPIs by date (**Fig. 4, Table S1**). We categorized these as internal NPIs (N1, N2, N4, N5, N8, N11), border control NPIs (N3, N6, N9, N10, N12), and NPI N7, which includes both internal and border control measures. Internal NPIs include measures for school closures, workplace closures, canceling public events, restrictions on gatherings, restrictions on people’s movements, obligation to wear surgical/FFP2 masks in public transport (from wearing of any kind of mask, including homemade cloth masks, to medical FFP2 or surgical masks), and availability of free antigen tests. Among the border control NPIs, N3, N6, and N12 made regulations more restrictive until 09-05-2021, when these NPIs were relaxed by allowing vaccinated people to bypass the mandatory quarantine upon arrival. Regarding internal measures, the six NPIs (N1, N2, N4, N5, N7, and N8) made measures stricter until mid-February 2021 when these restrictions were gradually relaxed.

**Fig. 4.**
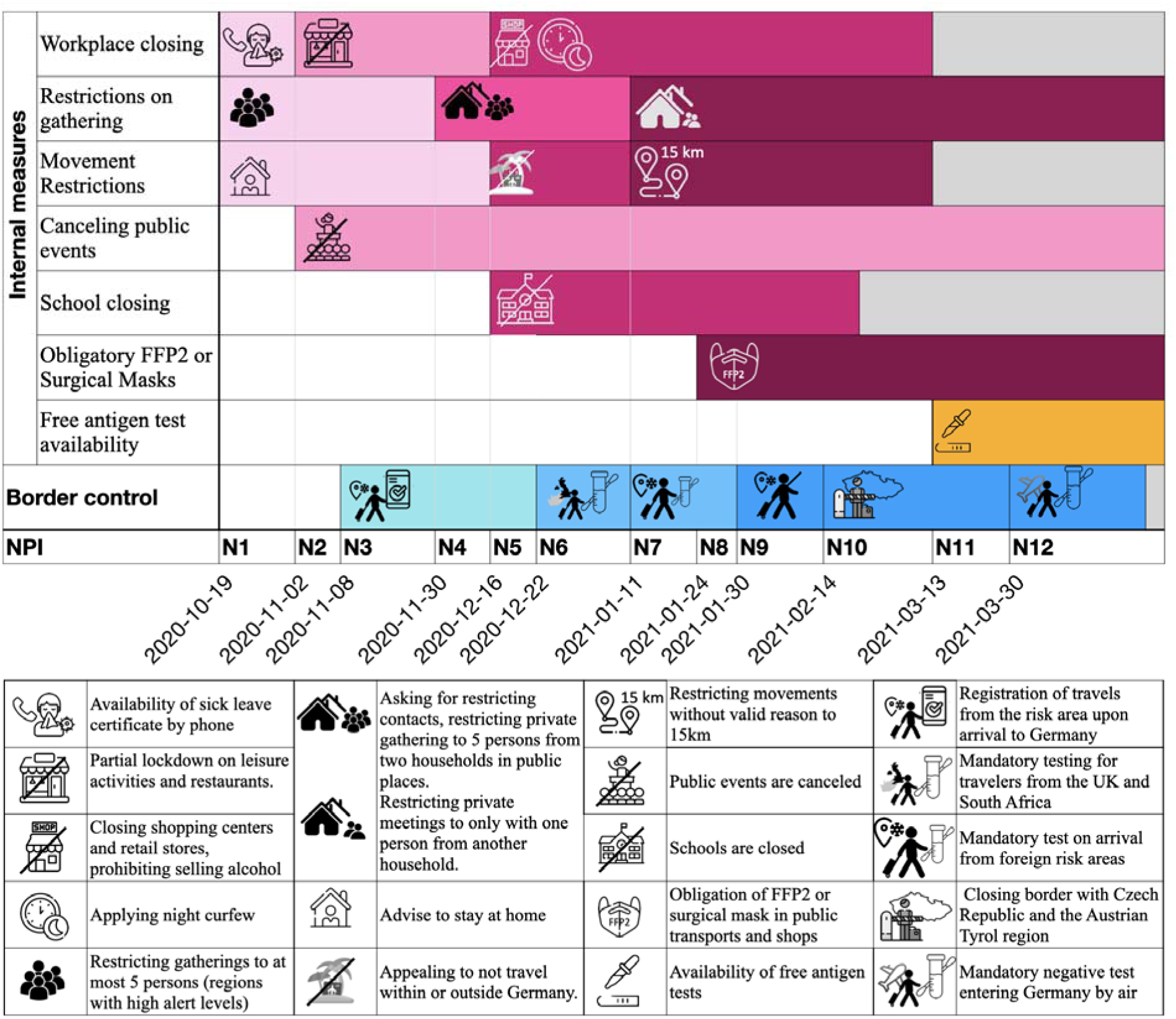
Application date and descriptions for twelve major NPIs implemented in Germany between October 2020 and March 2021. The first seven rows show the details of internal measures over time, while the next row represents the border control measure. In the pink and blue rows, lighter colors represent less restrictive measures, and darker colors represent more restrictive interventions. Gray color indicates relaxation of the restrictions applied at the end of the third wave. Note that the presented intensity and colors are for the illustrative purposes only. N1, N2, N4, N5, N8, and N12 are internal NPIs, while N3, N6, N9, N10, and N12 are border control NPIs. N7 includes both internal and border control measures.

We defined a daily effectiveness measure (DEM) based on the regulations put into effect, from the number of importation events in the previous 7 and the next 14 days (Methods; **Fig. 5A**). Lineage importations into the country were reduced after all twelve major nationwide NPIs as measured by this effectiveness (**Fig. 5**). Across samplings, the availability of free rapid tests starting from 08-03-2021 (N11) was the most effective NPI for the country (DEM=31), along with the strengthening of the mask wearing in public transports and shops on 24-01-2021 (N8), which made surgical/FFP2 mask-waring mandatory instead of wearing any kind of mask, which was required before this. Additionally, the restrictions implemented on 11-01-2021 (N7), which include limiting movements to a 15km radius, restriction of gatherings to be with at most one person from another household, and mandatory testing for all travelers from risk areas, demonstrated high effectiveness in two sampling strategies (**Fig. S7**).

**Fig. 5.**
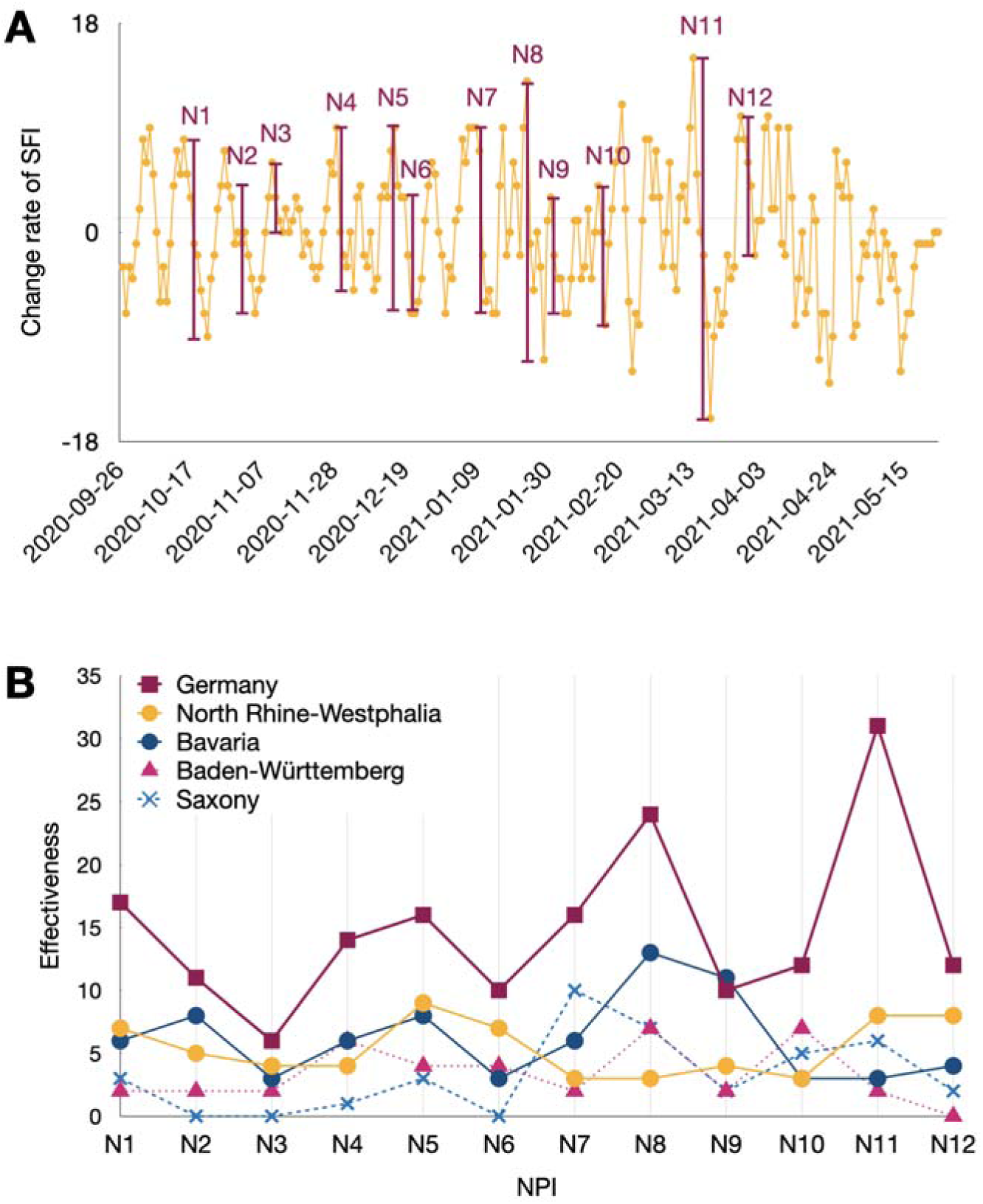
Effectiveness of NPIs. **(A)** Change rate of SIF over time. Red vertical bars indicate the implementation time of the twelve major NPIs in the country, and their heights represent the **effectiveness** metric for that day. **(B)** NPI effectiveness for Germany overall, as well as for North Rhine-Westphalia, Bavaria, Baden-Württemberg, and Saxony.

When examining the effectiveness of NPIs within individual states, N11 emerged as the most effective NPI for North Rhine-Westphalia (DEM=9, in two subsampling strategies, case ratio and 50:50), Bremen (DEM=2, in two subsampling strategies, case ratio and 50:50), and Rhineland-Palatinate (DEM=6, in all three subsampling strategies, **Fig. S8**). NPI N8 was the second most effective NPI for the country (DEM=24), as well as for Baden-Württemberg (DEM=9, in all three subsamplings), and Bavaria (DEM=15, in two subsampling strategies, case ratio and 50:50). Furthermore, NPI (N7) was also the most effective NPI for Lower Saxony (DEM=6, in all three subsampling strategies) and Bremen (DEM=2, in two subsampling strategies, case ratio and 50:50).

Overall, internal NPIs (average DEM=18.83) appeared more effective for the country than border control ones (average DEM=10). Hamburg, Bremen, Lower Saxony (only in 25:100 and 50:50 subsamplings), Hesse, Saarland, Brandenburg, and Berlin (only in the second and third subsamplings) have a higher average of effectiveness in border control NPIs than internal NPIs, indicating the benefit of border control NPis in these states. This is a stable result across the three sampling strategies (**Fig. S8**).

### Inter-state lineage spread

We analyzed the spread of lineages within Germany across states using a Bayesian 16-state DTA model and determined the number lineage transfers between states (Methods, **Fig. 6A**). The most lineages (ten) spread from North Rhine-Westphalia to Baden-Württemberg with all subsampling strategies, followed by lineage transmissions from North Rhine-Westphalia to Saxony, from Bavaria to Baden-Würtemberg, and from North Rhine-Westphalia to Lower Saxony. Four states, namely North Rhine-Westphalia, Bavaria, Saxony, and Baden-Württemberg, which except for Saxony are the most highly populated states, played the most substantial role in lineage spread within Germany. Specifically, North Rhine-Westphalia had the most outward-going lineage spread (56), followed by either Bavaria (23) or Saxony in all samplings, while Lower Saxony and Mecklenburg-Vorpommern had no lineage transmissions to any other state.

**Fig. 6.**
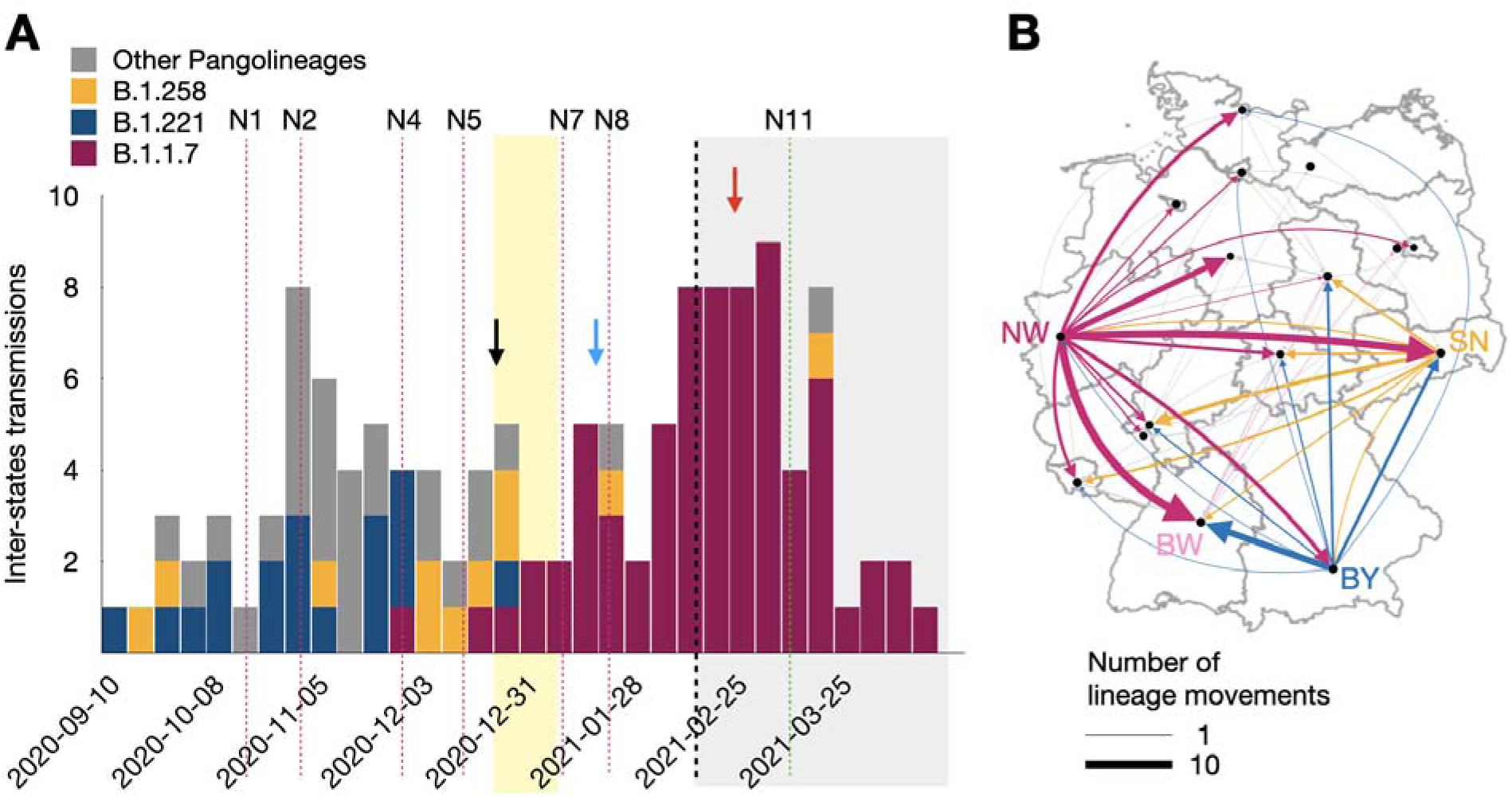
Inter-state spread of lineages. **(A)** Number of inter-state spread of lineages, colored by the Pango-lineage. The number of B.1.1.7 lineage transmissions rise at the beginning of Christmas holidays and after that (black arrow and blue arrow). There is a peak of B.1.1.7 lineages inter-state transmission in mid February 2021 (red arrow) when NPI restrictions were gradually relaxed. **(B)** Spread of transmission lineages across the states within Germany. Arrow widths depict the number of detected migration movements and the color reflects state of origin. The abbreviation of the states are provided in the legend of Fig. 2.

The inter-state spread of B.1.1.7 lineages started to rise before Christmas and reached a peak after the Christmas holidays (**Fig. 6B**), suggesting the impact of these holidays on the dissemination of these lineages within the country. Another peak occurred in late February, coinciding with the relaxation of internal restrictions, such as the closure of workplaces, movement restrictions to 15 kilometers, and the closure of schools. This suggests that the restrictions imposed by these internal NPIs had previously limited the spread of lineages across states. These two results were consistent in all three subsampling strategies (**Fig. S9**).

## Discussion

The onset of the SARS-CoV-2 pandemic brought about considerable difficulties for both societ and healthcare systems. Before vaccines were widely accessible, strategies to control SARS-CoV-2 transmission and cases mainly relied on nonpharmaceutical interventions such as reducing social interactions, using antigenic testing, and imposing travel limits. Studies have shown declining lineage importations and persistence after lockdowns for the UK and Switzerland [5,11], and decreasing case numbers after establishment of NPIs [31–34]. To prepare for future pandemics and to identify the most effective measures, it is crucial to evaluate how SARS-CoV-2 lineage importations into the country and their subsequent internal dissemination relate to the nonpharmaceutical interventions that were put into effect. Here, we performed a comprehensive analysis of how SARS-CoV-2 lineage importations were reduced after a range of NPIs that were implemented within Germany over the course of the third infection wave of the pandemic. We assessed viral genomes collected in representative genomic surveillance together with information on their sampling times and locations using large-scale Bayesian phylogenetic analyses, adapting the framework developed by du Plessis et al. [5].

The largest reductions of SARS-CoV-2 lineage importations were seen in Germany after the provision of free rapid tests, and the strengthening of regulations on mask wearing from wearing any kind of mask to obligatory surgical/FFP2 mask wearing in public transport and in stores, and on internal movements and gatherings. The provision of free rapid tests and the strengthening of regulations on mask-wearing have substantially fewer socioeconomic effects than restrictions on public gatherings and internal movements, which more strongly affect day-to-day activities [35]. We furthermore determined a particular relevance of the German Christmas holidays and highly populated states for SARS-CoV-2 lineage importations into the country and their subsequent internal dissemination, indicating a benefit in considering holiday-related dynamics when planning NPIs. Interestingly, studies of the dissemination of seasonal influenza viruses have shown an opposite effect, in that winter holidays delay epidemic peaks [36–38], likely because children play a key role in household transmission, due to their reduced immune protection compared to adults, which is a notable difference between seasonal epidemics and a pandemic involving novel infectious agent, where initial population-wide immune protection is low.

The substantial number of lineage importations into the German states with large populations and incidence numbers, as well as the prevalence of inter-state transmissions from these states highlights their relevance for maintaining the momentum of the pandemic in the third wave. These states most strongly contributed to the subsequent dissemination of imported SARS-CoV-2 lineages within the country, as most lineages were first observed in these states and transferred from thereon to others, and an effect of internal NPIs in controlling this internal spread was evident. This aligns with a study of the U.K. [12], where cell phone-derived population mobility revealed in phylogeographic analysis the spread of SARS-CoV-2 lineages dominantly from the Greater London area.

Bayesian phylogenetic analyses of SARS-CoV-2 viral genomes have provided profound insights into pathogen evolution and dissemination during the pandemic [15,16,39]. We studied SARS-CoV-2 lineage importations into Germany using this approach with one of the most comprehensive sets of viral genomes analyzed to date, covering 1.8 million genome sequences. To mitigate temporal and regional, e.g. across countries and states, variations in sequencing of infected patient samples, we used several sampling strategies on these data: random selection by region and time period proportional to the confirmed case numbers, as by Nadeau et al., Lemey et al., and Bollen et al. [4,8,21], using an equal number of samples from inside and outside of a country for a time period [5] and using a specific ratio for the number of samples from Germany to the number of samples from each other country, similar to a lag model inference by du Plessis et al. [5]. This allowed us to assess the potential effects of common sampling strategies on the results, such as e.g. the numbers of identified importation lineages, their sizes and importation times across data sets. We identified many consistent properties across samplings, such as the very skewed abundance distribution of imported SARS-CoV-2 transmission lineages, with very few large lineages consistently observed, the number of imported lineages peaking after the German Christmas holidays, as well as the most populous German states having the most importations and contributing the most as a source of lineage spread within the country across states, and lineage importations being reduced following all twelve major NPIs that were implemented over the course of the third wave of the pandemic in Germany. Consistently, also the states with the fewest importations had the least relevance in spreading lineages within the country. There was greater variation observed for other properties though, such as the TMRCAs and the absolute number of importation lineages, with a tendency of increasing the ratio of in-country to out-of-country samples and absolute number of samples from within the country to cause inferred importation lineages to merge, thus reducing the absolute numbers of importation lineages and resulting in larger lineages with earlier TMRCAs, indicating that these properties need to be interpreted cautiously.

By integrating a phylogenetic and phylogeographic analyses to infer lineage importations with a longitudinal assessment of importation dynamics, we provide an analytical framework for exploring the connections between environmental factors, such as holiday and nonpharmaceutical intervention at a population level to lineage dissemination. The effectiveness measure that we introduce allows us to systematically compare and quantify links between NPis and importations [7,11]. Through the design of this effectiveness measure, which assesses temporally local changes in the decline of lineage importations, potential longer-term effects on lineage importations, such as a seasonality, do not affect the results. However, it is conceivable that confounding variables that coincide in time with a measure might contribute to the observed effects, such as behavioral changes resulting from increased concern or NPI-related fatigue in the population.

The SARS-CoV-2 pandemic was the first in which viral genomic surveillance was used to systematically monitor infection dynamics, enabling almost real-time insights into the evolution and dissemination of viral lineages worldwide, as well as allowing to link public health measures to these processes. Substantial efforts are being invested in improving preparedness in terms of rapid creation of vaccines, diagnostics and therapeutics for future pandemics [40]. The results of this study and others demonstrate that establishing and analyzing the results of systematic viral genome surveillance is also essential for linking environmental factors to pathogen dissemination, informing about the relevance of non pharmaceutical measures for the early days of an emerging pandemic.

## Contributors

SG, MHFA developed the codes, performed experiments and analyzed the data together with AMH. AMH conceived the study. All authors discussed the results. SG, MHFA, and AMH wrote the article with comments by all authors.

## Declaration of interests

We declare no competing interests.

## Data availability

The dataset was downloaded from GISAID and is accessible through their website. The Accession IDs for genome sequences are available at https://github.com/hzi-bifo/covid-germany-mcmc.

## Acknowledgements

We gratefully acknowledge all data contributors, i. e., the authors and their originating laboratories responsible for obtaining the specimens, and their submitting laboratories for generating the genetic sequence and metadata and sharing via the GISAID Initiative, on which this research is based. The authors gratefully acknowledge funding by the German Centre for Infection Research (DZIF project number TI 12.002_00) and funding by the German Research Foundation (DFG), for the Excellence Cluster RESIST EXC 2155 project number 390874280, as well as all data contributors, i.e, the authors and their originating laboratories responsible for obtaining the specimens, and their submitting laboratories for generating the genetic sequence and metadata and sharing via the GISAID Initiative, on which this research is based.

## Declaration of generative AI and AI-assisted technologies in the writing process

During the preparation of this work the authors used ChatGPT in order to improve the grammar of the text. After using this tool/service, the authors reviewed and edited the content as needed and take full responsibility for the content of the publication.

## Notes

### Competing Interest Statement

The authors have declared no competing interest.

### Funding Statement

This study was funded by German Centre for Infection Research (DZIF project number TI 12.002_00) and by the German Research Foundation (DFG), and the Excellence Cluster RESIST EXC 2155 project number 390874280.

