## Supplementary Materials for "Importations of SARS-CoV-2 lineages decline after nonpharmaceutical interventions in phylogeographic analyses"

### Supplementary Materials: Importations of SARS-CoV-2 lineage and spread decline after nonpharmaceutical interventions in phylogeographic analyses

#### Supplementary Methods

##### Data preprocessing

All available nucleotide sequences of SARS-CoV-2, including sequences from the third wave, and the corresponding metadata, were downloaded from the GISAID database on June 2nd, 2021.[1,2] In total, 1,819,996 sequences were available. Samples with missing or ambiguous locations and erroneous dates, i.e., with sampling dates earlier than the first confirmed occurrence of the respective lineage, were removed. Low-quality samples, corresponding to sequences shorter than 28,000 bp or with more than 1000 ambiguous bases, were removed as well (see Phylogenetic analyses).

##### Data sampling schemes

Bayesian phylogeography is a technique to study the complex evolutionary and epidemiological processes of viral pathogen spread, allowing the integration of genomic information with different types of metadata and epidemiological spread models.[3] As these methods are computationally intensive, they cannot directly process the hundred of thousands of viral genome sequences generated during the SARS-CoV-2 pandemic. Instead, several studies employed techniques to reduce the input data, such as splitting data into batches according to monophyletic clades.[3,4] Additional efficiency was obtained by tuning the parameters for Bayesian inference on a subset of the data, to accelerate the subsequent analysis.[4]

Following these approaches, we adapted a subsampling approach to ensure computational tractability. The data set was stratified based on the Pango lineage assignments.[5,6] Specifically, we identified twelve Pango lineages with more than 1000 sequences overall and more than 40 from Germany: A, B.1.1.7, B.1.1.519, B.1.1.70, B.1.1.317, B.1.177, B.1.160, B.1.221, B.1.36, B.1.258, B.1.351, and C. Among sequences assigned to these Pango lineages, we removed sequences with invalid dates or dates prior to the first observed case of their respective Pango lineage (as identified by Rambaut et al. [5]). This process resulted in a sequence set eligible for subsampling, covering the time frame from December 30, 2019, to May 31, 2021.

Similar to the studies of Bbosa et al., Bollen et al., Hodcroft et al., Lemey et al., and Nemira et al., [7–11] we subsampled viral genome sequences using sampling dates, confirmed case numbers, and the country of origin, to account for varying data availability. Specifically, we set the maximum number of total samples per week to 10,000 across all countries, and calculated the maximum sample size per country by distributing this value (10,000) proportionally to the number of confirmed cases for the countries. For each country, we then either sampled the specified number of sequences or all available sequences, if the number was lower. To ensure that the main lineages were well sampled, we furthermore added the five earliest and latest samples for each lineage to the data set. The final dataset includes 1194 samples from Germany and 16,486 from other countries.

To assess the consistency of results across sampling schemes, we created two more datasets using different genome subsampling schemes for phylogeographic analyses.[4,8] First, we randomly subsampled equal numbers of up to 100 non-German sequences per week from all countries combined together with 100 German sequences, depending on availability. Second, we randomly sampled up to 25 sequences per week from the genome dataset for each country other than Germany, and 100 sequences per week for Germany. To ensure the main lineages were sampled, we then added the five earliest and latest samples for each, as before, and randomly sampled additional sequences to reach at least 1000 sequences for each lineage per dataset. These further datasets contain 2689/2691 sequences from Germany and 1932/25,107 non-German sequences, respectively.

The **subsampling strategies** used in this study are referred to as follows:

- **Case ratio**: The sampling strategy with number of samples for each week and each country proportional to the number of confirmed cases.
- **50:50**: The sampling strategy with 100 samples from Germany and 100 samples from the rest of the world, per each week.
- **25:100**: The sampling strategy with 100 sequences from Germany and 25 sequences from each country per week.

**Phylogenetic analysis**

To create an initial multifurcating “template” tree for the subsequent Bayesian inference, we used the sarscov2phylo workflow version 22-7-20 for each dataset. This workflow first creates a reference-based multiple sequence alignment with mafft version v7.205 (2014/10/20)[12] using the WH1 sequence NC_045512.2 as reference. The workflow then masks any 7bp window with at least two uninformative characters (N or gap), and the first and last 30bp of every sequence. Then it removes sites with more than 50% gaps, after converting N's to gaps, as well as sites suggested by https://github.com/W-L/ProblematicSites_SARS-CoV2/ and filters sequences that are shorter than 28,000 bp or have more than 1000 ambiguities. A tree is then inferred using fasttree version 2.1.11 [13] and rooted using the sequence Wuhan/WH04/2020 as an outgroup. Short branches with a length of fewer than $5x10^{-6}$ mutations per site are collapsed, to create a multifurcating template tree for the subsequent Bayesian inference.

To infer a time-calibrated phylogenetic tree for each dataset, we performed a Bayesian analysis with Thorney BEAST version 0.1.1 (https://beast.community/thorney_beast). This method takes a multifurcating template tree to constrain topologies, as well as the topologically consistent uncollapsed trees from the previous step as starting trees, together with the sequence sampling dates as input. As by du Plessis et al.[4] a strict molecular clock model was used, with an initial clock rate of $7.5*10^{-4}$ substitutions/site/year and a skygrid coalescent prior.

For each analysis, five independent chains of Markov Chain Monte Carlo (MCMC) were run for 300 million steps. For each chain, trees were sampled every 100,000 steps, and the first 15 million steps were discarded as burn-in. Convergence was ensured using LogAnalyser v1.10.5 of the BEAST package by confirming that Effective Sample Sizes in the merged chain were above 200. Further, a two-state Bayesian asymmetric discrete trait analysis (DTA)[14] was used for phylogeographic reconstruction, and internal node locations (Germany or non-Germany) were inferred for all sampled trees, as by du Plessis et al.[4] For each inferred phylogeny for the subsampled sequence set, two MCMC chains were run for 5 million steps, sampling every 4500 steps and discarding the first 10% of each chain as burn-in. Then a maximum clade credibility (MCC) tree was created with TreeAnnotator for each sub-tree, with a location assigned to each internal node.

##### Identifying importation lineages

Importations of SARS-CoV-2 lineages into Germany were identified as described by du Plessis et al.,[4] corresponding to lineages that contain at least one German viral genome and descend from an ancestral lineage inferred to have circulated outside of Germany (**Fig. 1A**). Different from [4], we also included single sample lineages in the analysis, as these represent larger lineages in the sampled dataset. Based on the size of the lineage and the time of the most recent common ancestor (TMRCA), each lineage's importation time was calculated using an importation lag model.

Following the inference of importation lineages, we examined the lineage diversity within states using two metrics: the Shannon Index (SI) and lineage evenness.[15,16] These measures were derived from the analysis of both the number of lineages and the number of samples associated with each lineage. States exhibiting higher SI values indicate a greater diversity of lineages, accompanied by a larger number of samples, suggesting the presence of states harboring more significant lineages.

##### Inferring inter-state spread of lineages

To infer the spread of SARS-CoV-2 importation lineages within Germany between German states, we applied a 16-state Bayesian DTA method to the phylogenies of the inferred importation lineages using BEAST. This assigns states to the internal nodes of the phylogeny. Importation lineages, which are represented as MCC phylogenies with all internal and leaf nodes labeled as belonging to Germany, were combined, and a 16-state non-symmetric model with 240 parameters was simulated in two independent runs for 5 million steps. The states for samples were obtained from their metadata information and the samples without state information were removed. The Bayesian model parameters, including likelihood, clock rate, and between-state transmission rate parameters, all exhibited ESS values exceeding 200 after analyzing two combined log files. Finally, 500,000 first samples of the inferred trees were ignored, and the rest were sampled with a rate of 1 over 4500. An MCC tree was then determined from the tree sample as the final tree. A pair of parent-child nodes in the MCC tree with different inferred states represents a lineage movement across states, and we assign each movement to the mid-time of the branch, as the average of the times associated with the parent and child nodes.

**Table S2.** The number of German sequences, non-German sequences, and German to non-German sequences ratio, for the three subsampling strategies used.

| Subsampling | Case Ratio (main subsampling) | 50:50 | 25:100 |
|---|---|---|---|
| German Seqs | 1194 | 2696 | 2691 |
| Non German Seqs | 16486 | 1932 | 25107 |
| German/non German | 0.07 | 1.39 | 0.11 |

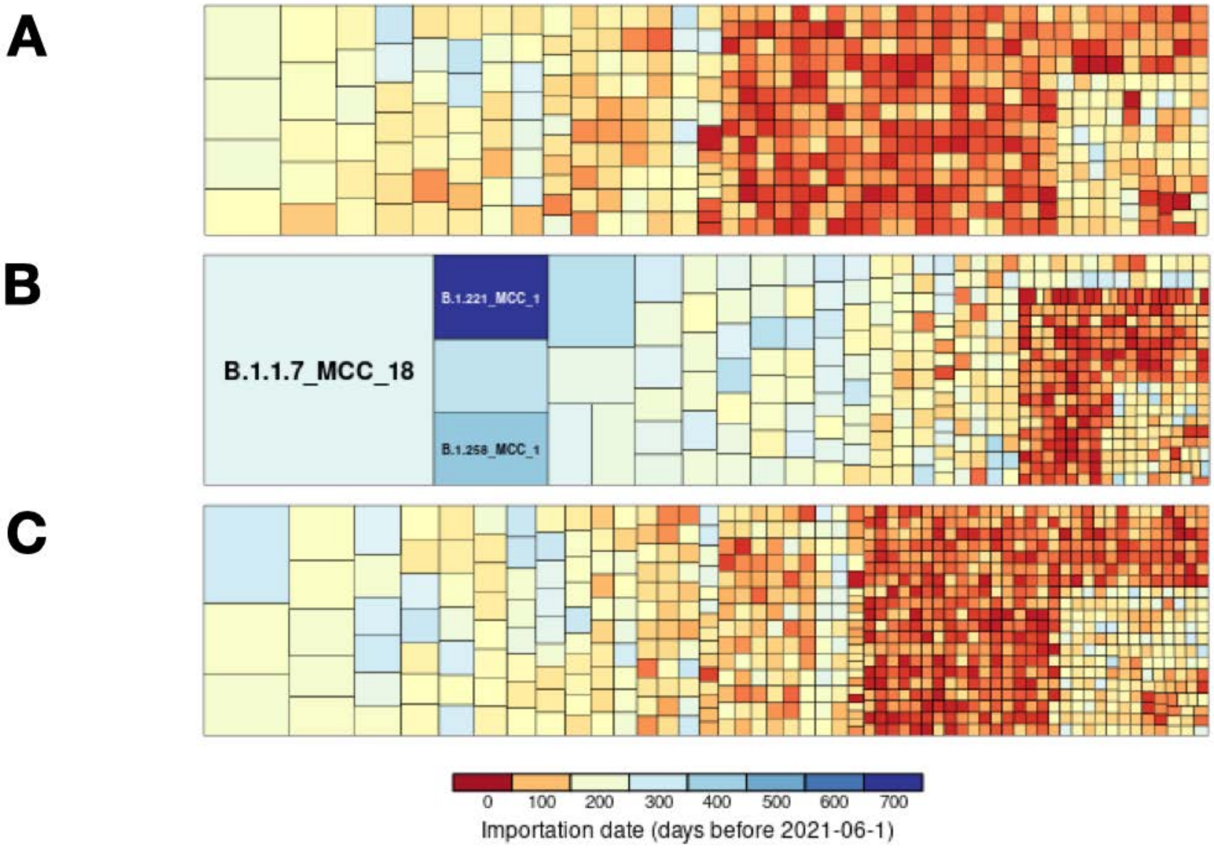


**Fig. S1 - Comparison of importation lineages in different subsampling strategies.** Importation lineages in three sub-samplings **(A)** case ratio, **(B)** 50:50, and **(C)** 25:100. The lineages are colored by importation date (blue is oldest, red is most recent). Consistently, very few large lineages were identified in each subsampling. In comparison to the main subsampling (**A**), both second and third subsampling (**B**, **C**)  had more German sequences (**Table S2**). As it was expected, **B** and **C** had more small lineages (**Table S3**). Also, **B** and **C** had a higher German to non-German sequence ratio, so we expected that lineages to be merged. As it was expected, **B** and **C** had larger lineages and TMRCAs are shifted to earlier time (more blue colors in big lineages). In comparison to the third subsampling (**C**), the second subsampling (**B**), has a higher German to non-German sequences ratio. As it was expected, **B** had larger lineages and TMRCAs are shifted to earlier time (more blue colors).

**Table S3.** The number of identified importation lineages and sequences across data sampling strategies.

| | Case ratio subsampling | | 50:50 subsampling | | 25:100 subsampling | |
|---|---|---|---|---|---|---|
| **Lineage size** | **lineages** | **sequences** | **lineages** | **Sequences** | **lineages** | **sequences** |
| **All** | 677 | 1194 | 624 | 2689 | 1181 | 2691 |
| **1** | 492 | 492 | 356 | 356 | 799 | 799 |
| **2 to 10** | 172 | 507 | 239 | 840 | 355 | 1192 |
| **11 to 100** | 11 | 193 | 25 | 575 | 27 | 700 |
| **> 100** | 0 | 0 | 4 | 918 | 0 | 0 |

130

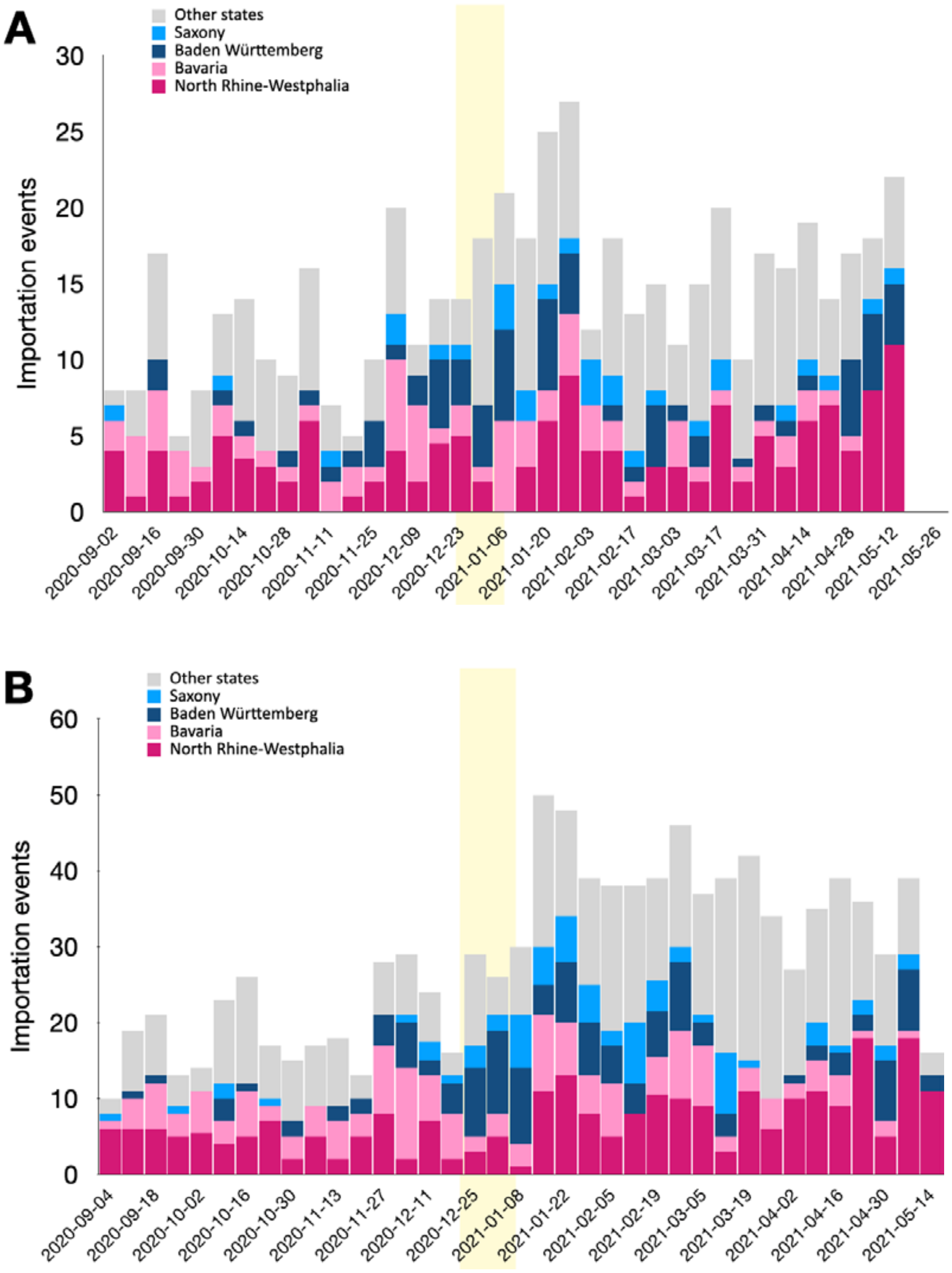


**Fig. S2 - Weekly number of importation events**. Weekly number of importation events, colored by the state of the oldest sequence. **(A)** 50:50 subsampling **(B)** 25:100 subsampling.

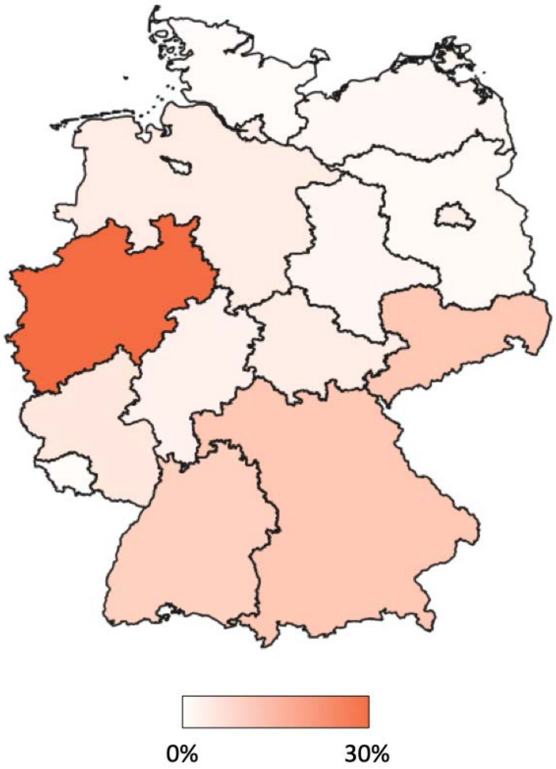


**Fig. S3 - Origin state of importation lineages of B.1.1.7 sequences.** The percentage of B.1.1.7 importation lineages, first observed in states (white: less importation lineages first observed in the state, orange: more importation lineages first observed in the state). The states with the most importation lineages are North Rhine-Westphalia (27.9%), Bavaria (10.5%), Saxony (10.5%), Baden-Wurttemberg (9%).

135

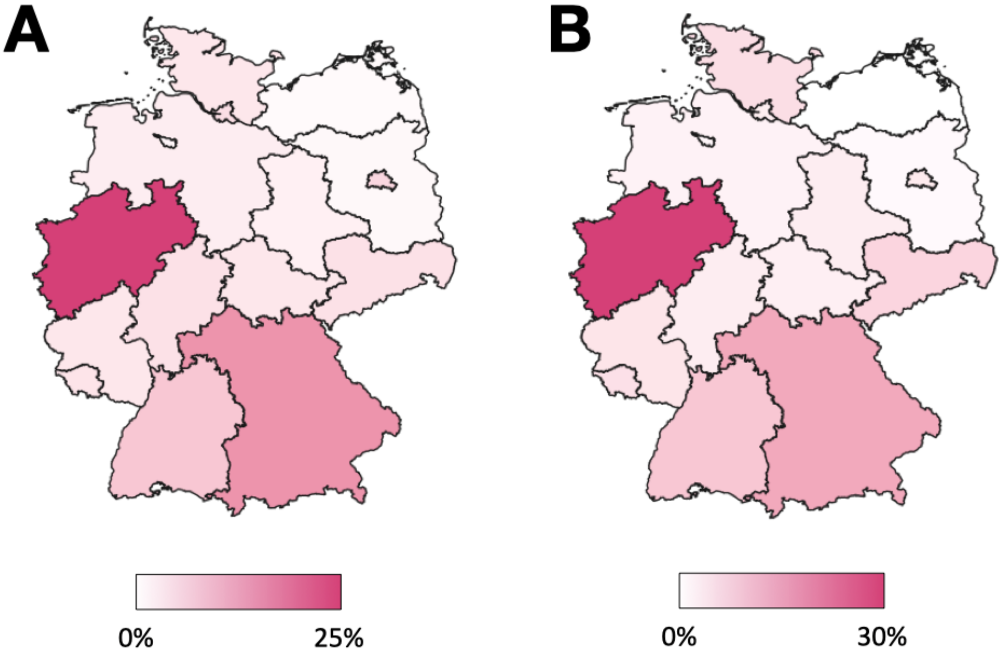


**Fig. S4 - Origin state of importation lineages.** The percentage of detected importation lineages, first observed in states (white: less importation lineages first observed in the state, red: more importation lineages first observed in the state). **(A)** In 50:50 subsampling, most importation lineages belong to North Rhine-Westphalia (26.9%), Bavaria (15.9%), Baden-Wurttemberg (8.3%), and Berlin (6%). **(B)** In 25:100 subsampling, most importation lineages belong to North Rhine-Westphalia (29.6%), Bavaria (14%), Baden-Wurttemberg (9.2%), Saxony (6.9%).

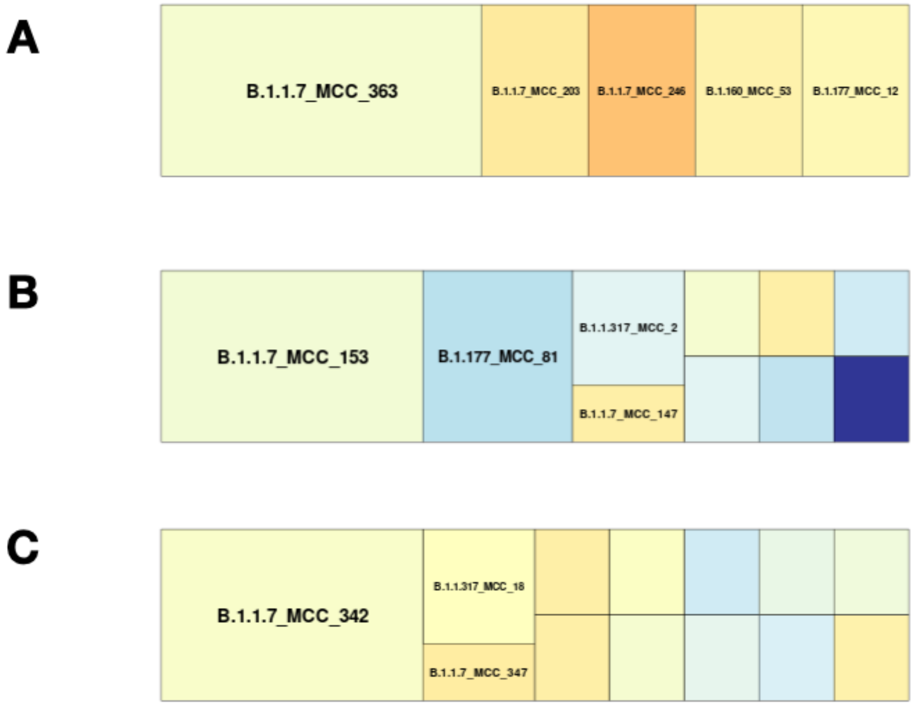


**Fig. S5 - Comparison of lineages imported into Schleswig-Holstein.** Comparison of inferred importation lineages into Schleswig-Holstein by the three subsampling strategies, colored by the importation time (orange = most recent, blue=earlier). **(A)** Case-ratio subsampling, **(B)** 50:50 subsampling, **(C)** 25:100 subsampling.

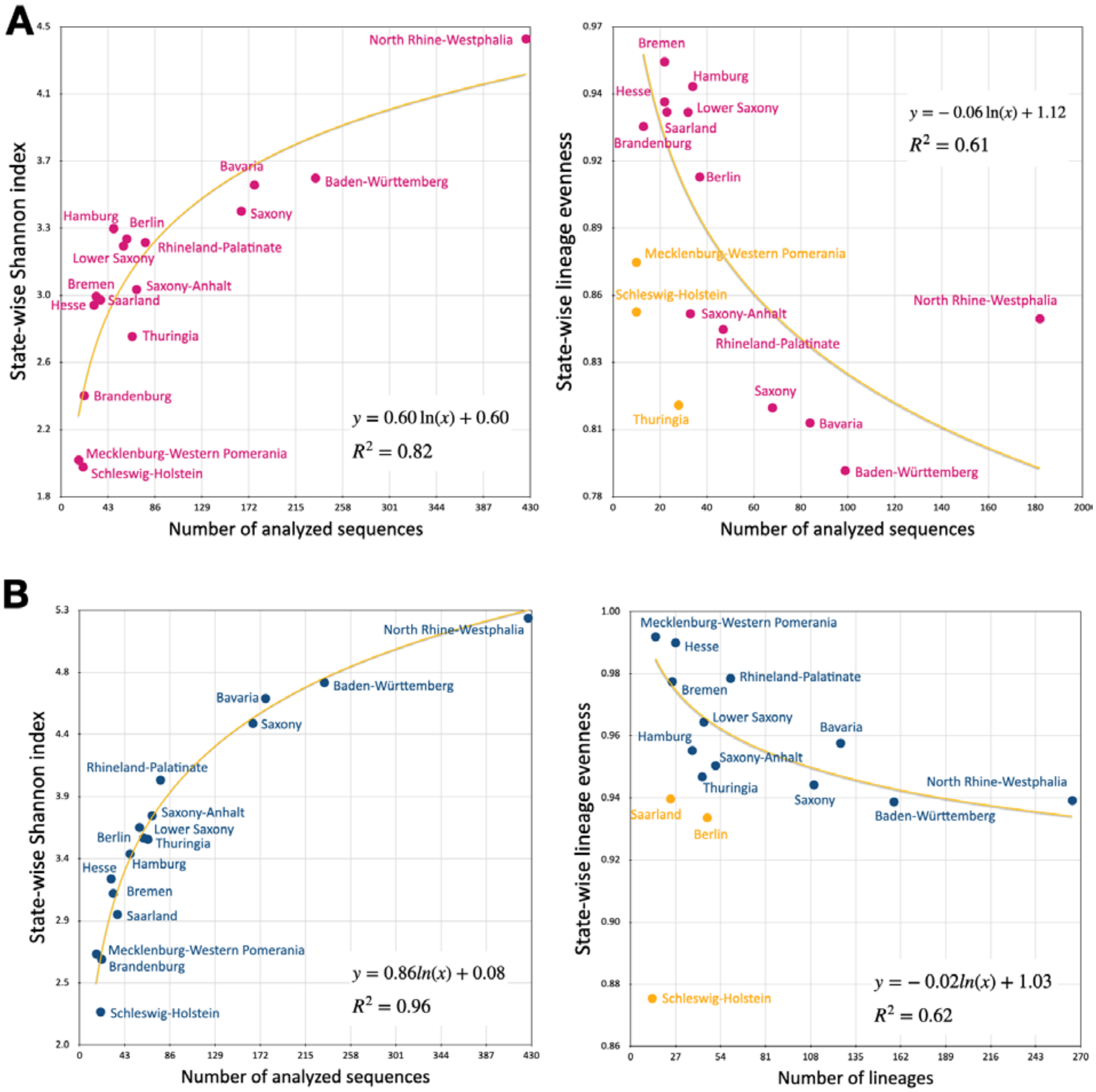


**Fig. S6 - Shannon index and evenness of importation lineages across states**. State-wise Shannon index and lineage evenness. **(A)** 50:50 subsampling, **(B)** 25:100 subsampling. The yellow curve represents the fitted logarithmic function to the data, with yellow points indicating outliers removed during fitting.

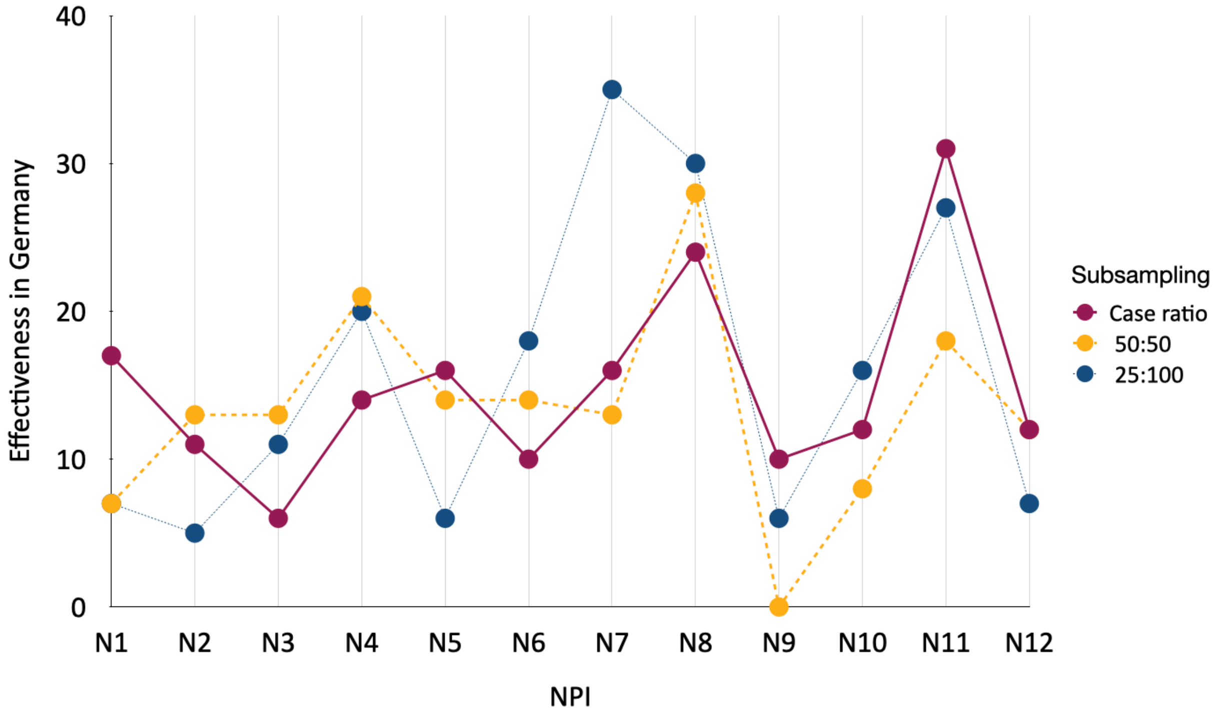


**Fig. S7 - Effectiveness of NPIs.** Effectiveness of NPIs in the country, for three subsamplings case ratio, 50:50, and 25:100. NPIs N11, N8, and N7 are consistently highly effective in all three subsamplings.

NPIs:

- N1: Restrictions on gatherings, workplaces, internal movements
- N2: Restrictions on gatherings, workplaces, internal movements
- N3: Border control, Mandatory Quarantine for high risk areas.
- N4: Restrictions on gatherings, internal movements (Bavaria)
- N5: Restrictions on workplaces, internal movements, schools
- N6: Border control (UK, South Africa)
- N7: Restrictions on gatherings, internal movements, border control
- N8: Border control (Czech Republic, USA, Spain, Portugal)
- N9: Border control (UK, Brazil, Ireland, Portugal)
- N10: Border control (Czech Rep, Tyrol region of Austria)
- N11: Availability of free tests
- N12: Border control (Czech Rep, France, Slovakia, all passengers by air)

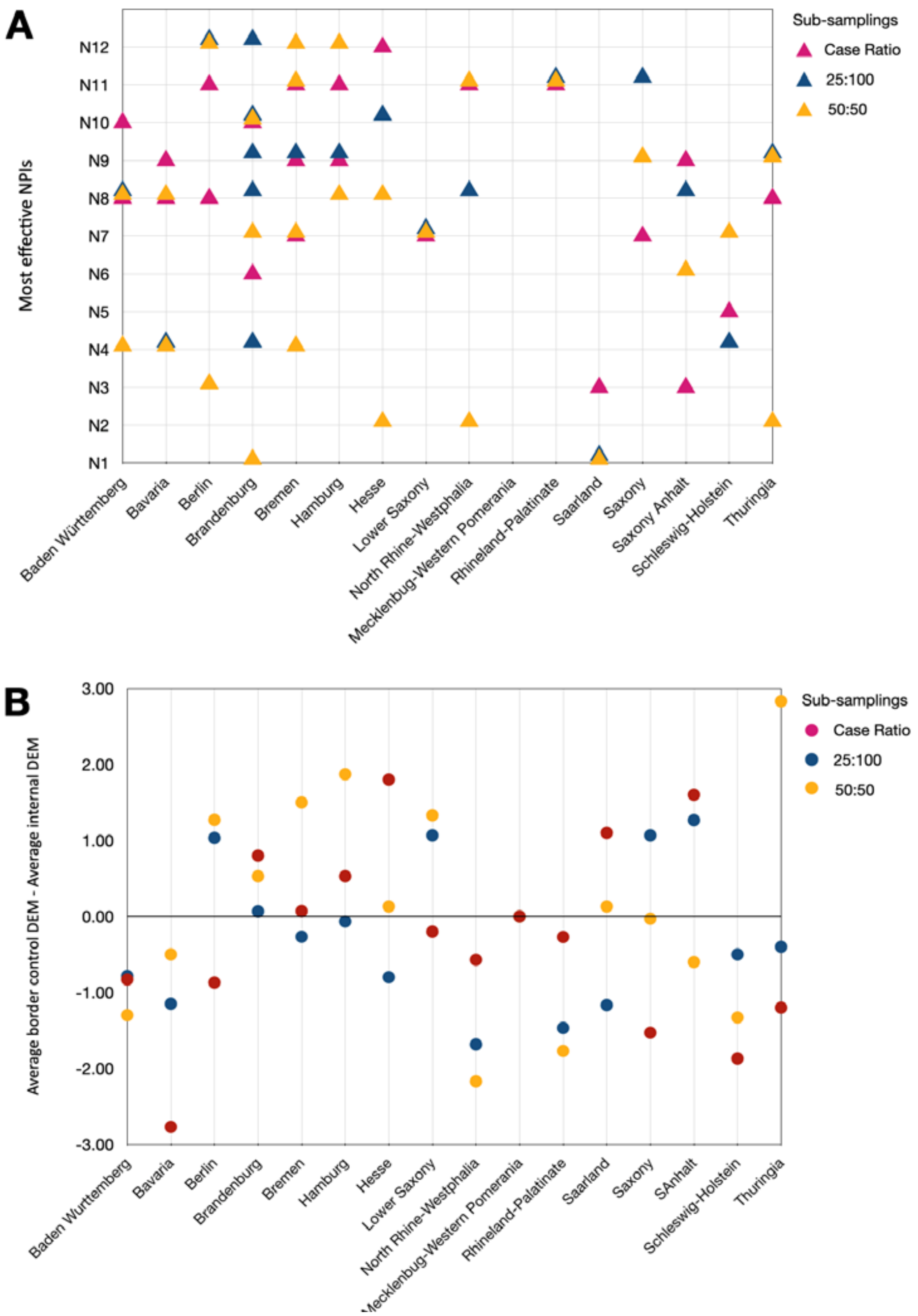


**Fig. S8 - State-wise effectiveness of NPIs. (A)** Maximum effective NPI in states. A state can have multiple maximum effective NPIs, all with equal effectiveness. **(B)** State-wise difference between average border control NPIs DEM and average internal NPIs DEM (positive values indicate border control NPI were more effective than internal NPIs in the state).

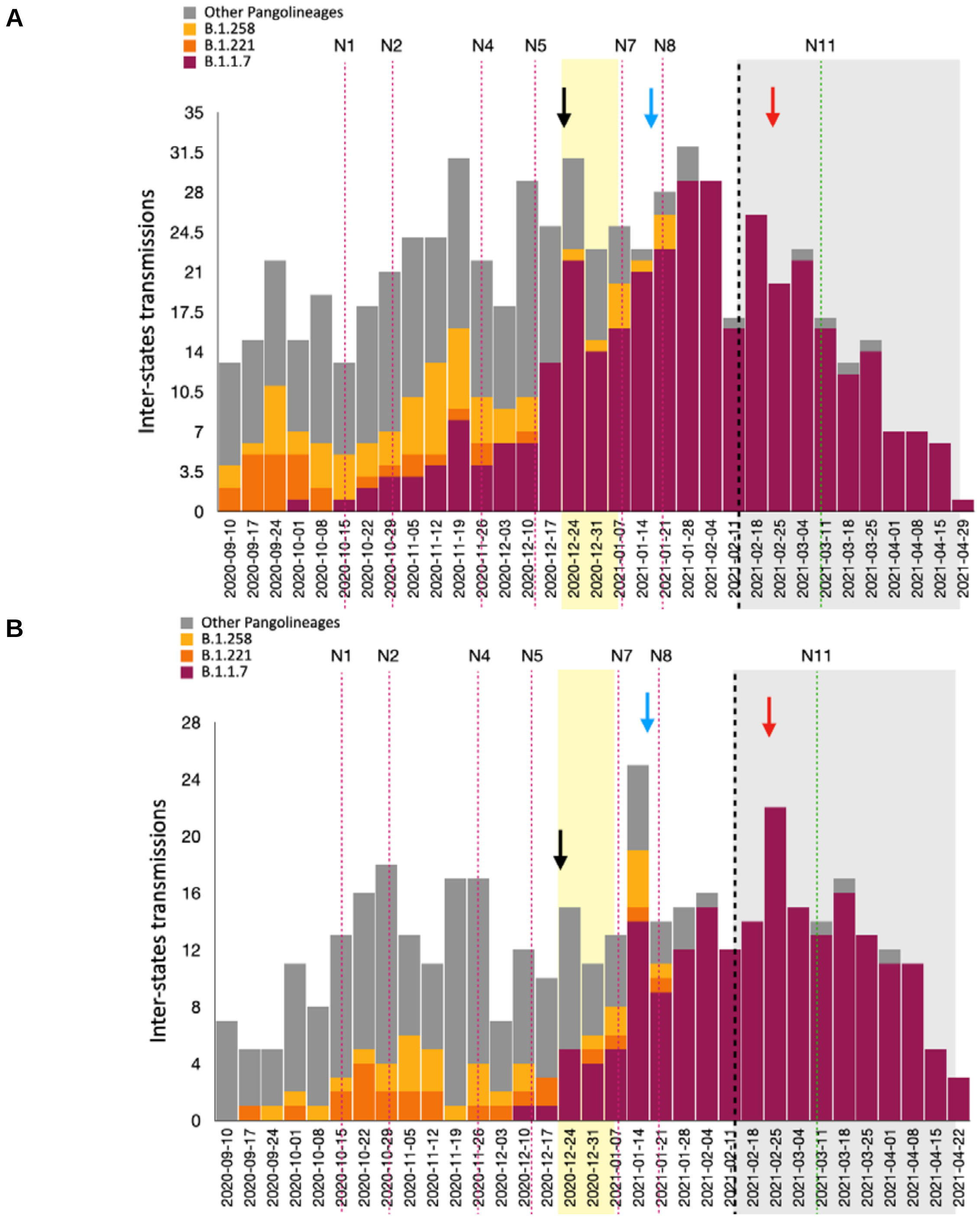


**Fig. S9 - Internal spread of lineages by Pango-lineage. (A)** 50:50 Subsampling **(B)** 25:100 Subsampling.

**Table S4 - Case ratio: Internal spread of lineages.** Number of lineage movements between states (row as source, column as destination) inferred with a Bayesian DTA model. German state abbreviations are **BW**: Baden-Württemberg, **BY**: Bavaria, **BE**: Berlin, **BB**: Brandenburg, **HB**: Bremen, **HH**: Hamburg, **HE**: Hesse, **NI**: Lower Saxony, **MV**: Mecklenburg-Vorpommern, **NW**: North Rhine-Westphalia, **RP**: Rhineland-Palatinate, **SL**: Saarland, **SN**: Saxony, **ST**: Saxony-Anhalt, **SH**: Schleswig-Holstein, TH: Thuringia. The red-highlighted cells represent the top three cells with the highest number of inter-state lineage movements.

Internal spread of lineages in case ratio subsampling

| | BW | BY | BE | BB | HB | HH | HE | NI | MP | NW | RP | SL | SN | ST | SH | TH |
|---|---|---|---|---|---|---|---|---|---|---|---|---|---|---|---|---|
| BW | 28 | 0 | 0 | 1 | 0 | 0 | 0 | 0 | 0 | 0 | 0 | 0 | 0 | 1 | 0 | 0 |
| BY | 8 | 113 | 0 | 0 | 0 | 1 | 2 | 0 | 0 | 1 | 0 | 1 | 4 | 3 | 1 | 2 |
| BE | 1 | 0 | 8 | 0 | 0 | 0 | 0 | 0 | 0 | 0 | 0 | 1 | 0 | 0 | 0 | 0 |
| BB | 0 | 0 | 1 | 1 | 0 | 0 | 0 | 0 | 0 | 0 | 0 | 0 | 0 | 0 | 0 | 0 |
| HB | 0 | 0 | 0 | 0 | 5 | 0 | 0 | 0 | 0 | 1 | 0 | 0 | 0 | 0 | 0 | 0 |
| HH | 0 | 0 | 1 | 0 | 0 | 127 | 0 | 0 | 0 | 0 | 1 | 0 | 1 | 1 | 1 | 0 |
| HE | 0 | 0 | 0 | 0 | 0 | 0 | 6 | 1 | 1 | 0 | 0 | 0 | 1 | 0 | 0 | 1 |
| NI | 0 | 0 | 0 | 0 | 0 | 0 | 0 | 8 | 0 | 0 | 0 | 0 | 0 | 0 | 0 | 0 |
| NW | 10 | 5 | 2 | 0 | 2 | 2 | 3 | 7 | 0 | 292 | 2 | 4 | 9 | 1 | 5 | 4 |
| RP | 0 | 0 | 0 | 0 | 0 | 0 | 0 | 0 | 0 | 0 | 9 | 1 | 0 | 0 | 0 | 0 |
| SL | 2 | 0 | 0 | 0 | 0 | 0 | 0 | 0 | 0 | 1 | 0 | 65 | 0 | 0 | 1 | 1 |
| SN | 2 | 0 | 0 | 0 | 0 | 0 | 5 | 0 | 0 | 2 | 0 | 3 | 46 | 3 | 0 | 3 |
| ST | 0 | 0 | 1 | 0 | 0 | 1 | 0 | 2 | 0 | 1 | 0 | 0 | 0 | 31 | 1 | 1 |
| SH | 0 | 0 | 1 | 0 | 0 | 0 | 0 | 0 | 0 | 0 | 0 | 0 | 0 | 0 | 21 | 0 |
| TH | 2 | 0 | 0 | 0 | 0 | 0 | 0 | 0 | 0 | 0 | 2 | 0 | 0 | 1 | 0 | 11 |

**Table S4 (continue) - Internal spread of lineages.**

| Internal spread of lineages in 50:50 subsampling | | | | | | | | | | | | | | | | |
|---|---|---|---|---|---|---|---|---|---|---|---|---|---|---|---|---|
| | BW | BY | BE | BB | HB | HH | HE | NI | MV | NW | RP | SL | SN | ST | SH | TH |
| BW | 212 | 14 | 1 | 1 | 3 | 0 | 1 | 0 | 0 | 23 | 1 | 0 | 3 | 0 | 0 | 1 |
| BY | 12 | 387 | 4 | 0 | 0 | 2 | 0 | 3 | 3 | 9 | 7 | 2 | 12 | 5 | 7 | 9 |
| BE | 4 | 3 | 139 | 11 | 1 | 2 | 1 | 1 | 2 | 4 | 1 | 6 | 11 | 4 | 3 | 3 |
| BB | 0 | 0 | 0 | 9 | 0 | 0 | 0 | 0 | 0 | 0 | 0 | 0 | 1 | 0 | 0 | 0 |
| HB | 0 | 0 | 0 | 0 | 11 | 0 | 1 | 0 | 0 | 0 | 0 | 0 | 0 | 0 | 0 | 0 |
| HH | 0 | 2 | 0 | 0 | 2 | 186 | 0 | 0 | 0 | 1 | 2 | 1 | 3 | 1 | 2 | 0 |
| HE | 3 | 3 | 0 | 0 | 0 | 1 | 42 | 1 | 0 | 3 | 0 | 1 | 0 | 0 | 2 | 0 |
| NI | 2 | 0 | 0 | 0 | 2 | 1 | 0 | 52 | 0 | 2 | 0 | 0 | 0 | 1 | 0 | 0 |
| NW | 63 | 59 | 25 | 5 | 10 | 23 | 19 | 29 | 4 | 1274 | 26 | 29 | 53 | 27 | 28 | 20 |
| RP | 5 | 1 | 0 | 0 | 0 | 0 | 0 | 0 | 1 | 1 | 43 | 0 | 0 | 0 | 0 | 1 |
| SL | 2 | 0 | 0 | 0 | 0 | 0 | 0 | 1 | 0 | 7 | 1 | 223 | 0 | 0 | 0 | 0 |
| SN | 9 | 6 | 2 | 1 | 1 | 0 | 9 | 1 | 1 | 5 | 4 | 3 | 151 | 2 | 3 | 0 |
| ST | 3 | 1 | 0 | 1 | 0 | 0 | 0 | 3 | 2 | 1 | 0 | 0 | 2 | 83 | 0 | 2 |
| SH | 3 | 4 | 0 | 0 | 0 | 4 | 0 | 2 | 0 | 1 | 0 | 1 | 0 | 0 | 109 | 0 |
| TH | 0 | 2 | 0 | 0 | 0 | 1 | 2 | 1 | 0 | 2 | 1 | 0 | 6 | 0 | 0 | 53 |

**Table S4 (continued) - Internal spread of lineages.**

| Internal spread of lineages in 25:100 subsampling | | | | | | | | | | | | | | | | |
|---|---|---|---|---|---|---|---|---|---|---|---|---|---|---|---|---|
| | BW | BY | BE | BB | HB | HH | HE | NI | MV | NW | RP | SL | SN | ST | SH | TH |
| BW | 153 | 5 | 0 | 1 | 0 | 1 | 2 | 0 | 1 | 5 | 3 | 0 | 2 | 0 | 1 | 2 |
| BY | 12 | 302 | 1 | 0 | 2 | 0 | 0 | 3 | 4 | 2 | 4 | 1 | 9 | 0 | 3 | 7 |
| BE | 4 | 2 | 132 | 9 | 1 | 0 | 0 | 2 | 2 | 7 | 1 | 6 | 9 | 4 | 3 | 2 |
| BB | 0 | 0 | 1 | 8 | 0 | 0 | 0 | 0 | 0 | 0 | 0 | 0 | 1 | 0 | 0 | 0 |
| HB | 0 | 0 | 0 | 0 | 9 | 0 | 1 | 0 | 0 | 0 | 0 | 0 | 0 | 0 | 0 | 0 |
| HH | 0 | 2 | 0 | 0 | 4 | 184 | 0 | 0 | 0 | 1 | 2 | 0 | 2 | 0 | 5 | 0 |
| HE | 3 | 3 | 0 | 0 | 0 | 0 | 23 | 0 | 0 | 1 | 0 | 2 | 0 | 0 | 1 | 1 |
| NI | 2 | 0 | 0 | 2 | 3 | 0 | 0 | 60 | 1 | 1 | 1 | 0 | 1 | 4 | 1 | 0 |
| NW | 31 | 25 | 9 | 1 | 5 | 7 | 8 | 15 | 1 | 773 | 10 | 8 | 20 | 7 | 11 | 5 |
| RP | 0 | 0 | 0 | 0 | 0 | 0 | 0 | 0 | 1 | 2 | 32 | 0 | 1 | 0 | 0 | 0 |
| SL | 3 | 0 | 0 | 0 | 0 | 0 | 0 | 1 | 0 | 6 | 0 | 224 | 0 | 0 | 2 | 0 |
| SN | 10 | 5 | 4 | 2 | 0 | 2 | 10 | 4 | 0 | 4 | 0 | 3 | 158 | 4 | 7 | 9 |
| ST | 4 | 0 | 0 | 1 | 0 | 0 | 0 | 1 | 0 | 1 | 0 | 0 | 2 | 70 | 0 | 1 |
| SH | 0 | 0 | 0 | 0 | 0 | 1 | 0 | 1 | 0 | 0 | 0 | 0 | 0 | 0 | 84 | 0 |
| TH | 2 | 0 | 0 | 0 | 0 | 0 | 0 | 2 | 0 | 6 | 4 | 0 | 0 | 1 | 0 | 51 |

| | Population | Number of incidence | Number of viral genomes | Sequencing rate |
|---|---|---|---|---|
| Population | | 0.98 | 0.86 | -0.31 |
| Number of incidence | 0.98 | | 0.9 | -0.25 |
| Number of viral genomes | 0.86 | 0.9 | | 0.05 |
| Sequencing rate | -0.31 | -0.25 | 0.05 | |

**Fig. S10** Correlation coefficient between measures for German states.

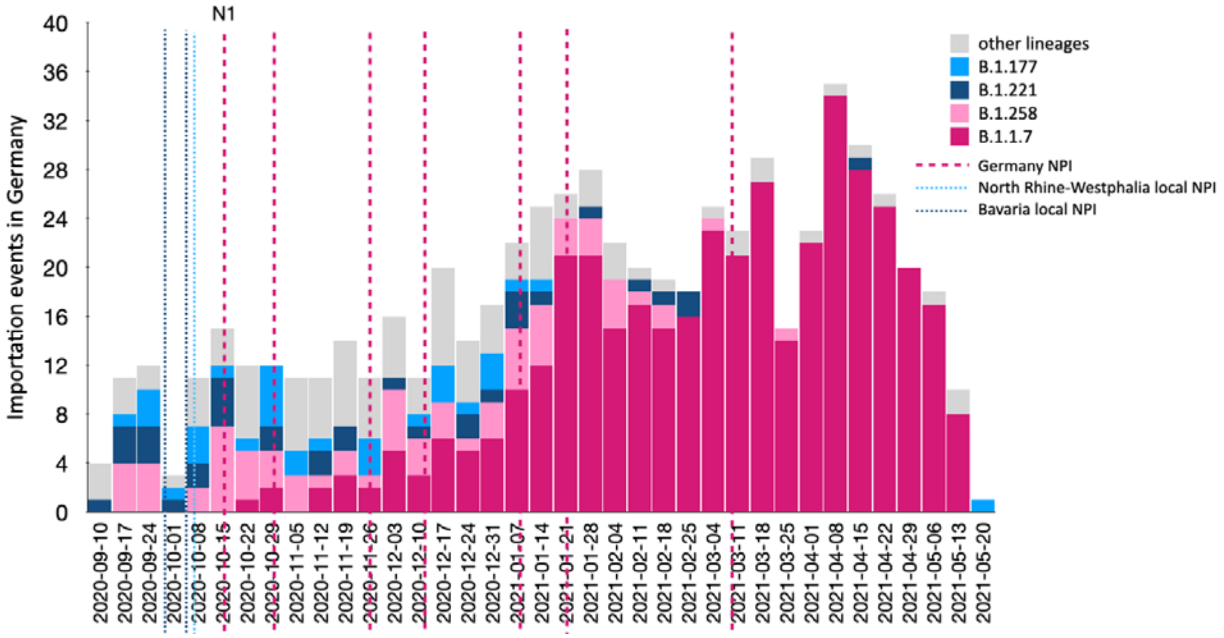


**Fig. S11 - Weekly number of importations colored by Pango-lineage.** Weekly number of importation events in Germany in the main subsampling, colored by the corresponding Pango-lineage (Dark pink for B.1.1.7, light pink for B.1.258, dark blue for B.1.221, light blue for B.1.77, gray for other Pango-lineages). Dotted lines represent the time of NPIs (dark pink for major NPIs in the country, light blue for local NPI in North Rhine-Westphalia, dark blue for local NPIs in Bavaria). The first major NPI (N1) was implemented before the arrival of the third wave and the introduction of B.1.1.7 into the country.

| **Table S1 - NPI Descriptions** Date and descriptions of non pharmaceutical interventions in Germany from 01-10-2020 until 01-06-2021 | | | | | | |
|---|---|---|---|---|---|---|
| | **Name** | **Date** | **Description** | **Labels** | **State** | **End Date** |
| 1 | | 01-10-2020 | *“"The extensive restrictions on entry at Germany’s external Schengen borders that Federal Minister Seehofer ordered on 17 March 2020 to contain the spread of the coronavirus were eased on 2 July. On 30 June 2020, the Council of the EU adopted the Council Recommendation on the temporary restriction on non-essential travel into the EU and the possible lifting of such restriction (Council document 2020/912). According to this recommendation, member states intend to lift, in a coordinated and gradual way, the temporary restriction of non-essential travel into the EU for residents of certain third countries (that is, those who have their domicile or habitual residence there). The list of these third countries is regularly reviewed and updated as necessary. Based on this recommendation, Germany lifted the restrictions on travel for residents of eight third countries as of 2 July. A first update removed two of these countries from the EU list of safe countries based on the Council Recommendation amending Recommendation (EU) 2020/912 of the Council on the temporary restriction on non-essential travel into the EU and the possible lifting of such restriction of 16 July 2020, which Germany implemented on 17 July 2020. Two more countries were classified as risk areas on 7 October 2020, which made it necessary to update the national list of safe countries. As of 27 October 2020, Germany allows unrestricted entry for residents of the following countries: Australia, New Zealand, Singapore, Thailand, Uruguay In addition, the list is to be expanded to include: Japan, South Korea, China, Hong Kong and Macao SARs of the People’s Republic of China as soon as the possibility of mutual entry is confirmed. For all persons residing in a third country that is not included in the above list, the current restrictions on travel continue to apply, i.e.* | *Border control (general policy)* | **Nationwide** | |

| Table S1 - NPI Descriptions Date and descriptions of non pharmaceutical interventions in Germany from 01-10-2020 until 01-06-2021 | | | | | | |
|---|---|---|---|---|---|---|
| | **Name** | **Date** | **Description** | **Labels** | **State** | **End Date** |
| | | | *these persons may enter Germany only if they serve in an important role or if there is an urgent need for their travel. A person is considered to reside in a country if they have their domicile or habitual residence there. A person is specifically considered to have their domicile in a country if they have spent the past six months there. The restrictions do not apply to German citizens. EU citizens and nationals of the United Kingdom, Liechtenstein, Switzerland, Norway and Iceland and the members of their immediate family (spouse, unmarried minor children, parents of minors) are exempted from the travel restrictions." Source: http://web.archive.org/web/20201130113325/https://www.bmi.bund.de/SharedDocs/faqs/EN/topics/civil-protection/coronavirus/travel-restrictions-border-control/travel-restriction-border-control-list.html (see "What restrictions apply to air and see travel outside of the EU)" [OXF, 276AC]* | | | |
| 2 | **A-** Bavaria | 02-10-2020 | *"From 2 October 2020, the Bavarian State Ministry for Health and Care in Germany decreed that* ***individuals are urged to reduce physical contact as much as possible, keep their social circle constant and maintain a minimum distance of 1,5 meters to others.*** *In public spaces (whenever minimum distance measures cannot be followed), public transport and touristic tour bus journeys, individuals are obliged to wear a face mask. Exceptions exist."*<br><br>*"From 2 October 2020, events and gatherings outside of religious services ("Gottesdienste"), gatherings of other public religious groups and public assemblies may be allowed if a minimum distance of 1,5 meters (2 meters in the case of singing and playing wind instruments) is maintained, the number of attendants does* | *Public events* | **Bavaria** | |

| Table S1 - NPI Descriptions Date and descriptions of non pharmaceutical interventions in Germany from 01-10-2020 until 01-06-2021 | | | | | | |
|---|---|---|---|---|---|---|
| | **Name** | **Date** | **Description** | **Labels** | **State** | **End Date** |
| | | | *not exceed 100 in confined spaces and 200 outdoors (if marked and assigned seats are available the number of attendants may comprise up to 200 in confined spaces and 400 outdoors), face covering measures in confined spaces are followed (except when at one's seat or rising to speak) and organizers have worked out a protection and hygiene concept and collected attendant's contact data. If in an administrative district or independent city ("kreisfreie Stadt") new SARS-CoV-2 infections surpass the number of 35 per 100.000 residents in the last 7 days, private events in public or rented space may only comprise up to 50 attendants and, in private spaces,* ***it is strongly recommended to limit the number of attendants to 25****.”* | | | |
| 3 | **A-<br>BY** | 02-10-2020 | *“From 2 October 2020, the Bavarian State Ministry for Health and Care in Germany decrees that all travellers from abroad who in the past 14 days have been to* ***"risk areas"*** *(based on information from the Robert Koch Institute) are obliged to* ***quarantine*** *themselves for* ***14 days upon arrival.*** *Exceptions exist. Risk areas have changed when compared to the regulatory situation of September 30. The policy is slated to end on October 18.”[DENPI, 3599G]* | *Border control (change of risk area)* | **Bavaria** | |
| 4 | **A -BY** | 02-10-2020 | *“From 30 September 2020, the Bavarian State Ministry for Health and Care in Germany decrees that all travellers from abroad who in the past 14 days have been to "risk areas" (based on information from the Robert Koch Institute) are obliged to quarantine themselves for 14 days upon* | *Border control* | **Bavaria** | |

| **Table S1 - NPI Descriptions** Date and descriptions of non pharmaceutical interventions in Germany from 01-10-2020 until 01-06-2021 | | | | | | |
|---|---|---|---|---|---|---|
| | **Name** | **Date** | **Description** | **Labels** | **State** | **End Date** |
| | | | *arrival. Exceptions exist. Risk areas have changed when compared to the regulatory situation of September 25. The policy is slated to end on October 3. UPDATE: From 2 October 2020, risk areas have changed in the following way:*<br><br>*"Netherlands: the complete country with exception of the provinces/constituent countries Zeeland and Limburg is considered as risk area.*<br><br>*United Kingdom of Great Britain and Northern Ireland: Scotland and the nations in England North West, North East, Yorkshire and the Humber are considered as risk areas.*<br><br>*In Austria the municipalities Mittelberg / Kleinwalsertal (Vorarlberg) and Jungholz (Tirol) are no longer considered risk areas."*<br><br>*Also, quarantine measures are extended to October 18." [DENPI, 3601G]* | *(change of risk area)* | | |
| 5 | **A-BW** | 02-10-2020 | *"From 2 October, the Baden-Wuerttemberg Ministry of Social Affairs in Germany decrees that all travelers from abroad (including individuals who necessarily have to enter BW for educational reasons) who in the past 14 days have been to "risk areas" are obliged to quarantine themselves for 14 days upon arrival." [DENPI, 3598G]* | *Border control* | **Baden Württemberg** | |
| 6 | **A-BW** | 02-10-2020 | *From 30 September, the Baden-Wuerttemberg Ministry of Social Affairs in Germany decrees that all travellers from abroad (including individuals who necessarily have to enter BW for educational reasons) who in the past 14 days have been to "risk areas" are obliged to quarantine themselves for 14 days upon arrival. Exceptions exist. Risk areas have* | *Border control* | **Baden Württemberg** | |

| Table S1 - NPI Descriptions Date and descriptions of non pharmaceutical interventions in Germany from 01-10-2020 until 01-06-2021 | | | | | | |
|---|---|---|---|---|---|---|
| | **Name** | **Date** | **Description** | **Labels** | **State** | **End Date** |
| | | | *changed when compared to the regulatory situation of September 25. The end of this policy goes along with the end of "CoronaVO" from June 23. It is slated to end on November 30. UPDATE: From 2 October, risk areas have changed in the following way:*<br><br>*"Netherlands: the complete country with exception of the provinces/constituent countries Zeeland and Limburg is considered as risk area.*<br><br>*United Kingdom of Great Britain and Northern Ireland: Scotland and the nations in England North West, North East, Yorkshire and the Humber are considered as risk areas.*<br><br>*In Austria the municipalities Mittelberg / Kleinwalsertal (Vorarlberg) and Jungholz (Tirol) are no longer considered risk areas."[DENPI, 3600G]* | | | |
| 7 | | 05-10-2020 | *"As of October 05, 2020, anyone entering Saarland (Germany) from a risk area, except returning travelers who spent less than 24 hours in either Luxembourg or France, has to quarantine for 14 days and take a mandatory test upon arrival. Previously announced quarantine policies (ordinance from 08-24-2020) regarding asylum seekers, enforcer, and exceptions regarding commuters remain in place as well."[DENPI, 3629G]*<br><br>*"In Saarland (Germany), as of August 24, 2020, anyone entering from a risk area has to quarantine for 14 days. Additionally, anyone who is newly arriving at a federal arrival centre for asylum seekers in Saarland (Germany), or anyone who is returning there after being absent for several days is not allowed to leave the assigned accommodation and utilities sector for 14 days. Commuters to Luxembourg and anyone with valid reasons* | *Border control* | Saarland | |

| **Table S1 - NPI Descriptions** Date and descriptions of non pharmaceutical interventions in Germany from 01-10-2020 until 01-06-2021 | | | | | | |
|---|---|---|---|---|---|---|
| | **Name** | **Date** | **Description** | **Labels** | **State** | **End Date** |
| | | | *for travel are exempt from the mandatory quarantine of 14 days. Exceptions also apply to anyone whose work is necessary to maintain diplomatic relations, to people who spent less than 72 hours in Luxembourg and to anyone providing a negative test from within 48 hours. UPDATE: On October 02, 2020 the government in Saarland (Germany) announced that it has decided to make additional exceptions to the quarantine policy for people who travel back and forth to Saarland for professional and private reasons from Luxembourg and France. As of October 05, anyone spending less than 24 hours in either Luxembourg or France does not have to quarantine after entering Saarland." [DENPI, 3630G]* | | | |
| 8 | | 07-10-2020 | *"From 7 October 2020, the Bavarian State Ministry for Health and Care in Germany decrees that all travellers from abroad who in the past 14 days have been to "risk areas" (based on information from the Robert Koch Institute) are obliged to quarantine themselves for 14 days upon arrival. Exceptions exist. Risk areas have changed when compared to the regulatory situation of October 2. The policy is slated to end on October 18." [DENPI, 3634G]*<br><br>*From 2 October 2020, the Bavarian State Ministry for Health and Care in Germany decrees that all travellers from abroad who in the past 14 days have been to "risk areas" (based on information from the Robert Koch Institute) are obliged to quarantine themselves for 14 days upon arrival. Exceptions exist. Risk areas have changed when compared to the regulatory situation of September 30. The policy is slated to end on October 18. UPDATE: From 7 October 2020, risk areas have changed in the following way:* | *Border control*<br><br>*(change of risk area)* | **Bavaria** | |

| **Table S1 - NPI Descriptions** Date and descriptions of non pharmaceutical interventions in Germany from 01-10-2020 until 01-06-2021 | | | | | | |
|---|---|---|---|---|---|---|
| | **Name** | **Date** | **Description** | **Labels** | **State** | **End Date** |
| | | | *"Bulgaria: the province Targovishte is considered as a risk area.*<br><br>*Croatia: the counties Vukovarsko-srijemska, Sisačko-moslavačka, Krapinsko-zagorska županija are considered as risk areas.*<br><br>*Hungary: the counties Nógrád, Baranya, Hajdú-Bihar, Jász-Nagykun-Szolnok, BorsodAbaúj-Zemplén, Komárom-Esztergom and Szabolcs-Szatmár-Bereg are considered as risk areas.*<br><br>*Lithuania: the county Kaunas is considered as a risk area.*<br><br>*Netherlands: the complete country with exception of the province Zeeland and the constituent country Curacao are considered as risk areas.*<br><br>*Romania: the complete country is considered as a risk area.*<br><br>*Slovakia: the regions/Kraj Zilina, Prešov, Bratislava, Nitra and Trnava are considered as risk areas.*<br><br>*Slovenia: the regions Zasavska, Gorenjska, Osrednjeslovenska, and Savinjska are considered as risk areas.*<br><br>*Tunisia: the complete country is considered as a risk area.*<br><br>*Georgia: the complete country is considered as a risk area.*<br><br>*Jordan: the complete country is considered as a risk area.*<br><br>*In Croatia the county Brodsko-Posavska and in France the region/ island Corsica is no longer considered as a risk area." [DENPI, 3637G]* | | | |

**Table S1 - NPI Descriptions** Date and descriptions of non pharmaceutical interventions in Germany from 01-10-2020 until 01-06-2021

| | Name | Date | Description | Labels | State | End Date |
|---|---|---|---|---|---|---|
| 9 | **B-BW** | 07-10-2020 | *“From 7 October, the* Baden-Wuerttemberg *Ministry of Social Affairs in Germany decrees that all travellers from abroad (including individuals who necessarily have to enter BW for educational reasons) who in the past 14 days have been to "risk areas" are obliged to quarantine themselves for 14 days upon arrival. Exceptions exist. Risk areas have changed when compared to the regulatory situation of October 2. The end of this policy goes along with the end of "CoronaVO" from June 23. It is slated to end on November 30.” [DENPI, 3633G]*<br><br>*“From 2 October, the Baden-Wuerttemberg Ministry of Social Affairs in Germany decrees that all travellers from abroad (including individuals who necessarily have to enter BW for educational reasons) who in the past 14 days have been to "risk areas" are obliged to quarantine themselves for 14 days upon arrival. Exceptions exist. Risk areas have changed when compared to the regulatory situation of September 30. The end of this policy goes along with the end of "CoronaVO" from June 23. It is slated to end on November 30. UPDATE: From 7 October, risk areas have changed in the following way:*<br><br>*"Bulgaria: the province Targovishte is considered as a risk area.*<br><br>*Croatia: the counties Vukovarsko-srijemska, Sisačko-moslavačka, Krapinsko-zagorska županija are considered as risk areas.*<br><br>*Hungary: the counties Nógrád, Baranya, Hajdú-Bihar, Jász-Nagykun-Szolnok, BorsodAbaúj-Zemplén, Komárom-Esztergom and Szabolcs-Szatmár-Bereg are considered as risk areas.* | *Border control* | **Baden Württemberg** | |

| Table S1 - NPI Descriptions Date and descriptions of non pharmaceutical interventions in Germany from 01-10-2020 until 01-06-2021 | | | | | | |
|---|---|---|---|---|---|---|
| | **Name** | **Date** | **Description** | **Labels** | **State** | **End Date** |
| | | | *Lithuania: the county Kaunas is considered as a risk area.*<br><br>*Netherlands: the complete country with exception of the province Zeeland and the constituent country Curacao are considered as risk areas.*<br><br>*Romania: the complete country is considered as a risk area.*<br><br>*Slovakia: the regions/Kraj Zilina, Prešov, Bratislava, Nitra and Trnava are considered as risk areas.*<br><br>*Slovenia: the regions Zasavska, Gorenjska, Osrednjeslovenska, and Savinjska are considered as risk areas.*<br><br>*Tunisia: the complete country is considered as a risk area.*<br><br>*Georgia: the complete country is considered as a risk area.*<br><br>*Jordan: the complete country is considered as a risk area.*<br><br>*In Croatia the county Brodsko-Posavska and in France the region/ island Corsica is no longer considered as a risk area."[DENPI, 3635G]* | | | |
| 10 | | 07-10-2020 | *“Bremen (Germany) is enforcing a 14 day quarantine for all persons who have been confirmed via laboratory diagnosis with coronavirus SARS-CoV-2. UPDATE: The original quarantine was set to last until May 3, it has been extended a fifteenth time to November 3.” [DENPI, 3636G]* | *Border Control* | Bremen | |
| 11 | **N1-a** | 14-10-2020 | *Due to the continuing high number of new infections, a curfew was introduced in North Rhine-Westphalia, Germany. This is intended to put a stop to new infections with* | | **North Rhine-Westphalia** | 02-11-2020 |

**Table S1 - NPI Descriptions** Date and descriptions of non pharmaceutical interventions in Germany from 01-10-2020 until 01-06-2021

| | Name | Date | Description | Labels | State | End Date |
|---|---|---|---|---|---|---|
| | | | *the coronavirus in restaurants. In the capital, since October 10, pubs, bars and late-night bars have had to close between 11 p.m. and 6 a.m. Then no more alcohol may be served. In Hamm in NRW, bars and restaurants must close at 1 a.m. at the latest. [DENPI,3658G]* | | | |
| 12 | **N1-b** | 15-10-2020 | *"As of October 15, counties or regions that are above a certain alert level, have restrictions on* ***gatherings of fewer than 10 people (to be specific 5)****" [OXFNPI, 290R].*<br><br>*"On October 15, Germany introduced county-based policies. If regions/cities pass a certain alert level, increased restrictions will apply. This also concerns* ***stores*** *that closed due to store restrictions of* ***10m2 per person****" [OXFNPI, 290L].*<br><br>*In light of the new restrictions, the German Chancellor has advised German citizens to* ***stay at home*** *as much as possible.[OXFNPI, 290Y]* | *Gathering, Workplaces* | **Nationwide** | **02-11-2020 (more restricted)**<br><br>**-**<br><br>**05-01-2021 (more restricted)** |
| 13 | **N1-c** | 16-10-2020 | *Due to the corona pandemic, the North Rhine-Westphalian (Germany) state government is imposing extensive restrictions on the most populous state in Germany, effective from 16 October 2020. In addition, masks should be mandatory "in heavily frequented areas", for example in pedestrian zones.* | | **North Rhine-Westphalia** | |
| 14 | **N1-d** | 17-10-2020 | *"Starting 17 October 2020, in NRW, Germany are enforcing new measures for a cumulative incidence above 50. Catering facilities must be completely* ***closed between 23.00 and 6.00 hours (curfew)****. Then no out-of-home sales are allowed." [DENPI, 3693G]* | *Gathering, Workplaces, night curfew* | **North Rhine-Westphalia** | **01-11-2020 (more restricted)** |

| **Table S1 - NPI Descriptions** Date and descriptions of non pharmaceutical interventions in Germany from 01-10-2020 until 01-06-2021 | | | | | | |
|---|---|---|---|---|---|---|
| | **Name** | **Date** | **Description** | **Labels** | **State** | **End Date** |
| | | | *"Starting 17 October 2020, in NRW, Germany are enforcing new measures for a cumulative incidence above 50. Up to five people may meet in public space. More persons may only be present if they are close relatives (straight relatives, spouses, life partners and siblings) or if they **come from two households.**" [DENPI, 3707G]*<br><br>*"Starting 17 October 2020, in NRW, Germany are enforcing new measures for a cumulative incidence above 35. **Mass gatherings are restricted to 25 people, and are only allowed for special occasions** (e.g. wedding). For public events, the number of participants is limited to 1000. Starting at 300 people, a hygiene concept must be in place. Starting at 500, the organizer has to have a permission from local authorities. Participants have to wear face masks at all times." [DENPI ,3831G]* | | | |
| 15 | **N1-e** | 17-10-2020 | *"On Oct. 17 2020, the German Chancellor Angela Merkel urges the population to **stay at home**, whenever possible."* [DENPI,*3681G]* | *Internal movements* | **Nationwide** | **Just advice** |
| 16 | **N1-f** | 17-10-2020 | *"From 17 October 2020, there are no longer explicit rules for gastronomic businesses in areas with incidence rates above 50. However, in areas where incidence rates have surpassed the threshold of 35, **food and drinks may not be consumed within gastronomic businesses from 11 PM to 6 AM**." [DENPI, 3728]* | *Workplaces (night curfew)* | **Bavaria** | **This is a stricting a previous NPI** |
| 17 | **N1-g** | 17-10-2020 | *"From 17 October 2020, the Bavarian State Ministry for Health and Care in Germany decrees that all travelers from abroad who in the past 14 days have been to "risk areas" (based on information from the Robert Koch Institute) are obliged to* | *Border Control* | **Bavaria** | **08-11-2020**<br><br>**(less restricted)** |

| Table S1 - NPI Descriptions Date and descriptions of non pharmaceutical interventions in Germany from 01-10-2020 until 01-06-2021 | | | | | | |
|---|---|---|---|---|---|---|
| | **Name** | **Date** | **Description** | **Labels** | **State** | **End Date** |
| | | | *quarantine themselves for 14 days upon arrival. Exceptions exist. Risk areas have changed when compared to the regulatory situation of October 7. The policy is slated to end on November 8." [DENPI, 3686G]*<br><br>*(Self-Quarantine)*<br><br>*From 7 October 2020, the Bavarian State Ministry for Health and Care in Germany decrees that all travelers from abroad who in the past 14 days have been to "risk areas" (based on information from the Robert Koch Institute) are obliged to quarantine themselves for 14 days upon arrival. Exceptions exist. Risk areas have changed when compared to the regulatory situation of October 2. The policy is slated to end on October 18. UPDATE: From 17 October 2020, risk areas have changed in the following way:*<br><br>*"Croatia: the counties Grad (city) Zagreb und Međimurska are considered as additional risk areas.*<br><br>*Finland: the region Ostrobothnia is considered as risk area.*<br><br>*France: the complete Mainland France and the French Overseas Territory Martinique are considered as additional risk areas.*<br><br>*Hungary: the county Veszprém is considered as additional risk area.*<br><br>*Ireland: the regions Mid-West, South-West, Mid-East, West and Midlands are considered as additional risk areas.*<br><br>*Italy: the regions Campania and Liguria are considered as risk areas.*<br><br>*Malta: the complete country is considered a risk area.* | *(change of risk areas)* | | **20-12-2020 (end)**<br><br>**,**<br><br>**01-11-2020 (change of risk areas)** |

| **Table S1 - NPI Descriptions** Date and descriptions of non pharmaceutical interventions in Germany from 01-10-2020 until 01-06-2021 | | | | | | |
|---|---|---|---|---|---|---|
| | **Name** | **Date** | **Description** | **Labels** | **State** | **End Date** |
| | | | *Netherlands: the complete country (incl. constituent countries) is considered as a risk area.*<br><br>*Poland: the regions Kujawsko-Pomorskie, Małopolskie, Podlaski, Pomorskie and Świętokrzyskie are considered as risk areas.*<br><br>*Portugal: the region Norte is considered an additional risk area.*<br><br>*Slovakia: the complete country is considered as a risk area.*<br><br>*Slovenia: the regions Jugovzhodna Slovenija, Pomurska and Podravska are considered as additional risk areas.*<br><br>*Sweden: the provinces Jämtland, Örebro, Uppsala, Stockholm and are considered as risk areas.*<br><br>*Switzerland: the cantons Fribourg, Jura, Neuchâtel, Nidwalden, Schwyz, Uri, Zurich und Zug are considered as additional risk areas.*<br><br>*United Kingdom of Great Britain and Northern Ireland: England: the regions East Midlands and West Midlands are considered as additional risk areas.*<br><br>*Namibia is no longer considered as risk area."*<br><br>*Also, quarantine measures are extended to November 8." [DENPI, 3688G]* | | | |
| 18 | **N1-h** | 17-10-2020 | *"From 17 October, the Baden-Wuerttemberg Ministry of Social Affairs in Germany decrees that all travellers from abroad who in the past 14 days have been to "risk areas" are obliged to quarantine themselves for 14 days upon arrival. Exceptions include, among others, individuals who necessarily have to enter* | *Border Control* | **Baden Württembe rg** | **30-11-2020**<br><br>**(It was loosening itself. What happened then?)** |

| Table S1 - NPI Descriptions Date and descriptions of non pharmaceutical interventions in Germany from 01-10-2020 until 01-06-2021 | | | | | | |
|---|---|---|---|---|---|---|
| | **Name** | **Date** | **Description** | **Labels** | **State** | **End Date** |
| | | | *BW for educational reasons or from border regions who stay in BW for no longer than 24 hours. The end of this policy goes along with the end of "CoronaVO" from June 23. It is slated to end on November 30." [DENPI, 3684G]*<br><br>*"From 7 October, the Baden-Wuerttemberg Ministry of Social Affairs in Germany decrees that all travellers from abroad (including individuals who necessarily have to enter BW for educational reasons) who in the past 14 days have been to "risk areas" are obliged to quarantine themselves for 14 days upon arrival. Exceptions exist. Risk areas have changed when compared to the regulatory situation of October 2. The end of this policy goes along with the end of "CoronaVO" from June 23. It is slated to end on November 30. UPDATE: From 17 October, risk areas have changed in the following way:*<br><br>*"Croatia: the counties Grad (city) Zagreb und Međimurska are considered as additional risk areas.*<br><br>*Finland: the region Ostrobothnia is considered as risk area.*<br><br>*France: the complete Mainland France and the French Overseas Territory Martinique are considered as additional risk areas.*<br><br>*Hungary: the county Veszprém is considered as additional risk area.*<br><br>*Ireland: the regions Mid-West, South-West, Mid-East, West and Midlands are considered as additional risk areas.*<br><br>*Italy: the regions Campania and Liguria are considered as risk areas.* | | | **,**<br><br>**24-10-2020 (next change in risk areas)** |

| Table S1 - NPI Descriptions Date and descriptions of non pharmaceutical interventions in Germany from 01-10-2020 until 01-06-2021 | | | | | | |
|---|---|---|---|---|---|---|
| | **Name** | **Date** | **Description** | **Labels** | **State** | **End Date** |
| | | | *Malta: the complete country is considered as risk area.*<br><br>*Netherlands: the complete country (incl. constituent countries) is considered as risk area.*<br><br>*Poland: the regions Kujawsko-Pomorskie, Małopolskie, Podlaski, Pomorskie and Świętokrzyskie are considered as risk areas.*<br><br>*Portugal: the region Norte is considered additional as risk area.*<br><br>*Slovakia: the complete country is considered as risk area.*<br><br>*Slovenia: the regions Jugovzhodna Slovenija, Pomurska and Podravska are considered as additional risk areas.*<br><br>*Sweden: the provinces Jämtland, Örebro, Uppsala, Stockholm and are considered as risk areas.*<br><br>*Switzerland: the cantons Fribourg, Jura, Neuchâtel, Nidwalden, Schwyz, Uri, Zurich und Zug are considered as additional risk areas.*<br><br>*United Kingdom of Great Britain and Northern Ireland: England: the regions East Midlands and West Midlands are considered as additional risk areas.*<br><br>*Namibia is no longer considered as risk area."*<br><br>*Additionally, from 17 October, quarantine measures do no longer apply to individuals: Who necessarily have to enter BW for educational reasons; from border regions (the Austrian province of Vorarlberg; Liechtenstein; the Swiss cantons of Appenzell, Aargau, Basel, Basel-Landschaft, Jura, Schaffhausen, Solothurn, St. Gallen, Thurgau and Zurich; the French departments of Bas-Rhin and Haut-Rhin)* | | | |

| **Table S1 - NPI Descriptions** Date and descriptions of non pharmaceutical interventions in Germany from 01-10-2020 until 01-06-2021 | | | | | | |
|---|---|---|---|---|---|---|
| | **Name** | **Date** | **Description** | **Labels** | **State** | **End Date** |
| | | | *who stay in BW for no longer than 24 hours." [DENPI,3687G]* | | | |
| 19 | **N1-i** | 19-10-2020 | *"Germany has enacted a regulation that from October 19, 2020,* ***people with cold symptoms can have a medical certificate from a doctor for work via telephone*** *so that they do not have to enter the doctor's office and thus possibly infect doctors, practice staff or other patients with COVID-19." [DENPI,3736G]* | *Workplac es* | **Nationwide** | **31-12-2020** |
| 20 | **N1-j** | 19-10-2020 | ***"From 19 October, gatherings may now comprise only up to 10 individuals.*** *Exceptions are extended and now include up to 2 households. Private events may now comprise only up to 10, other events up to 100 attendants." [DENPI, 3756G]*<br><br>*"From 19 October,* ***face covering is now also mandatory in all pedestrian areas where keeping a minimum distance is not possible,*** *except for instances of sporting activity, and in public facility areas open for the public, except for a selective number of events (as stated elsewhere). Additionally, exceptions now explicitly include instances of food intake in gastronomic facilities." [DENPI, 3763]* | *Gatherin g*<br><br>*Masks* | **Baden Württembe rg** | **30-11-2020**<br><br>**(it was strengtheni ng)**<br><br>**,**<br><br>**30-11-2020**<br><br>**(I don't think the mask rule is gone)** |
| 21 | **B-BY** | 23-10-2020 | *"From 23 October 2020, if in an administrative district or independent city ("kreisfreie Stadt") new SARS-CoV-2 infections surpass the number of 100 per 100.000 residents in the last 7 days or have been surpassed less than 6 days ago,* ***events and assemblies not open for the public as well as sports events may comprise only up to 50 attendants or viewers, respectively.****"* | *Public events* | **Bavaria** | |

| **Table S1 - NPI Descriptions** Date and descriptions of non pharmaceutical interventions in Germany from 01-10-2020 until 01-06-2021 | | | | | | |
|---|---|---|---|---|---|---|
| | **Name** | **Date** | **Description** | **Labels** | **State** | **End Date** |
| 22 | **B-BY** | 23-10-2020 | *“From 17 October 2020, the Bavarian State Ministry for Health and Care in Germany decrees that all travellers from abroad who in the past 14 days have been to "risk areas" (based on information from the Robert Koch Institute) are obliged to quarantine themselves for 14 days upon arrival. Exceptions exist. Risk areas have changed when compared to the regulatory situation of October 7. The policy is slated to end on November 8. UPDATE: From 23 October 2020, having a health certificate to be exempted from quarantine measures is replaced by having a test result. Those from risk areas who have to enter Bavaria at least once a weak on a regular basis for occupational, commercial or educational reasons are required to provide a test result related to SARS-CoV-2 within 7 days of their first arrival in Bavaria starting from 23 October and, after that, once per week on a regular basis (provided that entry into Bavaria has taken place).” [DENPI, 3792G]* | *Border Control* | **Bavaria** | |
| **23** | | 24-10-2020 | *“From 24 October 2020, the Bavarian State Ministry for Health and Care in Germany decrees that all travelers from abroad who in the past 14 days have been to "risk areas" (based on information from the Robert Koch Institute) are obliged to quarantine themselves for 14 days upon arrival. Exceptions exist. Risk areas have changed when compared to the regulatory situation of October 23. The policy is slated to end on November 8.” [DENPI, 3811G]*<br><br>*“From 17 October 2020, the Bavarian State Ministry for Health and Care in Germany decrees that all travelers from abroad who in the past 14 days have been to "risk areas" (based on information from the Robert Koch Institute) are obliged to quarantine themselves for 14 days upon* | *Border Control (risk area)* | **Bavaria** | |

| | Name | Date | Description | Labels | State | End Date |
|---|---|---|---|---|---|---|
| | | | *arrival. Exceptions exist. Risk areas have changed when compared to the regulatory situation of October 7. The policy is slated to end on November 8. UPDATE: From 24 October 2020, risk areas have changed in the following way:*<br><br>*"Austria: the provinces Salzburg, Oberösterreich, Niederösterreich, Burgenland and Steiermark are considered as additional risk areas.*<br><br>*Bulgaria: the provinces Razgrad, Sofia City und Sliven are considered as additional risk areas.*<br><br>*Croatia: the counties Karlovac, Osijek-Baranja, Zagreb, Varaždin and Bjelovar-Bilogora are considered as additional risk areas.*<br><br>*Estonia: the region Jogeva is considered as risk area.*<br><br>*Hungary: the counties Heves, Zala and Somogy are considered as additional risk areas.*<br><br>*Ireland: the complete country is considered as risk area.*<br><br>*Italy: the regions Valle d'Aosta, Umbria, Lombardia, Piemonte, Toskana, Veneto, Lazio, Abrruzo, Friuli Venezia Giulia, Emilia-Romagna, Sardinien and the autonomous province Bolzano-South Tyrol are considered as additional risk areas.*<br><br>*Liechtenstein: the complete country is considered a risk area.*<br><br>*Poland: the complete country is considered a risk area.*<br><br>*Slovenia: the regions Posavska and Goriška are considered as additional risk areas.* | | | |

| **Table S1 - NPI Descriptions** Date and descriptions of non pharmaceutical interventions in Germany from 01-10-2020 until 01-06-2021 | | | | | | |
|---|---|---|---|---|---|---|
| | **Name** | **Date** | **Description** | **Labels** | **State** | **End Date** |
| | | | *Sweden: the provinces Jönköping and Östergötland are considered as additional risk areas.*<br><br>*Switzerland: the complete country is considered a risk area.*<br><br>*United Kingdom of Great Britain and Northern Ireland: the complete country United Kingdom of Great Britain and Northern Ireland and Gibraltar is considered as risk area. Excluded are further British Overseas Territories, Isle of Man and the Channel Islands (Guernsey, Jersey).*<br><br>*The Canary Islands in Spain and the region Ida-Viru in Estonia is no longer considered as risk areas." [DENPI,3813G]* | | | |
| 24 | | 24-10-2020 | *"From 24 October, the Baden-Wuerttemberg Ministry of Social Affairs in Germany decrees that all travellers from abroad who in the past 14 days have been to "risk areas" are obliged to quarantine themselves for 14 days upon arrival. Exceptions include, among others, individuals who necessarily have to enter BW for educational reasons or from border regions who stay in BW for no longer than 24 hours. Risk areas have changed when compared to the regulatory situation of October 17. The end of this policy goes along with the end of "CoronaVO" from June 23. It is slated to end on November 30." [DENPI, 3810G]*<br><br>*"From 17 October, the Baden-Wuerttemberg Ministry of Social Affairs in Germany decreed that all travelers from abroad who in the past 14 days have been to "risk areas" are obliged to quarantine themselves for 14 days upon arrival. Exceptions include, among others, individuals who necessarily have to enter BW for educational reasons or from border regions who stay in BW for no longer than* | *Border Control* | **Baden Württembe rg** | |

| Table S1 - NPI Descriptions Date and descriptions of non pharmaceutical interventions in Germany from 01-10-2020 until 01-06-2021 | | | | | | |
|---|---|---|---|---|---|---|
| | **Name** | **Date** | **Description** | **Labels** | **State** | **End Date** |
| | | | *24 hours. Risk areas have changed when compared to the regulatory situation of October 7. The end of this policy goes along with the end of "CoronaVO" from June 23. It is slated to end on November 30. UPDATE: From 24 October, risk areas have changed in the following way:*<br><br>*"Austria: the provinces Salzburg, Oberösterreich, Niederösterreich, Burgenland and Steiermark are considered as additional risk areas.*<br><br>*Bulgaria: the provinces Razgrad, Sofia City und Sliven are considered as additional risk areas.*<br><br>*Croatia: the counties Karlovac, Osijek-Baranja, Zagreb, Varaždin and Bjelovar-Bilogora are considered as additional risk areas.*<br><br>*Estonia: the region Jogeva is considered as risk area.*<br><br>*Hungary: the counties Heves, Zala and Somogy are considered as additional risk areas.*<br><br>*Ireland: the complete country is considered as risk area.*<br><br>*Italy: the regions Valle d'Aosta, Umbria, Lombardia, Piemonte, Toskana, Veneto, Lazio, Abrruzo, Friuli Venezia Giulia, Emilia-Romagna, Sardinien and the autonomous province Bolzano-South Tyrol are considered as additional risk areas.*<br><br>*Liechtenstein: the complete country is considered as risk area.*<br><br>*Poland: the complete country is considered as risk area.*<br><br>*Slovenia: the regions Posavska and Goriška are considered as additional risk areas.* | | | |

| Table S1 - NPI Descriptions Date and descriptions of non pharmaceutical interventions in Germany from 01-10-2020 until 01-06-2021 | | | | | | |
|---|---|---|---|---|---|---|
| | **Name** | **Date** | **Description** | **Labels** | **State** | **End Date** |
| | | | *Sweden: the provinces Jönköping and Östergötland are considered as additional risk areas.*<br><br>*Switzerland: the complete country is considered as risk area.*<br><br>*United Kingdom of Great Britain and Northern Ireland: the complete country United Kingdom of Great Britain and Northern Ireland and Gibraltar is considered as risk area. Excluded are further British Overseas Territories, Isle of Man and the Channel Islands (Guernsey, Jersey).*<br><br>*The Canary Islands in Spain and the region Ida-Viru in Estonia is no longer considered as risk areas." [DENPI,3812G]* | | | |
| 25 | **N2-a** | 01-11-2020 | *"From 1 November 2020, the Bavarian State Ministry for Health and Care in Germany decrees that all travellers from abroad who in the past 14 days have been to "risk areas" (based on information from the Robert Koch Institute) are obliged to quarantine themselves for 14 days upon arrival. Exceptions exist. Risk areas have changed when compared to the regulatory situation of October 24. The policy is slated to end on November 8." [DENPI, 3829G]*<br><br>*"From 24 October 2020, the Bavarian State Ministry for Health and Care in Germany decrees that all travellers from abroad who in the past 14 days have been to "risk areas" (based on information from the Robert Koch Institute) are obliged to quarantine themselves for 14 days upon arrival. Exceptions exist. Risk areas have changed when compared to the regulatory situation of October 23. The policy is slated to end on November 8. UPDATE: From 1* | *Border Control*<br><br>*(Change in risk area)* | **Bavaria** | **08-11-2020**<br><br>**,**<br><br>**08-11-2020 (change in risk areas)** |

| Table S1 - NPI Descriptions Date and descriptions of non pharmaceutical interventions in Germany from 01-10-2020 until 01-06-2021 | | | | | | |
|---|---|---|---|---|---|---|
| | **Name** | **Date** | **Description** | **Labels** | **State** | **End Date** |
| | | | *November 2020, risk areas have changed in the following way:*<br><br>*"Austria: the complete country is considered as risk area with exception of the municipality Jungholz and Mittelberg/ Kleinwalsertal.*<br><br>*Bulgaria: the complete country is considered as risk area.*<br><br>*Croatia: the complete country is considered as risk area.*<br><br>*Cyprus: Cyprus is considered as risk area.*<br><br>*Denmark: the region Nordjylland is considered as additional risk area.*<br><br>*Greece: the region Western Macedonia is considered as additional risk area.*<br><br>*Hungary: the complete country is considered as risk area.*<br><br>*Italy: the complete country is considered as risk area with exception of the region of Calabria.*<br><br>*Latvia: the regions Latgale, Riga and Vidzeme are considered as additional risk areas.*<br><br>*Lithuania: the counties Klaipéda, Marijanpolé, Telšiai and Vilnius are considered as additional risk areas.*<br><br>*Monaco: the complete country is considered as risk area.*<br><br>*Portugal: the region Centro is considered as additional risk area.*<br><br>*San Marino: the complete country is considered as risk area.*<br><br>*Slovenia: the complete country is considered as risk area.* | | | |

| Table S1 - NPI Descriptions Date and descriptions of non pharmaceutical interventions in Germany from 01-10-2020 until 01-06-2021 | | | | | | |
|---|---|---|---|---|---|---|
| | **Name** | **Date** | **Description** | **Labels** | **State** | **End Date** |
| | | | *Sweden: the provinces Dalarna, Halland, Kronoberg, Skåne, Västmanland and Västra Götaland are considered as additional risk areas.*<br><br>*Vatican City State: the complete country is considered as risk area.*<br><br>*The region Jogeva in Estonia is no longer considered as risk area." [DENPI, 3830G]* | | | |
| 26 | **N2-b** | 02-11-2020 | *"**Public events** have to be canceled." [OXFNPI, 308O]*<br><br>*"Most **workplaces** will have to close, except for grocery stores and shopping stores." [OXFNPI, 308L]*<br><br>*"Restrictions are now in place across the country, limiting the max of **people meeting to be 2 households**." [OXFNPI, 308R]*<br><br>*"The German government STRONGLY advises everyone to **not travel within Germany**. It may be changed to a 2, since you cannot travel within Germany for tourist reasons but there are no explicit prohibitions for internal movement as of now. The German government urges everyone to reduce contact as much as possible." [OXFNPI, 308AA]*<br><br>*"Germany is enforcing a **partial lockdown targeting restaurants, theaters, operas, museums, concert halls, fitness studios, beauty salons, leisure and sports facilities**. All these facilities are forced to close from November 2, 2020 for four weeks. Amateur sports operations will be discontinued, so clubs will no longer be allowed to train." [DENPI, 3850G]* | *Public events, Workplaces, Gathering, Internal movements* | **Nationwide** | **10-01-2021**<br><br>**,**<br><br>**30-11-2020**<br><br>**More restricted,**<br><br>**30-11-2020**<br><br>**More restricted**<br><br>**,** |

| **Table S1 - NPI Descriptions** Date and descriptions of non pharmaceutical interventions in Germany from 01-10-2020 until 01-06-2021 | | | | | | |
|---|---|---|---|---|---|---|
| | **Name** | **Date** | **Description** | **Labels** | **State** | **End Date** |
| 27 | **N2-c** | 02-11-2020 | *"From 2 November, the Baden-Wuerttemberg State Government in Germany decrees that, in BW, facilities within the **catering industry are to be closed for the public.** Catering facilities and services are exempted." [DENPI, 3853G]*<br><br>*"From 2 November, the Baden-Wuerttemberg State Government in Germany decreed that, in BW, **amusement parks** are to be closed for the public." [DENPI, 3857G]*<br><br>*"From 2 November, infection protection principles are stepped up: Retailing operations and markets according to §§ 66 to 68 of the Gewerbeordnung (Trade Regulation) have to make sure that, within closed space, t**he number of customers per 10 m² of selling space does not exceed 1** at the same time. In facilities with less than 10 m² of selling space, a maximum number of 1 customer is allowed at the same time." [DENPI, 3879G]* | *Workplaces,* | **Baden-Württemberg** | **30-11-2020**<br><br>**More restricted** |
| 28 | **N2-d** | 02-11-2020 | *"Saxony (Germany) institutes a **closure of diverse businesses**. From swimming pools, sauna and gyms to casinos, betting facilities, leisure parks, botanical gardens, christmas markets, fairs, clubs, restaurants, bars, prostitution facilities." [DENPI, 3859G]*<br><br>*"From November 2, Saxony (Germany) closes education and training institutions that do not serve the purpose of vocational, school or academic training." [DENPI, 3865G]*<br><br>*"Saxony (Germany) allows **gatherings with up to 10 people from two households** in public space." [DENPI, 3901G]* | *Workplaces, Gathering* | Saxony | 07-02-2021 |

| Table S1 - NPI Descriptions Date and descriptions of non pharmaceutical interventions in Germany from 01-10-2020 until 01-06-2021 | | | | | | |
|---|---|---|---|---|---|---|
| | **Name** | **Date** | **Description** | **Labels** | **State** | **End Date** |
| 29 | | 02-11-2020 | *“Persons entering* Saxony *(Germany) from abroad who have been in a foreign risk area (as published by the Robert Koch Institute) at any time in the ten days prior to entry must immediately remain permanently in their home for a period of ten days (previously 14 days) after entry: Domestic quarantine may be terminated no earlier than five days after entry by a negative Corona test. The test must have been taken no earlier than five days after entry.” [DENPI, 3843G]* | *Border Control* | Saxony | |
| 30 | | 03-11-2020 | *“Berlin (Germany) introduces a new quarantine policy from November 3. Now quarantine for returning travelers from risk areas can be shorted from 10 days to 5 days through a corona test on the 5th day of quarantine.”[DENPI,3945G]* | *Border Control* | Berlin | |
| 31 | | 03-11-2020 | *“Bremen (Germany) enforced a 14-day quarantine for persons who enter Bremen by land, sea or air from a risk area where they have stayed for 10 days. This does not pertain to person who spend less than 72 hours abroad, commuters (with residence in Bremen or abroad), system-relevant workers, vacation returnees who were tested negatively for SARS-CoV-2 no more than 48 hours before entry into the Federal Republic of Germany or immediately upon entry.” [DENPI, 3943G]*<br><br>*“Bremen (Germany) is enforcing an exception from the 14-day quarantine for people returning or traveling to Bremen from abroad. The quarantine can be shorted to 5 days if after entry the person has a negative corona test result on paper or in an electronic document in German, English or French and immediately submits it to health authorities within ten days after the entry. The underlying test must have been performed at least five days after entry* | *Border Control* | Bremen | |

| Table S1 - NPI Descriptions Date and descriptions of non pharmaceutical interventions in Germany from 01-10-2020 until 01-06-2021 | | | | | | |
|---|---|---|---|---|---|---|
| | **Name** | **Date** | **Description** | **Labels** | **State** | **End Date** |
| | | | *into the Federal Republic of Germany and must be kept for at least ten days after entry." [DENPI, 3944G]*<br><br>*"Bremen (Germany) is enforcing a 14 day quarantine for all persons who have been confirmed via laboratory diagnosis with coronavirus SARS-CoV-2. UPDATE: The original quarantine was set to last until May 3, it has been extended a sixtheenth time to November 30." [DENPI, 3946G]* | | | |
| 32 | | 06-11-2020 | *"From 1 November 2020, the Bavarian State Ministry for Health and Care in Germany decrees that all travellers from abroad who in the past 14 days have been to "risk areas" (based on information from the Robert Koch Institute) are obliged to quarantine themselves for 14 days upon arrival. Exceptions exist. Risk areas have changed when compared to the regulatory situation of October 24. The policy is slated to end on November 8. UPDATE: From 6 November 2020, test results may also be provided through Corona rapid tests." [DENPI, 3974G]*<br><br>*From 8 November 2020, the Bavarian State Ministry for Health and Care in Germany decrees that all travellers from abroad who in the past 14 days have been to "risk areas" (based on information from the Robert Koch Institute) are obliged to quarantine themselves for 14 days upon arrival. Exceptions exist. Risk areas have changed when compared to the regulatory situation of November 1. The policy is slated to end on November 8." [DENPI, 3976G]* | *Border Control* | **Bavaria** | |
| 33 | **N3-a** | 08-11-2020 | *"From 8 November 2020 all* ***travelers to Germany have to register online*** *prior to their entry on the website* | *Border control* | **Nationwide** | |

| Table S1 - NPI Descriptions Date and descriptions of non pharmaceutical interventions in Germany from 01-10-2020 until 01-06-2021 | | | | | | |
|---|---|---|---|---|---|---|
| | **Name** | **Date** | **Description** | **Labels** | **State** | **End Date** |
| | | | *www.einreiseanmeldung.de, if they have stayed in a risk area within the last 10 days prior to their entry to Germany. Also they must be able to present proof of this registration when entering Germany." Previous restrictions remain in place." [OXF, 314AC]* | | | |
| 34 | | 08-11-2020 | *"From 8 November 2020 all travelers to Germany have to register online prior to their entry on the website www.einreiseanmeldung.de , if they have stayed in a risk area within the last 10 days prior to their entry to Germany. Also they must be able to present proof of this registration when entering Germany." Previous restrictions remain in place. Source: http://web.archive.org/web/20201129121721/https://www.auswaertiges-amt.de/en/coronavirus/2317268 [OXF, 314AC]* | *Border Control* | **Nationwide** | |
| **35** | **N3-b** | 08-11-2020 | *"New quarantine rules will come into force this weekend for returnees from risk areas abroad. They will then only have to isolate themselves for 10 days instead of the previous 14 days. However, they can also be "freed" after five days. Until a negative test result is obtained, returnees will have to stay at home. The bottom line is that this should add up to about a week, because a test can take 48 hours or even longer." [deCov1]* | *Border Control* | **Nationwide** | **09-05-2021** |
| 36 | | 08-11-2020 | *"From 1 November 2020, the Bavarian State Ministry for Health and Care in Germany decrees that all travellers from abroad who in the past 14 days have been to "risk areas" (based on information from the Robert Koch Institute) are obliged to* | *Border Control* | **Bavaria** | |

| Table S1 - NPI Descriptions Date and descriptions of non pharmaceutical interventions in Germany from 01-10-2020 until 01-06-2021 | | | | | | |
|---|---|---|---|---|---|---|
| | **Name** | **Date** | **Description** | **Labels** | **State** | **End Date** |
| | | | *quarantine themselves for 14 days upon arrival. Exceptions exist. Risk areas have changed when compared to the regulatory situation of October 24. The policy is slated to end on November 8. UPDATE: From 8 November 2020, risk areas have changed in the following way:*<br><br>*"Denmark: the complete country is considered as risk area with exception of the Faroe Islands*<br><br>*and Greenland.*<br><br>*Estonia: the region Ida-Viru is considered as risk area.*<br><br>*Greece: the regions Attica, Central Macdeonia, Eastern Macdeonia and Thrace, Epirus and*<br><br>*Thessaly are considered as additional risk areas.*<br><br>*Italy: the complete country is considered as risk area.*<br><br>*Latvia: the region Pierīga is considered as additional risk area.*<br><br>*Lithuania: the complete country is considered as risk area with exception of the county Utena.*<br><br>*Norway: the county Oslo is considered as risk area.*<br><br>*Portugal: the complete country is considered as risk area with exception of the autonomous*<br><br>*regions Azores and Madeira.*<br><br>*Sweden: the complete country is considered as risk area with exception of the province*<br><br>*Västernorrland." [DENPI, 3977G]* | | | |

| **Table S1 - NPI Descriptions** Date and descriptions of non pharmaceutical interventions in Germany from 01-10-2020 until 01-06-2021 | | | | | | |
|---|---|---|---|---|---|---|
| | **Name** | **Date** | **Description** | **Labels** | **State** | **End Date** |
| | | | *"From 8 November 2020, the Bavarian State Ministry for Health and Care in Germany decrees that all travellers from abroad who in the past 14 days have been to "risk areas" (based on information from the Robert Koch Institute) are obliged to quarantine themselves for 14 days upon arrival. Exceptions exist. Risk areas have changed when compared to the regulatory situation of November 1. The policy is slated to end on November 8. UPDATE: It expired on November 8." [DENPI, 3978G]* | | | |
| 37 | | 08-11-2020 | *"All people returning from a risk area to Saxony (Germany) (see Robert Koch-Institute for risk area definition) must get tested within ten days upon their entry to Saxony or provide a test result that is no older than 48 hours." [DENPI, 3975G]* | *Border Control* | Saxony | |
| 38 | | 09-11-2020 | *"From 9 November 2020, the Bavarian State Ministry for Health and Care in Germany decrees that all travelers from abroad who in the past 10 days have been to "risk areas" (based on information from the Robert Koch Institute) are obliged to* ***quarantine themselves for 10 days upon arrival****. Exceptions exist. Individuals are required to be tested if, within 10 days upon arrival, they show symptoms of a SARS-CoV-2 infection. Quarantine may be left for the duration of tests. Individuals that have entered Bavaria prior to November 9 have their duration of quarantine reduced to 10 days. The policy is slated to end on November 30." [DENPI, 3983G]* | *Border Control* | **Bavaria** | |
| 39 | **N3-c** | 10-11-2020 | *"For Germany, travel restrictions apply for entry from a large number of countries. These are issued by the Federal Ministry of the Interior, Building and Community. In principle, entry is possible from:* | *Border control* | **Nationwide** | **30-11-2020** |

| Table S1 - NPI Descriptions Date and descriptions of non pharmaceutical interventions in Germany from 01-10-2020 until 01-06-2021 | | | | | | |
|---|---|---|---|---|---|---|
| | **Name** | **Date** | **Description** | **Labels** | **State** | **End Date** |
| | | | *- EU member states associated with Schengen: Iceland, Norway, Switzerland and Liechtenstein the United Kingdom;*<br><br>*- Other countries, from which entry is possible due to the epidemiological situation assessment by the EU.*<br><br>*- Entry from other countries is only possible in exceptional cases and is conditional on there being an urgent Need." [OXF, 316AC]* | | | **(more restrictions )** |
| 40 | | 11-11-2020 | *Bremen (Germany) enforced a 10-day quarantine for persons who enter Bremen by land, sea or air from a risk area where they have stayed for 10 days. This does not pertain to person who spend less than 72 hours abroad, commuters (with residence in Bremen or abroad), system-relevant workers, vacation returnees who were tested negatively for SARS-CoV-2 no more than 48 hours before entry into the Federal Republic of Germany or immediately upon entry.*<br><br>*All people entering are obliged to undertake a digital entry registration at https:\\www.einreiseanmeldung.de. Quarantine shall end at the earliest from the fifth day after entry if a person has a negative corona test result from a test that was undertaken 5 days after entry in Germany." [DENPI, 3988G]*<br><br>*"Bremen (Germany) enforced a 14-day quarantine for persons who enter Bremen by land, sea or air from a risk area where they have stayed for 10 days. This does not pertain to person who spend less than 72 hours abroad, commuters (with residence in Bremen or abroad), system-relevant workers, vacation returnees who were tested negatively for SARS-CoV-2 no more than 48 hours before entry into the Federal* | *Border Control* | Bremen | |

| Table S1 - NPI Descriptions Date and descriptions of non pharmaceutical interventions in Germany from 01-10-2020 until 01-06-2021 | | | | | | |
|---|---|---|---|---|---|---|
| | **Name** | **Date** | **Description** | **Labels** | **State** | **End Date** |
| | | | *Republic of Germany or immediately upon entry. UPDATE: The original 14-day quarantine for people returning from abroad with exceptions was set to last until November 30, but ended on November 10 due to a change in the quality of the policy." [DENPI, 3989G]*<br><br>*"Bremen (Germany) is enforcing an exception from the 14-day quarantine for people returning or traveling to Bremen from abroad. The quarantine can be shorted to 5 days if after entry the person has a negative corona test result on paper or in an electronic document in German, English or French and immediately submits it to health authorities within ten days after the entry. The underlying test must have been performed at least five days after entry into the Federal Republic of Germany and must be kept for at least ten days after entry. UPDATE: The original quarantine exception after the fifth day with a positive corona test was set to last until November 30, it has been extended to January 9, 2021."* | | | |
| 41 | | 15-11-2020 | *"From 15 November 2020, the Bavarian State Ministry for Health and Care in Germany decrees that all travellers from abroad who in the past 10 days have been to "risk areas" (based on information from the Robert Koch Institute) are obliged to quarantine themselves for 10 days upon arrival. Exceptions exist. Risk areas have changed when compared to the regulatory situation of November 9. Individuals are required to be tested if, within 10 days upon arrival, showing symptoms of a SARS-CoV-2 infection. Quarantine may be left for the duration of tests. Individuals that have entered Bavaria prior to November 9 have their duration of quarantine reduced to 10 days. The policy is slated to end on November 30." [DENPI, 4002G]* | *Border Control* | **Bavaria** | |

| Table S1 - NPI Descriptions Date and descriptions of non pharmaceutical interventions in Germany from 01-10-2020 until 01-06-2021 | | | | | | |
|---|---|---|---|---|---|---|
| | **Name** | **Date** | **Description** | **Labels** | **State** | **End Date** |
| | | | *“From 9 November 2020, the Bavarian State Ministry for Health and Care in Germany decrees that all travellers from abroad who in the past 10 days have been to "risk areas" (based on information from the Robert Koch Institute) are obliged to quarantine themselves for 10 days upon arrival. Exceptions exist. Individuals are required to be tested if, within 10 days upon arrival, showing symptoms of a SARS-CoV-2 infection. Quarantine may be left for the duration of tests. Individuals that have entered Bavaria prior to November 9 have their duration of quarantine reduced to 10 days. The policy is slated to end on November 30. UPDATE: From 15 November 2020, risk areas have changed in the following way:*<br><br>*"Canada: the complete country is considered as risk area.*<br><br>*Estonia: the regions Harju, Hiiu and Rapla are considered as additional risk areas.*<br><br>*France: the French overseas territory French Polynesia is considered as additional risk area.*<br><br>*Greece: the regions North Aegean and Peloponnese are considered as additional risk areas.*<br><br>*Latvia: the region Zemgale is considered as additional risk area.*<br><br>*Norway: the counties Vestland and Viken are considered as additional risk areas.*<br><br>*Sweden: the complete country is considered as risk area.*<br><br>*United Kingdom of Great Britain and Northern Ireland: the Channel Island Jersey is*<br><br>*considered as additional risk area.* | | | |

| **Table S1 - NPI Descriptions** Date and descriptions of non pharmaceutical interventions in Germany from 01-10-2020 until 01-06-2021 | | | | | | |
|---|---|---|---|---|---|---|
| | **Name** | **Date** | **Description** | **Labels** | **State** | **End Date** |
| | | | *The region Ostrobothnia in Finland is no longer considered as risk area." [DENPI, 4003G]* | | | |
| 42 | | 18-11-2020 | *"Bremen (Germany) has enforced a quarantine of 14 days for category 1 contact persons who has had close contact with an infected person (15 min face-to-face, less than 1,5 m distance, in a room with poor ventilation or without having worn a mouth cover." [DENPI, 4007G]* | *Border Control* | Bremen | |
| 43 | | 22-11-2020 | *"From 22 November 2020, the Bavarian State Ministry for Health and Care in Germany decrees that all travellers from abroad who in the past 10 days have been to "risk areas" (based on information from the Robert Koch Institute) are obliged to quarantine themselves for 10 days upon arrival. Exceptions exist. Risk areas have changed when compared to the regulatory situation of November 15. Individuals are required to be tested if, within 10 days upon arrival, showing symptoms of a SARS-CoV-2 infection. Quarantine may be left for the duration of tests. Individuals that have entered Bavaria prior to November 9 have their duration of quarantine reduced to 10 days. The policy is slated to end on November 30." [DENPI, 4013G]*<br><br>*"From 15 November 2020, the Bavarian State Ministry for Health and Care in Germany decrees that all travellers from abroad who in the past 10 days have been to "risk areas" (based on information from the Robert Koch Institute) are obliged to quarantine themselves for 10 days upon arrival. Exceptions exist. Risk areas have changed when compared to the regulatory situation of November 9. Individuals are required to be tested if, within 10 days upon arrival, showing symptoms of a SARS-CoV-* | *Border Control* | **Bavaria** | |

| Table S1 - NPI Descriptions Date and descriptions of non pharmaceutical interventions in Germany from 01-10-2020 until 01-06-2021 | | | | | | |
|---|---|---|---|---|---|---|
| | **Name** | **Date** | **Description** | **Labels** | **State** | **End Date** |
| | | | *2 infection. Quarantine may be left for the duration of tests. Individuals that have entered Bavaria prior to November 9 have their duration of quarantine reduced to 10 days. The policy is slated to end on November 30. UPDATE: From 22 November 2020, risk areas have changed in the following way:*<br><br>*"Botswana: the complete country is considered as risk area.*<br><br>*Finland: the region Uusimaa (contains capital city Helsinki) is considered as risk area.*<br><br>*Greece: the region Central Greece is considered as additional risk area.*<br><br>*Latvia: the complete country is considered as risk area.*<br><br>*Lithuania: the complete country is considered as risk area.*<br><br>*The complete country Iceland is no longer considered as risk area." [DENPI, 4014G]* | | | |
| 44 | **N4-a** | 29-11-2020 | *"From 29 November 2020, the Bavarian State Ministry for Health and Care in Germany decrees that all travellers from abroad who in the past 10 days have been to "risk areas" (based on information from the Robert Koch Institute) are obliged to quarantine themselves for 10 days upon arrival. Exceptions exist. Risk areas have changed when compared to the regulatory situation of November 22. Individuals are required to be tested if, within 10 days upon arrival, showing symptoms of a SARS-CoV-2 infection. Quarantine may be left for the duration of tests. Individuals that have entered Bavaria prior to November 9 have their duration of quarantine reduced to 10 days. The policy is slated to end on November 30." [DENPI, 4028G]* | *Border Control* | **Bavaria** | **06-12-2020**<br><br>**(change in risk area in Bavaria)** |

| **Table S1 - NPI Descriptions** Date and descriptions of non pharmaceutical interventions in Germany from 01-10-2020 until 01-06-2021 | | | | | | |
|---|---|---|---|---|---|---|
| | **Name** | **Date** | **Description** | **Labels** | **State** | **End Date** |
| | | | *"From 22 November 2020, the Bavarian State Ministry for Health and Care in Germany decrees that all travellers from abroad who in the past 10 days have been to "risk areas" (based on information from the Robert Koch Institute) are obliged to quarantine themselves for 10 days upon arrival. Exceptions exist. Risk areas have changed when compared to the regulatory situation of November 15. Individuals are required to be tested if, within 10 days upon arrival, showing symptoms of a SARS-CoV-2 infection. Quarantine may be left for the duration of tests. Individuals that have entered Bavaria prior to November 9 have their duration of quarantine reduced to 10 days. The policy is slated to end on November 30. UPDATE:* ***From 29 November 2020, risk areas have changed in the following way****:*<br><br>*"Estonia: the region Tartu is considered as additional risk area.*<br><br>*Greece: the region West Greece is considered as additional risk area.*<br><br>*Portugal: the Portuguese Mainland and the autonomous region Azores are considered as risk areas (with exception of the autonomous region Madeira).*<br><br>*The regions Midlands, South-West and West in Ireland are no longer considered as risk areas.*<br><br>*The region Peloponnes in Greece is no longer considered as risk area." [DENPI, 4029G]* | | | |
| 45 | **N4-b** | 30-11-2020 | *"According to the resolution of November 25th, a stricter contact restriction applies. Citizens are asked to reduce contact with other people outside of their own household to "an absolutely necessary minimum". The members of your own household and of another household may be in public* | *Gathering* | **Nationwide** | **16-12-2020**<br><br>**(more restrictening)** |

| Table S1 - NPI Descriptions Date and descriptions of non pharmaceutical interventions in Germany from 01-10-2020 until 01-06-2021 | | | | | | |
|---|---|---|---|---|---|---|
| | **Name** | **Date** | **Description** | **Labels** | **State** | **End Date** |
| | | | *together. The upper limit is **5 people** from December 1st." [OXF, 336R]* | | | |
| 46 | **N4-c** | 30-11-2020 | *"Since November 8th, a mandatory quarantine of ten days has been in effect for all people entering from risk areas. The quarantine can be shortened if travelers can present a negative corona test no earlier than the fifth day after arrival. There are no pandemic-related restrictions for entry from the other member states of the European Union as well as from the United Kingdom, Switzerland, Liechtenstein, Norway and Iceland. The current entry restrictions relate to entries from third countries. However, the respective infection protection regulations of the federal states (including with regard to quarantine and reporting obligations) as well as the obligation to digitally register for entry must be observed by all people entering from risk areas. Für Personen, die in anderen als den vorstehend genannten Drittstaaten ansässig sind, gelten die bisherigen Einreisebeschränkungen fort, d.h. sie dürfen nur nach Deutschland einreisen, wenn sie eine wichtige Funktion ausüben oder ihre Reise zwingend notwendig ist (siehe hierzu "Wann ist die zwingende Notwendigkeit der Einreise gegeben?"). Ansässig ist eine Person in einem Staat, wenn sie dort entweder ihren Wohnsitz oder ihren gewöhnlichen Aufenthalt hat. Ein Wohnsitz ist insbesondere dann begründet, wenn die letzten 6 Monate dort verbracht wurden. https://web.archive.org/save/https://www.zusammengegencorona.de/informieren/alltag-gestalten/#faqitem=e2df90f7-8f7a-5c51-b42f-ecaffbcfe5f9 https://web.archive.org/web/20201207221431/https://www.bmi.bund.de/SharedDocs/faqs/DE/themen/bevoelkerungsschutz/coronavirus/coronavirus-faqs.html;jsessionid=36F79BE0E1058BB* | *Border control (general policy)* | **Nationwide** | **14-02-2021**<br><br>**(more restrictions )**<br><br>**09-05-2021**<br><br>**Quarantine is removed for vaccinated peopels** |

| Table S1 - NPI Descriptions Date and descriptions of non pharmaceutical interventions in Germany from 01-10-2020 until 01-06-2021 | | | | | | |
|---|---|---|---|---|---|---|
| | **Name** | **Date** | **Description** | **Labels** | **State** | **End Date** |
| | | | *7E14840089EE197C4.1_cid373” [OXF, 336AC]* | | | |
| 47 | **N4-d** | 30-11-2020 | *https://www.baden-wuerttemberg.de/fileadmin/redaktion/dateien/PDF/Coronainfos/210210_CoronaVO_konsolidierte_Fassung_ab_210211.pdf* | | **Baden Württemberg** | |
| 48 | **N4-e** | 30-11-2020 | *A cloth face mask must be worn in enclosed public spaces. This also includes at the workplace, if a distance of 1.5 metres from other persons cannot be adhered to. In commercial facilities such as supermarkets, shopping centres and home improvement stores with a total sales space of more than 800 square metres, there are additional restrictions concerning the number of customers per square metre.*<br><br>*(1) Accommodation offers for tourist purposes are prohibited. The use of permanently rented or owned properties and permanently parked properties caravans, mobile homes and so on is exclusively by the authorized user no tourist use within the meaning of sentence 1. When operating community facilities on campsites and so on and when accommodating travelers including their catering, the hygiene and infection protection standards according to § 4 must be observed.*<br><br>*(2) Coach trips and other group trips with buses for tourist purposes are not permitted.* [NRWPnpi] | *Internal movements* | **North Rhine-Westphalia** | |
| 49 | **N4-f** | 30-11-2020 | *“From 29 November 2020, the Bavarian State Ministry for Health and Care in Germany decrees that all travellers from abroad who in the past 10 days have been to "risk areas" (based on information from the Robert Koch Institute) are obliged to* | *Border Control* | **Bavaria** | |

| Table S1 - NPI Descriptions Date and descriptions of non pharmaceutical interventions in Germany from 01-10-2020 until 01-06-2021 | | | | | | |
|---|---|---|---|---|---|---|
| | **Name** | **Date** | **Description** | **Labels** | **State** | **End Date** |
| | | | *quarantine themselves for 10 days upon arrival. Exceptions exist. Risk areas have changed when compared to the regulatory situation of November 22. Individuals are required to be tested if, within 10 days upon arrival, showing symptoms of a SARS-CoV-2 infection. Quarantine may be left for the duration of tests. Individuals that have entered Bavaria prior to November 9 have their duration of quarantine reduced to 10 days. The policy is slated to end on November 30. UPDATE: From 30 November 2020, quarantine measures are extended to December 20." [DENPI, 4030G]* | | | |
| 50 | **N4-g** | 01-12-2020 | *"From 1 December 2020, institutions of higher education are again to be closed for in-person learning."* *"From 1 December 2020, the Bavarian State Ministry for Health and Care in Germany re-introduces general restrictions on outdoor activities ("Allgemeine Ausgangsbeschränkungen") for areas where the incidence rate surpasses the threshold of 300. Here,* ***leaving one's home is only allowed if there are compelling reasons to do so."*** | *Internal movements* | **Bavaria** | |
| 51 | **N4-h** | 01-12-2020 | *"From 29 November 2020, the Bavarian State Ministry for Health and Care in Germany decrees that all travellers from abroad who in the past 10 days have been to "risk areas" (based on information from the Robert Koch Institute) are obliged to quarantine themselves for 10 days upon arrival. Exceptions exist. Risk areas have changed when compared to the regulatory situation of November 22. Individuals are required to be tested if, within 10 days upon arrival, showing symptoms of a SARS-CoV-2 infection. Quarantine may be left for the duration of tests. Individuals that have entered Bavaria prior to November 9 have* | *Border Control* | **Bavaria** | |

| Table S1 - NPI Descriptions Date and descriptions of non pharmaceutical interventions in Germany from 01-10-2020 until 01-06-2021 | | | | | | |
|---|---|---|---|---|---|---|
| | **Name** | **Date** | **Description** | **Labels** | **State** | **End Date** |
| | | | *their duration of quarantine reduced to 10 days. The policy is slated to end on November 30. UPDATE:* ***From 1 December 2020, exceptions for those who have been to risk areas for less than 24 hours or stay in Bavaria for no more than 24 hours are required to have compelling reasons for having stayed*** *(in risk areas) abroad such as occupational, official, commercial, educational, medical or familial reasons or for carrying out errands related to daily needs. Exceptions for those that stay in Bavaria for no more than 72 hours now also apply to those that have stayed in risk areas for less than 72 hours (this no longer applies to those that have to transport persons, commodities and goods by street, rail, ship or plane for professional reasons across borders which are now listed separately). Border crossers ("Grenzgänger") are no longer required to get tested on a regular basis." [DENPI, 4059G]* | | | |
| 52 | | 01-12-2020 | *"Persons entering Saxony (Germany) from abroad who have been in a foreign risk area (as published by the Robert Koch Institute) at any time in the ten days prior to entry must immediately remain permanently in their home for a period of ten days (previously 14 days) after entry. Domestic quarantine may be terminated no earlier than five days after entry by a negative Corona test. The test must have been taken no earlier than five days after entry. UPDATE: The policy was extended several times, with the last extension on January 18, 2021. All travelers returning from a risk area must provide a test result that is younger than 24 hours, or get tested immediately upon entry, or get tested within 48 hours after entry." [DENPI, 4060G]* | *Border Control* | Saxony | |

| Table S1 - NPI Descriptions Date and descriptions of non pharmaceutical interventions in Germany from 01-10-2020 until 01-06-2021 | | | | | | |
|---|---|---|---|---|---|---|
| | **Name** | **Date** | **Description** | **Labels** | **State** | **End Date** |
| 53 | | 01-12-2020 | *“From 1 December 2020, the Bavarian State Ministry for Health and Care in Germany decrees that all travellers from abroad who in the past 10 days have been to "risk areas" (based on information from the Robert Koch Institute) are obliged to quarantine themselves for 10 days upon arrival. Exceptions have changed when compared to the regulatory situation of November 30. Individuals are required to be tested if, within 10 days upon arrival, showing symptoms of a SARS-CoV-2 infection. Quarantine may be left for the duration of tests. The policy is slated to end on December 20.” [DENPI, 4056G]* | *Border Control* | **Bavaria** | |
| 54 | | 06-12-2020 | *“From 6 December 2020, the Bavarian State Ministry for Health and Care in Germany decrees that all travellers from abroad who in the past 10 days have been to "risk areas" (based on information from the Robert Koch Institute) are obliged to quarantine themselves for 10 days upon arrival. Exceptions exist. Risk areas have changed when compared to the regulatory situation of December 1. Individuals are required to be tested if, within 10 days upon arrival, showing symptoms of a SARS-CoV-2 infection. Quarantine may be left for the duration of tests. The policy is slated to end on December 20.” [DENPI, 4088G]*<br><br>*“From 1 December 2020, the Bavarian State Ministry for Health and Care in Germany decrees that all travelers from abroad who in the past 10 days have been to "risk areas" (based on information from the Robert Koch Institute) are obliged to quarantine themselves for 10 days upon arrival. Exceptions have changed when compared to the regulatory situation of November 30. Individuals are required to be tested if, within 10 days upon arrival, showing symptoms of a SARS-CoV-2 infection. Quarantine may be left for the* | *Border Control* | **Bavaria** | |

| Table S1 - NPI Descriptions Date and descriptions of non pharmaceutical interventions in Germany from 01-10-2020 until 01-06-2021 | | | | | | |
|---|---|---|---|---|---|---|
| | **Name** | **Date** | **Description** | **Labels** | **State** | **End Date** |
| | | | *duration of tests. The policy is slated to end on December 20. UPDATE: From 6 December 2020, risk areas have changed in the following way:*<br><br>*"Estonia: the regions Põlva, Viljandi and Võru are considered as additional risk areas.*<br><br>*Finland: the regions Päijät-Häme and North Ostrobothnia are considered as additional risk*<br><br>*areas.*<br><br>*The regions Central Greece and Epirus are no longer considered as risk areas.*<br><br>*The region South-East in Ireland is no longer considered as risk area."" [DENPI, 4089G]* | | | |
| 55 | | 08-12-2020 | *No changes for the last week. New changes are being proposed for the upcoming weeks Links - http://web.archive.org/web/20201214162625/https://www.deutschland.de/en/news/coronavirus-in-germany-informations http://web.archive.org/web/20201214162943/https://de.usembassy.gov/covid-19-information/ Details: - Germany is imposing a hard lockdown from Wednesday.Trade, with the exception of shops for daily needs, will be closed from 16 December to 10 January - For the days from 24 to 26 December, "meetings with 5 persons plus children up to 14 years of age in the closest family circle" are per* | *Border control (general policy)* | **Nationwide** | |
| 56 | **C-BY** | 09-12-2020 | *"From 9 December 2020, the Bavarian State Ministry for Health and Care in Germany enacts that hotels, accommodation providers, youth and country hostels, camping sites (or any provision of accommodation)* ***may only*** | *Internal movements* | **Bavaria** | |

| Table S1 - NPI Descriptions Date and descriptions of non pharmaceutical interventions in Germany from 01-10-2020 until 01-06-2021 | | | | | | |
|---|---|---|---|---|---|---|
| | **Name** | **Date** | **Description** | **Labels** | **State** | **End Date** |
| | | | ***provide (over-night) accommodation for convincingly necessary reasons, such as occupational or business-related purposes. Touristic accommodation is not allowed***. *Furthermore, they have to meet conditions according to § 14, section 2, numbers 1 to 5 of the source document. "*<br><br>*"From 6 December 2020, the Bavarian State Ministry for Health and Care in Germany decrees that all travellers from abroad who in the past 10 days have been to "risk areas" (based on information from the Robert Koch Institute) are obliged to quarantine themselves for 10 days upon arrival. Exceptions exist. Risk areas have changed when compared to the regulatory situation of December 1. Individuals are required to be tested if, within 10 days upon arrival, showing symptoms of a SARS-CoV-2 infection. Quarantine may be left for the duration of tests. The policy is slated to end on December 20. UPDATE: From 9 December 2020, exceptions to quarantining are slightly changed. The policy is extended to January 5, 2021." [DENPI, 4100G]*<br><br>*"From 6 December 2020, the Bavarian State Ministry for Health and Care in Germany decrees that all travellers from abroad who in the past 10 days have been to "risk areas" (based on information from the Robert Koch Institute) are obliged to quarantine themselves for 10 days upon arrival. Exceptions exist. Risk areas have changed when compared to the regulatory situation of December 1. Individuals are required to be tested if, within 10 days upon arrival, showing symptoms of a SARS-CoV-2 infection. Quarantine may be left for the duration of tests. The policy is slated to end on December 20. UPDATE: From 9 December 2020, exceptions to quarantining are slightly changed. The policy is extended to January 5, 2021." [DENPI, 4101G]* | | | |

| **Table S1 - NPI Descriptions** Date and descriptions of non pharmaceutical interventions in Germany from 01-10-2020 until 01-06-2021 | | | | | | |
|---|---|---|---|---|---|---|
| | **Name** | **Date** | **Description** | **Labels** | **State** | **End Date** |
| 57 | **N5-a** | 14-12-2020 | *"In Saxony (Germany), **schools, boarding schools and daycare facilities will be closed up** to and including January 8, 2021." [DENPI, 4117G]*<br><br>*"The Free State of Saxony (Germany) issued a prohibition of movement. An **extended curfew applies between 10 p.m. and 6 a.m. the following day**. Leaving the accommodation during this time is only permitted for valid reasons." [DENPI, 4120G]*<br><br>*"Saxony (Germany) introduces **a prohibition of drinking and selling alcohol in public**. The dispensing of alcoholic beverages is permitted only in containers that can be carried and sealed." [DENPI, 4122G]*<br><br>*"In Saxony (Germany) closure is required for **shopping centers and retailers,** as well as for retail shops, with the exception of permissible telephone and online offers for shipping or delivery only." [DENPI, 4142G]* | *Schools, Internal movement (night curfew), Workplaces* | Saxony | |
| 58 | **N5-b** | 16-12-2020 | *"New Lockdown in place, starting on December 15th, provisional running until January 10th. Contacts in schools, too, are to be drastically reduced between 16 December 2020 and 10 January 2021. During this period, **children should be kept at home** wherever possible." [OXF, 352I]*<br><br>*"**Retail shops will close** from 16 December 2020 until 10 January 2021, with the exception of grocery stores, open-air food markets, direct marketers of food products, food collection and delivery services, beverage shops, health food shops, specialist baby shops, pharmacies, medical supply stores, drugstores, opticians, hearing aid shops, petrol stations, garages, bicycle workshops, banks and savings banks, post offices, dry cleaning services,* | *Schools, Workplaces, Internal movements* | **Nationwide** | **22-02-2021**<br><br>**(Schools reopen)**<br><br>**08-03-2021 all schools are open**<br><br>**, 10-03-2021 to 23-04-2021**<br><br>**,**<br><br>**10-02-2021** |

| **Table S1 - NPI Descriptions** Date and descriptions of non pharmaceutical interventions in Germany from 01-10-2020 until 01-06-2021 | | | | | | |
|---|---|---|---|---|---|---|
| | **Name** | **Date** | **Description** | **Labels** | **State** | **End Date** |
| | | | *launderettes, newsagents, pet supply shops, animal feed shops, the sale of Christmas trees and wholesalers. The sale in grocery stores of non-food products that are not related to daily needs can also be restricted and must on no account be extended." [OXF, 352L]*<br><br>*"The Federal Government and the Länder urgently appeal to all citizens to **avoid all non-essential movement** and **recommend that they stay at home**. Individual federal states also impose specific exit restrictions. For example, in the southern states in particular, leaving the home is only permitted if there are valid reasons for doing so (sports, shopping, doctor's appointments, etc.). **Local night-time curfew restrictions apply**, which generally prohibit leaving the home except in emergencies." [OXF, 352X]*<br><br>*"The Federal Government and the Länder urgently appeal to all citizens to **avoid all non-essential travel within and outside Germany** between now and 10 January. Individual federal states also impose specific exit restrictions. For example, in Mecklenburg Vorpommer, travel from other German states is currently not allowed." [OXF, 352AA]* | | | **,**<br><br>**05-01-2021**<br><br>**(internal movement more restricted by 15 km law)** |
| 59 | **N5-c** | 16-12-2020 | *"New Lockdown in place, starting on December 15th, provisional running until January 10th. For Germany, travel restrictions apply for entry from a large number of countries. These are issued by the Federal Ministry of the Interior, Building and Community (...) In principle, entry is possible from: - EU member states associated with Schengen: Iceland, Norway, Switzerland and Liechtenstein the United Kingdom; - Other countries, from which entry is possible due to the epidemiological situation assessment by the EU. - Entry* | *Border control (general policy)* | **Nationwide** | **14-02-2021**<br><br>**(more restricted)** |

| **Table S1 - NPI Descriptions** Date and descriptions of non pharmaceutical interventions in Germany from 01-10-2020 until 01-06-2021 | | | | | | |
|---|---|---|---|---|---|---|
| | **Name** | **Date** | **Description** | **Labels** | **State** | **End Date** |
|  |  |  | *from other countries is only possible in exceptional cases and is conditional on there being an urgent Need." Online entry registration is obligatory for travellers entering Germany from high-risk areas abroad, and that quarantine is mandatory for a period of 10 days following return. Quarantine can only be ended with a negative test, which can be taken on the 5th day after entry at the earliest. http://web.archive.org/web/20201129121721/https://www.auswaertiges-amt.de/en/coronavirus/2317268 "[XOF,352AC]* |  |  |  |
| 60 | **N5-d** | 16-12-2020 | *"Saxony (Germany)* ***closes hair salons****." [DENPI, 4160G]* | *Workplaces* | Saxony | 07-02-2021 |
| 61 | **N5-e** | 16-12-2020 | *"From January 9 until prospectively Feb 14 persons who enter Hesse by land, sea or air from abroad and have been in a risk area for infection with SARS-CoV-2 in the last 10 days before entry must be tested and quarantined. This also applies to persons who first entered another county of the Federal Republic of Germany." [DENPI, 4152G]* | *Border Control* | Hesse | 14-02-2021 |
| 62 | **N6** | 22-12-2020 | *"Since 22 December 2020, travelers from the* ***United Kingdom as well as South Africa*** *and those who have stayed there in the last ten days prior to entry must be tested for the Coronavirus. The* ***existing quarantine obligation*** *for entries from the risk areas applies without prejudice.*<br><br>*The reason for this measure is a new variant (mutation) of the corona virus found in the United Kingdom and South Africa, which, according to the British government, is up to 70 per cent easier to* | *Border control* | **Nationwide** | |

| **Table S1 - NPI Descriptions** Date and descriptions of non pharmaceutical interventions in Germany from 01-10-2020 until 01-06-2021 | | | | | | |
|---|---|---|---|---|---|---|
| | **Name** | **Date** | **Description** | **Labels** | **State** | **End Date** |
| | | | *transmit than the previously known variant SARS-CoV-2." [DeCov]* | | | |
| 63 | | 31-12-2020 | *"In Saxony (Germany) people returning from abroad must get tested immediately upon entry, or within 48 hours after if they cannot provide a test result that is no older than 24 hours." [DENPI, 4210G]* | *Border Control* | Saxony | |
| 64 | **N7-a** | 05-01-2021 | *"From the 5th of January, residents in hotspot districts (more than 200 per 100,000 cases) are* ***not permitted to travel more than 15 km from their town*** *without a valid reason. Day trips are not classified as a valid reason. " [OXF, 372AA]*<br><br>*"From the 5th of January,* ***private meetings*** *with one other person not living in the same household are permitted (previously a maximum of 5 from 2 households.)" [OXF, 273R]* | *Internal movements, Gathering* | **Nationwide** | **08-03-2021 to 01-08-2021**<br><br>**The law is removed,**<br><br>**08-03-2021 it is loosening** |
| 65 | **N7-b** | 06-01-2021 | *"In future, anyone entering the country from a foreign risk area must be tested on entry or in the 48 hours prior to entry. The obligation to undergo a ten-day quarantine, which can be terminated from the fifth day onwards by a negative test, remains in place." [deCov1]* | *Border Control* | **Nationwide** | |
| 66 | **N7-c** | 10-01-2021 | *"**Bremen (Germany) enforced a 10-day quarantine for persons who enter Bremen by land, sea or air from a risk area** where they have stayed for 10 days. This does not pertain to person who spend less than 72 hours abroad, commuters (with residence in Bremen or abroad), system-relevant workers, vacation returnees who were tested negative for SARS-CoV-2 no more than 48 hours before entry into the Federal* | *Border control, Internal movements* | Bremen | **28-03-2021** |

| Table S1 - NPI Descriptions Date and descriptions of non pharmaceutical interventions in Germany from 01-10-2020 until 01-06-2021 | | | | | | |
|---|---|---|---|---|---|---|
| | **Name** | **Date** | **Description** | **Labels** | **State** | **End Date** |
| | | | *Republic of Germany or immediately upon entry." [DENPI, 4221G]* | | | |
| 67 | **N7-d** | 10-01-2021 | *"Bremen (Germany) is enforcing a 14 day quarantine for all persons who have been confirmed via laboratory diagnosis with coronavirus SARS-CoV-2. UPDATE: The policy has been extended to March 28, 2021." [DENPI, 4220G]*<br><br>*"Bremen (Germany) enforced a 10-day quarantine for persons who enter Bremen by land, sea or air from a risk area where they have stayed for 10 days. This does not pertain to person who spend less than 72 hours abroad, commuters (with residence in Bremen or abroad), system-relevant workers, vacation returnees who were tested negatively for SARS-CoV-2 no more than 48 hours before entry into the Federal Republic of Germany or immediately upon entry.*<br><br>*All people entering are obliged to undertake a digital entry registration at https:\\www.einreiseanmeldung.de.*<br><br>*UPDATE: The policy was extended again to March 28, 2021." [DENPI, 4221G]*<br><br>*"Bremen (Germany) enforced a 10-day quarantine for persons who enter Bremen by land, sea or air from a risk area where they have stayed for 10 days. This does not pertain to person who spend less than 72 hours abroad, commuters (with residence in Bremen or abroad), system-relevant workers, vacation returnees who were tested negatively for SARS-CoV-2 no more than 48 hours before entry into the Federal Republic of Germany or immediately upon entry.*<br><br>*All people entering are obliged to undertake a digital entry registration at https:\\www.einreiseanmeldung.de. Quarantine shall end at the earliest from* | *Border Control* | Bremen | 28-03-2021 |

| Table S1 - NPI Descriptions Date and descriptions of non pharmaceutical interventions in Germany from 01-10-2020 until 01-06-2021 | | | | | | |
|---|---|---|---|---|---|---|
| | **Name** | **Date** | **Description** | **Labels** | **State** | **End Date** |
| | | | *the fifth day after entry if a person has a negative corona test result from a test that was undertaken 5 days after entry in Germany.*<br><br>*UPDATE: The original quarantine for persons returning from risk areas with several exceptions who have to digitally register was set to last until November 30, it has been extended to January 9, 2021." [DENPI, 4222G]*<br><br>*"Bremen (Germany) has enforced a quarantine of 14 days for category 1 contact persons who has had close contact with an infected person (15 min face-to-face, less than 1,5 m distance, in a room with poor ventilation or without having worn a mouth cover. UPDATE: The policy was extended to March 28, 2021. Since January 10, 2021, in the case of a person infected with the novel variant of the coronavirus B.1.1.7, B.1.351 or B.1.1.28, isolation shall not be terminated until a negative result of a PoC antigen test is demonstrated. If the infected person has been found to be infected with a novel variant of coronavirus, the period of isolation for category I contacts shall be twenty-one days. Isolation may be terminated only from the fourteenth day after the last contact with the infected person, if the contact person has a negative test result with regard to test result.*<br><br>*Isolation of category contacts shall end at the earliest from the fifth day after the last contact within the same cohort if the contact has a negative test result obtained during isolation at the earliest from the day five after the last contact with the person who had coronavirus. The period of isolation shall not be reduced if the infected person has been found to be infected with a novel variant of the coronavirus." [DENPI, 4223G]* | | | |

| Table S1 - NPI Descriptions Date and descriptions of non pharmaceutical interventions in Germany from 01-10-2020 until 01-06-2021 | | | | | | |
|---|---|---|---|---|---|---|
| | **Name** | **Date** | **Description** | **Labels** | **State** | **End Date** |
| 68 | **N7-e** | 11-01-2021 | *Since Monday [11-01-2021], the measures taken by the federal and state governments on 5 January to contain the Corona pandemic have come into force nationwide. [DeCov2]*<br><br>***meetings** in the future are to be possible only with a person who is not part of the state's own household. In counties where more than 200 new infections per 100,000 inhabitants have been reported within seven days, people **should not be allowed to travel more than 15 kilometers from their place of residence** without a valid reason. "Day-trip excursions explicitly do not constitute a valid reason.*<br><br>*In future, **anyone entering the country from a foreign risk area must be tested on entry or in the 48 hours prior to entry. The obligation to undergo a ten-day quarantine**, which can be terminated from the fifth day onwards by a negative test, remains in place.* | *Internal movements, Gathering, Border control* | **Nationwide** | |
| 69 | **N7-f** | 11-01-2021 | *"In **Saxony-Anhalt**, Germany the Prime Minister of Saxony-Anhalt announced on January 8th, 2021 that **movement restrictions** will apply. The radius of movement of 15 kilometers around one's own place of residence applies from an incidence rate of 200 and this incidence rate lasts for a period of 5 days." [DENPI, 4237G]* | *Internal movements* | Saxony-Anhalt | 10-03-2021 |
| 70 | **N7-g** | 11-01-2021 | *"As of January 11, 2021 a 'two-test strategy' has been in force in **Lower Saxony** (Germany) for e**ntries from risk areas (according to RKI), in addition to the 14-day quarantine obligation:** a first test must be carried out on entry from a risk area, then a 14-day quarantine begins regardless of the test result. However, this can be ended prematurely on the fifth day* | *Border Control* | Lower Saxony | |

| **Table S1 - NPI Descriptions** Date and descriptions of non pharmaceutical interventions in Germany from 01-10-2020 until 01-06-2021 | | | | | | |
|---|---|---|---|---|---|---|
| | **Name** | **Date** | **Description** | **Labels** | **State** | **End Date** |
| | | | *of quarantine if the result of a second test (PCR test) is negative. The testing obligation can be fulfilled for entry from (normal) risk areas by testing within 48 hours before arrival or by testing immediately after entry. Only when entering or returning from a "normal" risk area is it possible to be "cleared" via a PCR test. In high-incidence or virus-variant areas, the fourteen-day obligation of isolation applies without exception." [DENPI, 4233G]* | | | |
| 71 | **N7-h** | 11-01-2021 | *"In* ***Saxony*** *(Germany) public meetings are allowed with* ***one person from another household."*** *[DENPI, 4241G]* | *Gatherin g* | *Saxony* | *07-02-2021* |
| 72 | **N7-i** | 11-01-2021 | *"From January 11 until prospectively Feb. 14, contact restrictions in* ***Hesse*** *(Germany) were tightened. In public spaces,* ***members of a household may only meet with one other person****. Children also count towards this limit. There are exceptions for joint childcare of up to three families or for the accompaniment and care of minors or persons in need of support. Previously, meetings of relatives from two households, but no more than five people, were allowed." [DENPI, 4242G]* | *Gatherin g* | *Hesse* | *14-02-2021* |
| 73 | **N7-j** | 11-01-2021 | *"In Saxony (Germany) public meetings are allowed with* ***one person from another household****." [DENPI, 4241G]* | *Gatherin g* | *Saxony* | *07-02-2021* |
| 74 | **N7-k** | 12-01-2021 | ***Berlin*** *introduced a new quarantine policy on January 12, 2021. Now,* ***a corona test needs to be no older than 48 hours when entering Berlin from a risk area abroad.*** *Additionally, a* ***10-quarantine*** *is instituted* | *Border control* | *Berlin* | |

| **Table S1 - NPI Descriptions** Date and descriptions of non pharmaceutical interventions in Germany from 01-10-2020 until 01-06-2021 | | | | | | |
|---|---|---|---|---|---|---|
| | **Name** | **Date** | **Description** | **Labels** | **State** | **End Date** |
| | | | *which can be shortened through a corona test on the 5th day. [DENPI, 4247G]* | | | |
| 75 | **N7-l** | 14-01-2021 | *"The president of the Robert Koch Institute (RKI) has called on people in Germany to refrain from non-essential travel, partly because of new coronavirus variants. "Anyone who does not absolutely have to should not travel at the moment," said Lothar Wieler in Berlin on Thursday." [deCov1]* | *Border Control* | **Nationwide** | |
| 76 | **N7-m** | 14-01-2021 | *"In future, anyone entering the Federal Republic of Germany from a risk area must be able to prove that they are not infected with the coronavirus no later than 48 hours after entering the country. Travelers from particularly affected regions must present a negative test result before entering the country. The regulation comes into force on 14 January. In addition, travelers who have been in a risk area in the last ten days before entry must use the electronic entry declaration (DEA). Those entering from an area outside the Schengen area must also present the DEA proof at the entry control. The quarantine obligations ordered by the federal states for entry from risk areas continue to apply." [DECov]* | *Border control* | **Nationwide** | |
| 77 | | 16-01-2021 | *"Upon receipt of a positive Corona test result, in Hesse (Germany) from Dec 16 on one is obligated to immediately seek a suitable accommodation for quarantine - without separate order of the Public Health Department. One must seclude themself for 14 days, i.e. stay there permanently, avoid contact with other people as far as possible, and do not receive any visitors. The responsible Public Health Department is to be immediately notified." [DENPI, 4250G]* | *Quaranti ne* | Hesse | |

| Table S1 - NPI Descriptions Date and descriptions of non pharmaceutical interventions in Germany from 01-10-2020 until 01-06-2021 | | | | | | |
|---|---|---|---|---|---|---|
| | **Name** | **Date** | **Description** | **Labels** | **State** | **End Date** |
| 78 | | 17-01-2021 | *No changes. All measures extended till 31st Jan 2021 remains Links https://web.archive.org/web/20210126231018/https://www.bundesregierung.de/resource/blob/656632/1834616/414f0f73976e720fd7fb7b33c74b30fb/2021-01-05-mpk-beschluss-corona-en-data.pdf https://web.archive.org/web/20210126230736/https://www.deutschland.de/en/news/coronavirus-in-germany-informations" [OXF,384AC]* | *Border control (general policy)* | **Nationwide** | |
| 79 | **N8-a** | 18-01-2021 | *From 18 January 2021, the Bavarian State Ministry for Health and Care in Germany decrees that individuals are urged to reduce physical contacting as much as possible, keep their social circle constant and maintain a minimum distance of 1,5 meters to others. In public spaces (whenever minimum distance measures cannot be followed), individuals are obliged to wear a face mask. In public transport (outside of long-distance traffic), individuals are obliged to wear a FFP2 mask. Exceptions exist.* | *Masks* | **Bavaria** | |
| 80 | **N8-b** | 19-01-2021 | *On 19 January, the German Chancellor Angela Merkel announced that all existing measures to contain the corona pandemic continue to apply and are temporarily extended until 14 February 2021. Face masks are mandatory in all places with public circulation. Medical masks (OP masks) or FFP2 masks (or KN95 or N95 masks) are to be worn in public transport and when shopping, according to the decision of the federal and state governments on 19 January.* | *Mask* | **Nationwide** | **2023** |
| 81 | **N8-c** | 20-01-2021 | *Since January 20, the following applies in Hamburg: For travelers arriving and returning from risk countries, there is an* | | | |

| Table S1 - NPI Descriptions Date and descriptions of non pharmaceutical interventions in Germany from 01-10-2020 until 01-06-2021 | | | | | | |
|---|---|---|---|---|---|---|
| | **Name** | **Date** | **Description** | **Labels** | **State** | **End Date** |
| | | | *obligation to test a maximum of 48 hours before or immediately after entry and, regardless of the result, a ten-day quarantine. This can be shortened by a negative test from the fifth day after arrival at the earliest - this must be a PCR test, antigen testing is not permitted. The travelers have to bear the costs of the tests themselves. In Hamburg there are no exceptions for people who have been vaccinated or who have already had a corona disease.* | | | |
| 82 | **N8-d** | 22-01-2021 | *In Hamburg, simple cloth masks are no longer sufficient to increase the protective effect. Instead, medical masks must be worn from January 22, 2021. They protect better than everyday masks made of fabric and are subject to tested standards.*<br><br>*Medical masks are mandatory*<br><br>*in public transport, on flights, in taxis and other commercial transport offers,*<br>*while shopping,*<br>*in public buildings,*<br>*in care facilities, hospitals and for health treatments,*<br>*at church services*<br>*as well as in a work context.*<br><br>*Medical masks include in particular medical face masks or surgical or respiratory protection masks as well as FFP2 masks and comparable masks with the KN95 standard.* | *Mask* | **Hamburg** | |

| **Table S1 - NPI Descriptions** Date and descriptions of non pharmaceutical interventions in Germany from 01-10-2020 until 01-06-2021 | | | | | | |
|---|---|---|---|---|---|---|
| | **Name** | **Date** | **Description** | **Labels** | **State** | **End Date** |
| | | | *Medical masks must be worn by anyone over the age of 14. Children can still use everyday fabric masks.*<br><br>*There is still a general mask requirement for outdoor events and gatherings. Cloth masks are still allowed here. However, we recommend using medical masks consistently. Medical masks must be worn in closed rooms. Note that events and meetings are strictly prohibited and require official approval.* | | | |
| 83 | | 22-01-2021 | *"Since 22 January, 25 countries have been newly designated as high incidence areas, including, for example,* ***the Czech Republic, Spain and the USA****. Great Britain, Ireland, South Africa and Brazil continue to be considered virus variant areas." [DECov]* | *Border control*<br><br>*(Change of virus variant areas)* | **Nationwide** | |
| 84 | | 23-01-2021 | *Berlin (Germany) closes all libraries from January 23, 2021. All libraries provide comprehensive digital offers instead.* | | **Berlin** | |
| 85 | **N8-f** | 24-01-2021 | *In Berlin (Germany), of January 24, 2021,* ***medical masks are compulsory in public transport including railroad stations, long-distance transport, airports, ferries, trade, doctors' and therapists' surgeries, hospitals, nursing homes and religious institutions.***<br><br>*Everyday masks may be worn in schools (including on the entire grounds), in office and administrative buildings, in elevators, at gatherings, at weekly markets, and in* | | **Berlin** | |

| Table S1 - NPI Descriptions Date and descriptions of non pharmaceutical interventions in Germany from 01-10-2020 until 01-06-2021 | | | | | | |
|---|---|---|---|---|---|---|
| | **Name** | **Date** | **Description** | **Labels** | **State** | **End Date** |
| | | | *certain crowded places and streets. [DENPI,4268G]* | | | |
| 86 | | 24-01-2021 | *Stricter rules on entering Germany have been in effect since midnight on Sunday for more than 20 countries with particularly high corona infection rates. These high-risk areas include the neighboring* ***Czech Republic****, the holiday destinations* ***Portugal, Spain*** *and* ***Egypt*** *as well as the* ***USA****. Anyone who wants to enter from there must be able to show a* ***negative corona test*** *(PCR test or laboratory or rapid tests of comparable quality) at the border.[DeCov]* | *Border control*<br>*(Change in high risk area)* | **Nationwide** | |
| 87 | **N8-g** | 25-01-2021 | *As of January 25, 2021 in* ***Lower Saxony, medical masks (KN95/N95 or FFP2 or surgical masks) must be worn in public transport****, in shops and around them. Children up to the age of six and people with illnesses that do not permit the wearing of a mask, such as cardiovascular diseases, are exempt. For children between the ages of 6 and 14, it is sufficient to wear a so-called everyday mask made of fabric.* | *Mask* | **Lower Saxony** | |
| 88 | **N8-h** | 28-01-2021 | *Saxony (Germany), as of January 28, 2021, introduces an obligation to wear a* ***medical mask*** *when using public transportation, in front of the entrance area of and in wholesale and retail stores, as well as in health care facilities and for gatherings in churches and religious worship.*<br><br>*The obligation to wear FFP2 masks or the comparable standard KN95/N95 applies from Thursday to employees of outpatient care services when practicing care, when visiting day care facilities, in care facilities for visitors, in correctional facilities, refugee shelters for staff and visitors.* | *Mask* | **Saxony** | |

| **Table S1 - NPI Descriptions** Date and descriptions of non pharmaceutical interventions in Germany from 01-10-2020 until 01-06-2021 | | | | | | |
|---|---|---|---|---|---|---|
| | **Name** | **Date** | **Description** | **Labels** | **State** | **End Date** |
| | | | *Children up to the age of 6 remain exempt from the mask requirement.* | | | |
| 89 | **N8-i** | 89 | *“Saxony (Germany), as of January 28, 2021, introduces an obligation to wear a **medical mask** when using public transportation, in front of the entrance area of and in wholesale and retail stores, as well as in health care facilities and for gatherings in churches and religious worship.*<br><br>*The obligation to wear FFP2 masks or the comparable standard KN95/N95 applies from Thursday to employees of outpatient care services when practicing care, when visiting day care facilities, in care facilities for visitors, in correctional facilities, refugee shelters for staff and visitors. Children up to the age of 6 remain exempt from the mask requirement.” [DENPI, 4273G]* | | Saxony | |
| 90 | **N9** | 30-01-2021 | *For countries were particularly infectious variants of the corona virus have spread widely, a far-reaching travel ban will apply in Germany from Saturday, 30 January. The countries affected are **Brazil, Great Britain, Ireland, Portugal and South Africa**; on Sunday, the African states of Eswatini and Lesotho will be added. The ban on **airlines, rail, bus and shipping companies** decided by the cabinet will apply until 17 February, but provides for numerous exceptions, including for all Germans and foreigners living in “Germany, as well as for transit passengers and the movement of goods.” [DeCov]*<br><br>*“For countries where particularly infectious variants of the corona virus have spread widely, a far-reaching travel ban will apply in Germany from Saturday, 30 January. The countries affected are Brazil,* | *Border control* | **Nationwide** | |

| Table S1 - NPI Descriptions Date and descriptions of non pharmaceutical interventions in Germany from 01-10-2020 until 01-06-2021 | | | | | | |
|---|---|---|---|---|---|---|
| | **Name** | **Date** | **Description** | **Labels** | **State** | **End Date** |
| | | | *Great Britain, Ireland, Portugal and South Africa; on Sunday, the African states of Eswatini and Lesotho will be added. The ban on airlines, rail, bus and shipping companies decided by the cabinet will apply until 17 February, but provides for numerous exceptions, including for all Germans and foreigners living in Germany, as well as for transit passengers and the movement of goods." [deCov1]* | | | |
| 91 | | 30-01-2021 | *"Travel restrictions apply for entry from a large number of countries. In principle, entry is possible from: • EU member states • states associated with Schengen: Iceland, Norway, Switzerland and Liechtenstein • Other countries, from which entry is possible due to the epidemiological situation assessment by the EU. Source: http://web.archive.org/web/20210202120840/https://www.auswaertiges-amt.de/en/einreiseundaufenthalt/coronavirus" [OXF,397AC]* | *Border control (general policy)* | **Nationwide** | |
| 92 | | 01-02-2021 | *In Bremen (Germany), as of February 01, 2021, medical masks must be worn everywhere where an obligation to wear a mask exists. This only applies to persons age 16 and above. [DENPI, 4321G]* | *Facial Mask* | **Bremen** | |
| 93 | | 12-02-2021 | *"No policy change. Source: http://web.archive.org/web/20210212104524/https://www.auswaertiges-amt.de/en/einreiseundaufenthalt/coronavirus"* | *Border control (general policy)* | **Nationwide** | |
| 94 | **N10** | 14-02-2021 | *"Germany is to close its borders with the **Czech Republic and the Austrian Tyrol region** -- both zones with high infection* | *Border control* | **Nationwide** | |

| **Table S1 - NPI Descriptions** Date and descriptions of non pharmaceutical interventions in Germany from 01-10-2020 until 01-06-2021 | | | | | | |
|---|---|---|---|---|---|---|
| | **Name** | **Date** | **Description** | **Labels** | **State** | **End Date** |
| | | | *rates of contagious COVID-19 variants." [OXF, 412AC]*<br><br>*"This is the first time that neighboring states of Germany with a common land border are virus variant areas. Therefore, measures must be taken to contain the pandemic and to strengthen protection against infection. To protect against virus mutations, the Federal Ministry of the Interior announced that entry from the corresponding virus variant areas would be restricted." [deCov2]*<br><br>*"With effect from 14 February 2021, the following have been classified as virus variant areas: The Czech Republic, Slovakia and Austria - in this case the federal state of Tyrol with the exception of the political district of Lienz (East Tyrol), the municipality of Jungholz and the Rißtal valley in the municipal area of Vomp and Eben am Achensee. The classification also applies to countries such as Great Britain, Ireland, Portugal, South Africa and Brazil, among others." [deCov2]* | | | |
| 95 | | 28-02-2021 | *"No policy change. A travel ban remains imposed on individuals coming from high-risk countries. Source: http://web.archive.org/web/20210307110134/https://www.bundesgesundheitsministerium.de/service/gesetze-und-verordnungen/guv-19-lp/coronaschv/coronaschv-en.html" [OXF,426AC]* | *Border control (general policy)* | **Nationwide** | |
| 96 | | 02-03-2021 | *"On Sunday, the Robert Koch Institute (RKI) classified the French department of Moselle near the border as a so-called virus variant area as of 00:00 on 2 March.In order to protect against the spread of viral mutations, persons entering Germany from the Département Moselle must be able to prove that they are not* | *Border Control* | **Nationwide** | |

| Table S1 - NPI Descriptions Date and descriptions of non pharmaceutical interventions in Germany from 01-10-2020 until 01-06-2021 | | | | | | |
|---|---|---|---|---|---|---|
| | **Name** | **Date** | **Description** | **Labels** | **State** | **End Date** |
| | | | *infected with the coronavirus as of 2 March. A negative test result by means of PCR or PoC antigen test is considered as proof. The federal and state police forces will intensify controls." [deCov2]* | | | |
| 97 | | 05-03-2021 | *Repeated: "The temporarily introduced border controls with the Czech Republic and Austria will also remain in place for the time being until 17 March 2021.*<br><br>*Virus variant areas are areas in which there is a particular risk of entry due to the spread of mutations of the virus. These areas are published on the website of the Robert Koch Institute and adapted to current developments. The Czech Republic, Slovakia and Austria – in this case the province of Tyrol (with some exceptions) and the department of Moselle in France – are now designated as virus variant areas. The classification also applies to countries such as Great Britain, Ireland, Portugal, South Africa and Brazil, among others.*<br><br>*People entering the country from virus-variant areas must always prove that they are not infected with the coronavirus. A negative test result by means of PCR or PoC antigen test is considered as proof. The test must have been taken no more than 48 hours before entry." [deCov2]* | *Border Control* | **Nationwide** | |
| 98 | **N11-a** | 08-03-2021 | *"From Mar. 08, 2021, the federal government **will finance a rapid test for all citizens in Germany at least once a week at local testing centers,** Federal Health Minister Jens Spahn announced." [DENPI,4345G]* | *Covid tests* | **Nationwide** | |

**Table S1 - NPI Descriptions** Date and descriptions of non pharmaceutical interventions in Germany from 01-10-2020 until 01-06-2021

| | Name | Date | Description | Labels | State | End Date |
|---|---|---|---|---|---|---|
| | | | *From Monday, the federal government will finance one rapid test per week for every citizen. In addition, retailers will offer self-tests. "There are more than enough of these quick tests, they are available, they are easy to order," says Federal Health Minister Jens Spahn. They can be done at local test centres and pharmacies from 8 March. [deCov]* | | | |
| 99 | | 08-03-2021 | *"No policy change. http://web.archive.org/web/20210307110134/https://www.bundesgesundheitsministerium.de/service/gesetze-und-verordnungen/guv-19-lp/coronaschv/coronaschv-en.html" [OXF,434AC]* | *Border control (general policy)* | **Nationwide** | |
| 100 | **N11-b** | 09-03-2021 | *In **Bremen** "Since March 9, 2021 for a person with a positive antigen test the obligation to segregate shall apply accordingly for a period of ten days. These requirements shall cease to apply if the first PCR test carried out on this person after the positive antigen test shows a negative result." [DENPI, 4375G]* | *Covid tests* | *Bremen* | *19-04-2021* |
| 101 | | 14-03-2021 | *"No policy change. Source: https://archive.fo/aDY19" [OXF,440AC]* | *Border control (general policy)* | **Nationwide** | |
| 102 | | 20-03-2021 | *"No policy change. Source: http://web.archive.org/web/20210327015622/https://www.bundesgesundheitsministerium.de/service/gesetze-und-verordnungen/guv-19-lp/coronaschv/coronaschv-en.html http://web.archive.org/web/20210327020057/https://www.auswaertiges-* | *Border control (general policy)* | **Nationwide** | |

| **Table S1 - NPI Descriptions** Date and descriptions of non pharmaceutical interventions in Germany from 01-10-2020 until 01-06-2021 | | | | | | |
|---|---|---|---|---|---|---|
| | **Name** | **Date** | **Description** | **Labels** | **State** | **End Date** |
| | | | *amt.de/en/einreiseundaufenthalt/coronavirus"* | | | |
| 103 | | 21-03-2021 | *"**Poland** has now been designated as a high incidence area. The classification as an area with a particularly high risk of infection was made with effect from 21 March 2021. Persons who have stayed in a high-incidence area within the last ten days before entering Germany are obliged to be tested before traveling to Germany. They must present a negative test result or an appropriate medical certificate to the carrier (for example the airline) before departure." [DeCov]*<br><br>*"Poland has now been designated as a high incidence area. The classification as an area with a particularly high risk of infection was made with effect from 21 March 2021. Persons who have stayed in a high-incidence area within the last ten days before entering Germany are obliged to be tested before travelling to Germany. They must present a negative test result or an appropriate medical certificate to the carrier (for example the airline) before departure.*<br><br>*The negative test result may also be required during checks by the Federal Police (for example, entry checks at the airport or checks close to the border when entering by land at internal borders that are free of border controls). The swab for the test must have been taken no earlier than 48 hours before entry.*<br><br>*According to the regulations of the German federal states, persons entering the country from risk areas and high-incidence areas can end the ten-day quarantine obligation prematurely by taking a second negative test. As a rule, this second test may be carried out at the earliest on the fifth day* | *Border Control* | **Nationwide** | **28-03-2021 (next one)** |

| **Table S1 - NPI Descriptions** Date and descriptions of non pharmaceutical interventions in Germany from 01-10-2020 until 01-06-2021 | | | | | | |
|---|---|---|---|---|---|---|
| | **Name** | **Date** | **Description** | **Labels** | **State** | **End Date** |
| | | | *after entry in the case of risk areas and high-incidence areas.*<br><br>*Quarantine is then terminated upon receipt of a negative test result. " [deCov2]* | | | |
| 104 | | 26-03-2021 | *"***Updated to add note. These measures take effect from March 30. Negative tests will be required for airport arrivals from Tuesday and all of France has been declared high-risk. From Sunday anyone traveling from France will have to submit a negative test and go into 10 days' quarantine. Random German checks and compulsory tests will be enforced on the French border, says France's foreign minister. "The pandemic in Germany is exploding faster than they thought," said Jean-Yves Le Drian. http://web.archive.org/web/20210328112220/https://www.bbc.com/news/world-europe-56537389 " [OXF,452AC]* | *Border control (general policy)* | **Nationwide** | |
| 105 | | 27-03-2021 | *"Bremen (Germany) enforced a 10-day quarantine for persons who enter Bremen by land, sea or air from a risk area where they have stayed for 10 days. This does not pertain to person who spend less than 72 hours abroad, commuters (with residence in Bremen or abroad), system-relevant workers, vacation returnees who were tested negatively for SARS-CoV-2 no more than 48 hours before entry into the Federal Republic of Germany or immediately upon entry.*<br><br>*All people entering are obliged to undertake a digital entry registration at https:\\www.einreiseanmeldung.de.*<br><br>*UPDATE: The policy is extended again to April 19, 2021. From March 27, for persons who have stayed in a virus-variant area in* | *Border Control* | Bremen | |

| Table S1 - NPI Descriptions Date and descriptions of non pharmaceutical interventions in Germany from 01-10-2020 until 01-06-2021 | | | | | | |
|---|---|---|---|---|---|---|
| | **Name** | **Date** | **Description** | **Labels** | **State** | **End Date** |
| | | | *the last ten days before entry, the period of isolation is 14 days." [DENPI, 4391G]* | | | |
| 106 | **N12-a** | 28-03-2021 | *"**France, the Czech Republic and Slovakia** are considered high incidence areas from 28 March 00:00. Persons who have stayed in a high incidence area within the last ten days before entering Germany are obliged to be tested before traveling to Germany. They must present a negative test result or an appropriate medical certificate to the carrier (for example the airline) before departure. According to the regulations of the German federal states, persons entering the country from risk areas and high-incidence areas can end the ten-day quarantine obligation prematurely by taking a second negative test. As a rule, this second test may be carried out at the earliest on the fifth day after entry in the case of risk areas and high-incidence areas. Quarantine is then terminated upon receipt of a negative test result. The competent authority may check the evidence of the second negative test until the end of the general quarantine period, i.e. until the end of the tenth day after entry. " [DeCov]* | *Border control* | **Nationwide** | |
| 107 | | 28-03-2021 | *"No policy change. Source: http://web.archive.org/web/20210405124438/https://www.auswaertiges-amt.de/en/einreiseundaufenthalt/coronavirus http://web.archive.org/web/20210405124235/https://www.bundesgesundheitsministerium.de/service/gesetze-und-verordnungen/guv-19-lp/coronaschv/coronaschv-en.html" [OXF,454AC]* | *Border control (general policy)* | **Nationwide** | |

**Table S1 - NPI Descriptions** Date and descriptions of non pharmaceutical interventions in Germany from 01-10-2020 until 01-06-2021

| | Name | Date | Description | Labels | State | End Date |
|---|---|---|---|---|---|---|
| 108 | | 28-03-2021 | *"France, the Czech Republic and Slovakia are considered high incidence areas from 28 March 00:00. Persons who have stayed in a high incidence area within the last ten days before entering Germany are obliged to be tested before travelling to Germany. They must present a negative test result or an appropriate medical certificate to the carrier (for example the airline) before departure.*<br><br>*The negative test result may also be required during checks by the Federal Police (for example, entry checks at the airport or checks close to the border when entering by land at internal borders that are free of border controls). The swab for the test must have been taken no earlier than 48 hours before entry." [deCov2]* | *Border Control* | **Nationwide** | |
| 109 | **N12-a** | 30-03-2021 | *"From 30 March,* ***all persons traveling to Germany by air*** *must present a negative COVID-19 test result before embarking on their journey." [OXF, 456AC]*<br><br>*"The regulations for entering Germany have been supplemented by a general testing obligation for air travelers. The new compulsory testing applies from 30 March 0:00 up to and including 12 May 2021. A negative test result must then be available for air travelers before departure. The test must not be older than 48 hours. Only those who can provide a negative test result will be allowed to travel. Air travelers must pay for the test themselves. The Coronavirus Entry Regulation aims to limit the entry of corona infections. This is important against the background of the spread of new virus variants. In addition to these new regulations, the previous regulations still apply to all travelers when they enter the country by car, ship or train." [DeCov]* | *Border control* | **Nationwide** | |

**Table S1 - NPI Descriptions** Date and descriptions of non pharmaceutical interventions in Germany from 01-10-2020 until 01-06-2021

| | Name | Date | Description | Labels | State | End Date |
|---|---|---|---|---|---|---|
| 110 | **N12-b** | 31-03-2021 | *"In the fight against the Corona pandemic,* ***a general testing obligation now applies to all air travel to Germany****. The test must be taken* ***before take-off*** *in the country of departure. Anyone who cannot provide the airline with proof of a negative result will not be allowed to board the plane. The new requirements came into force at 0.00 a.m. on Tuesday night and are to apply until 12 May inclusive for the time being." [DeCov]* | *Border control* | **Nationwide** | |
| 111 | | 31-03-2021 | *"From 30 March, all persons traveling to Germany by air must present a negative COVID-19 test result before embarking on their journey. Source: http://web.archive.org/web/20210405124438/https://www.auswaertiges-amt.de/en/einreiseundaufenthalt/coronavirus "[OXF,456AC]* | *Border control (general policy)* | **Nationwide** | |
| 112 | | 01-04-2021 | *"****Bremen*** *(Germany) orders a hard lockdown for the easter days.* ***Businesses need to close*** *on Thursday and Saturday, except for supermarkets on Saturday.* ***Private meetings*** *are only allowed with two households and a maximum of 5 people." [DENPI, 4397G]* | *Workplaces, Gathering* | *Bremen* | |
| 113 | | 01-04-2021 | *"****Berlin*** *(Germany) enters a hard* ***lockdown for the easter*** *days. Thursday and Saturday all* ***businesses need to close****, except for supermarkets on Saturday.* ***Private meetings*** *are only allowed with one other household and a maximum of five people. Vacation within Germany is prohibited." [DENPI, 4398G]* | *Workplaces, Gathering, Internal movements* | *Berlin* | |
| 114 | | 01-04-2021 | *"****Saxony*** *(Germany) enters a hard* ***lockdown for the easter*** *days. Thursday and Saturday* ***all businesses need to close****,* | *Workplaces, Gatherin* | *Saxony* | |

| **Table S1 - NPI Descriptions** Date and descriptions of non pharmaceutical interventions in Germany from 01-10-2020 until 01-06-2021 | | | | | | |
|---|---|---|---|---|---|---|
| | **Name** | **Date** | **Description** | **Labels** | **State** | **End Date** |
| | | | *except for supermarkets on Saturday. Private meetings are only allowed with one other household and a maximum of five people. Vacation within Germany is prohibited." [DENPI, 4399G]* | *g, Internal movements* | | |
| 115 | | 10-04-2021 | *"No change in policy. Source: http://web.archive.org/web/20210417142312/https://www.bundesgesundheitsministerium.de/service/gesetze-und-verordnungen/guv-19-lp/coronaschv/coronaschv-en.html" [OXF, 467AC]* | *Border control (general policy)* | **Nationwide** | |
| 116 | | 13-04-2021 | *"No policy change. Source: http://web.archive.org/web/20210409091637/https://www.auswaertiges-amt.de/en/einreiseundaufenthalt/coronavirus http://web.archive.org/web/20210409091629/https://www.bundesgesundheitsministerium.de/service/gesetze-und-verordnungen/guv-19-lp/coronaschv/coronaschv-en.html" [OXF,462AC]* | *Border control (general policy)* | **Nationwide** | |
| 117 | | 23-04-2021 | *"No change in policy. Source: http://web.archive.org/web/20210424110356/https://www.bundesgesundheitsministerium.de/service/gesetze-und-verordnungen/guv-19-lp/coronaschv/coronaschv-en.html" [OXF, 480AC]* | *Border control (general policy)* | **Nationwide** | |
| 118 | | 25-04-2021 | *"No policy change. Source: http://web.archive.org/web/20210501151733/https://www.bundesgesundheitsministerium.de/service/gesetze-und-verordnungen/guv-19-lp/coronaschv/coronaschv-en.html" [OXF,482AC]* | *Border control (general policy)* | **Nationwide** | |
| 119 | | 09-05-2021 | *"Quarantine obligations will no longer apply for people who have been vaccinated or recovered –* | *Border control* | **Nationwide** | |

**Table S1 - NPI Descriptions** Date and descriptions of non pharmaceutical interventions in Germany from 01-10-2020 until 01-06-2021

| | Name | Date | Description | Labels | State | End Date |
|---|---|---|---|---|---|---|
| | | | *for instance when they enter Germany from another country. Quarantine regulations will still apply, however, if they are entering Germany from an area classed as an "area of variant of concern". Source: http://web.archive.org/web/20210521090533/https://www.bundesregierung.de/breg-en/news/restrictions-eased-for-vaccinated-persons-1911192" [OXF,496AC]* | *(general policy)* | | |

Information sources for the NPIs are

DENPI CoronaNetDataScience/corona_tscs: Releasing new data fields in event dataset | Zenodo https://github.com/CoronaNetDataScience/corona_tscs/blob/master/data/CoronaNet/data_country/coronanet_release/coronanet_release_Germany.csv, accessed at 2022.03.03

OXFNPI Thomas Hale, Noam Angrist, Rafael Goldszmidt, Beatriz Kira, Anna Petherick, Toby Phillips, Samuel Webster, Emily Cameron-Blake, Laura Hallas, Saptarshi Majumdar, and Helen Tatlow. (2021). "A global panel database of pandemic policies (Oxford COVID-19 Government Response Tracker)." Nature Human Behaviour. https://doi.org/10.1038/s41562-021-01079-8, accessed at 2022.02.25

deCov1 "Latest coronavirus updates", https://www.deutschland.de/en/news/coronavirus-in-germany-information, accessed at 2022.05.06.

deCov2 "The Federal Government informs about the corona crisis", https://www.deutschland.de/en/news/german-federal-government-informs-about-the-corona-crisis, accessed at 06-05-2022.

16 Zhang H, John R, Peng Z, *et al.* The relationship between species richness and evenness in plant communities along a successional gradient: a study from sub-alpine meadows of

the Eastern Qinghai-Tibetan Plateau, China. *PLoS One* 2012; **7**: e49024.
